# Genome-wide association analyses of symptom severity among clozapine-treated patients with schizophrenia spectrum disorder

**DOI:** 10.1101/2021.06.06.21258200

**Authors:** C. Okhuijsen-Pfeifer, M.Z. van der Horst, C.A. Bousman, B. Lin, K.R. van Eijk, S. Ripke, Y. Ayhan, M.O. Babaoglu, M. Bak, W. Alink, H. van Beek, E. Beld, A. Bouhuis, M. Edlinger, I.M. Erdogan, A. Ertuğrul, G. Yoca, I.P. Everall, T. Görlitz, GROUP Genetic Risk and Outcome of Psychosis investigators, K.P. Grootens, S. Gutwinski, T. Hallikainen, E. Jeger-Land, M. de Koning, M. Lähteenvuo, S.E. Legge, S. Leucht, C. Morgenroth, A. Müderrisoğlu, A. Narang, C. Pantelis, A.F. Pardiñas, T. Oviedo-Salcedo, J. Schneider-Thoma, S. Schreiter, E. Repo-Tiihonen, H. Tuppurainen, M. Veereschild, S. Veerman, M. de Vos, E. Wagner, D. Cohen, J.P.A.M. Bogers, J.T.R. Walters, A.E. Anil Yağcıoğlu, J. Tiihonen, A. Hasan, J.J. Luykx

## Abstract

Clozapine is the most effective antipsychotic for patients with treatment-resistant schizophrenia. However, response is highly variable and possible genetic underpinnings of this variability remain unknown. Here, we performed polygenic risk score (PRS) analyses to estimate the amount of variance in symptom severity among clozapine-treated patients explained by PRSs (R2) and examined the association between symptom severity and genotype-predicted *CYP1A2, CYP2D6*, and *CYP2C19* enzyme activity. Genome-wide association (GWA) analyses were performed to explore loci associated with symptom severity. A multicenter cohort of 804 patients (after quality control N=684) with schizophrenia spectrum disorder treated with clozapine were cross-sectionally assessed using the Positive and Negative Syndrome Scale and/or the Clinical Global Impression-Severity (CGI-S) scale. GWA and PRS regression analyses were conducted. Genotype-predicted *CYP1A2, CYP2D6*, and *CYP2C19* enzyme activities were calculated. Schizophrenia-PRS was most significantly and positively associated with low symptom severity (*p*=1.03×10^−3^; R2=1.85). Cross-disorder-PRS was also positively associated with lower CGI-S score (*p*=0.01; R2=0.81). Compared to the lowest tertile, patients in the highest schizophrenia-PRS tertile had 1.94 times (*p*=6.84×10^−4^) increased probability of low symptom severity. Higher genotype-predicted *CYP2C19* enzyme activity was independently associated with lower symptom severity (*p*=8.44×10^−3^). While no locus surpassed the genome-wide significance threshold, rs1923778 within NFIB showed a suggestive association (*p*=3.78×10^−7^) with symptom severity. We show that high schizophrenia-PRS and genotype-predicted *CYP2C19* enzyme activity are independently associated with lower symptom severity among individuals treated with clozapine. Our findings open avenues for future pharmacogenomic projects investigating the potential of PRS and genotype-predicted *CYP-*activity in schizophrenia.

## Introduction

About one third of patients with schizophrenia is considered to have treatment-resistant schizophrenia (TRS).^1^ TRS has been defined as the persistence of symptoms despite at least two trials of antipsychotic medications of adequate dose and duration with documented adherence.^2–4^ For patients with TRS, clozapine is the most effective antipsychotic drug.^5,6^ However, 40% of TRS-patients achieve no sufficient response to clozapine, suggesting that up to 20% of schizophrenia patients are ultra-resistant (defined as failure to respond to adequate trials of two antipsychotics and clozapine).^7^ Despite its efficacy, the mean delay in clozapine prescription reaches up to 9 years,^8–10^ which in turn is associated with poor treatment outcomes and less functional recovery.^11,12^ Elucidating determinants of symptom severity while on clozapine may contribute to early identification of those more likely to be responsive to clozapine, enabling patients to start clozapine earlier in their disease course, resulting in a better quality of life, increased life expectancy, and lower economic burden.^13,14^ This is all the more important given the evidence that clozapine as first-or second-line therapy is more effective than other antipsychotics.^11,15,16^ In addition, patients who are less likely to be responsive to clozapine may delay clozapine treatment, avoiding unnecessary potential side effects and blood monitoring.

A promising strategy for the identification of genetic variation associated with symptom severity among clozapine users, is the genome-wide association study (GWAS) that also allows to generate data for polygenic risk scoring (PRS). Several GWASs have identified candidate loci for clozapine blood concentrations and severe adverse drug reactions associated with clozapine, but limited focus has been given to symptomatic outcomes in patients using clozapine.^17–21^ To our knowledge, there has only been one study that used a genome-wide approach to examine clozapine treatment outcome.^22^ In that study of 123 clozapine-treated individuals, no statistically significant differences in schizophrenia-PRS between responders and non-responders were detected (N=123, *p*=0.06).^22^ PRS analyses performed to uncover differences between responders and non-responders of other antipsychotics have yielded inconsistent results.^23–26^

Another promising approach that complements GWAS and PRS, is the examination of known haplotype variation in genes associated with clozapine metabolism. *CYP1A2* is considered the primary metabolism pathway for clozapine with secondary contributions from *CYP3A4/5, CYP2D6* and *CYP2C19*.^27^ Clozapine metabolism differs between persons and as a result, clozapine blood concentrations among individuals prescribed the same dose may vary. Because of this variation, therapeutic drug monitoring of clozapine blood concentrations is routinely applied. Previous studies have suggested that clozapine blood concentrations ≥350 ng/mL are associated with superior symptomatic outcome.^28^ However, determining the required dose for an individual to achieve this target blood concentration and symptomatic relief can be challenging. As such, several studies have used haplotype variation in *CYP1A2, CYP2C19*, and *CYP2D6* as markers of an individual’s capacity to metabolize clozapine and their probability of receiving symptomatic relief.^29–32^ Although results of these studies have been inconsistent, recent findings suggest these inconsistencies could be a result of phenoconversion, a phenomenon in which an individual’s genotype-predicted drug metabolism does not reflect their observed metabolism, due to the presence of non-genetic factors such as concomitant medications (e.g. (es)citalopram) and smoking behavior.^33^ Thus, additional investigation to identify associations between genotype-predicted drug metabolism and clozapine treatment outcome is warranted.

Given the lack of established knowledge about genetic mechanisms underlying clozapine treatment outcome, this is the first study in patients with severe schizophrenia that dissects associations between symptom severity and genome-wide data. An international team with a range of different backgrounds joined forces several years ago, resulting in a unique and the largest dataset of clozapine users with genome-wide and symptom severity data available. The aims of the study were to analyze the amount of variance in symptom severity among clozapine-treated patients explained by PRSs, to examine the association between symptom severity and genotype-predicted *CYP1A2, CYP2D6*, and *CYP2C19* enzyme activity, and explore loci associated with symptom severity using GWA analyses.

## Methods

### Participants

Participants (N=804) came from five independent cohorts: 470 participants were recruited by the Clozapine International (CLOZIN)^34–37^ consortium in the Netherlands, Germany, Austria, and Finland; 174 participants by the Genetic Risk and Outcome of Psychosis (GROUP) consortium in the Netherlands; 80 participants by the Cooperative Research Centre (CRC) in Australia; 50 participants by Hacettepe University in Turkey; and 30 participants by Mental Health Services Rivierduinen in the Netherlands. All studies were approved by their respective local Institutional Review Boards and all participants provided written informed consent prior to participation. The studies were compliant with the Declaration of Helsinki (2013).^38^ Participants were included if they: (1) were aged 18 years or older, (2) had a primary diagnosis of schizophrenia spectrum disorder (SSD), and (3) were using clozapine (no minimum duration of treatment). The eligibility criteria were not strict to represent ‘real world’ patients, as this is valuable for clinical value and applicability. Clozapine blood levels were measured in local accredited laboratories, ∼12 hours after the last clozapine dose intake. More information about each cohort is provided in the Supplementary Methods.

### Phenotyping

Symptom severity was assessed by treating physicians or trained study raters using the Clinical Global Impression-Severity (CGI-S) scale and/or the Positive and Negative Syndrome Scale (PANSS).^39,40^ Our main outcome was symptom severity defined as a quantitative measure and our secondary outcome was symptom severity defined as a binary measure (low vs. high symptom severity). See Supplementary Methods for more detailed information.

### Genotyping and quality control

Genotyping and quality control followed standard procedures and are described in Supplementary Methods.

### Statistical analysis

#### Genome-wide association analyses

Statistical analyses were conducted using PLINK v1.90b3z 64-bit and R version 3.2.2 (14 Aug 2015; http://www.r-project.org/) software packages. Explorative GWAS was conducted using linear regression for quantitative outcome and logistic regression for binary outcome. The quantitative outcome analysis was thus a quantitative trait locus (QTL) analysis, conducted similarly to previously reported.^41,42^ We performed GWA analyses of both outcomes, correcting for sex, age, age-squared, and the first 10 genetic-ancestry principal components (PCs). These analyses were conducted in our entire multi-ethnic cohort to assure diversity and inclusiveness of non-North Western European people.^43^ However, to evaluate the robustness of our findings, as sensitivity analyses, we repeated all analyses after removing participants deviating more than 3 standard deviations (SD) from the means of the first four PCs, based on the HapMap3 HRC r1.1 2016 (GRCh37/hg19) population (N=1397; Supplementary Figure 1). Genome-wide significance was set at *p*<5×10^−8^ and suggestive significance at *p*<5×10^−5^.

Post-GWAS analysis was performed for identification and annotation of independent associations within our data, using Functional Mapping and Annotation of genetic associations (FUMA)^44^ and Hi-C coupled Multi-marker Analysis of GenoMic Annotation (H-MAGMA v1.08; Supplementary Methods).^45^

#### Polygenic risk score analyses

PRS is an estimate of an individual’s polygenic liability to a certain trait.^46^ PRSs were calculated for the following three traits relevant for schizophrenia or clozapine metabolism, using the most recent GWAS’ summary statistics: schizophrenia,^47^ cross-disorder,^48^ and clozapine metabolism.^17^ The PRSs were corrected for sex, age, and 10 PCs, and the Bonferroni-corrected significance level was *p*<0.017 (see Supplementary Methods and Supplementary Table 2 for details).

#### Genotype-predicted enzyme activity score analysis

Multiple drug metabolizing enzymes contribute to the demethylation and oxidation of clozapine (physiologically active compound) to N-desmethylclozapine (“norclozapine”; putatively active metabolite) and clozapine n-oxide (considered to be an inactive metabolite).^27,49,50^ However, *CYP1A2*, and to a lesser extent *CYP2C19* and *CYP2D6*, are considered the primary clozapine metabolizing enzymes *in vivo*.^27^ Imputed genotype data (see ‘Supplementary Methods - *Genotyping and quality control’*) was subjected to Stargazer v1.08^51^ to call *CYP*-haplotypes (star alleles). Activity scores for *CYP2D6* were based on translation tables maintained by the Pharmacogene Variation (PharmVar) Consortium^52^ and the Pharmacogenomics Knowledgebase (PharmGKB), whereas activity scores for *CYP1A2* and *CYP2C19* followed previously published scoring methods (Supplementary Methods and Supplementary Table 4).^53–55^

Prior to analysis, *CYP-*activity scores were corrected for concomitant inhibitors or inducers of each of the corresponding genes using previously employed phenoconversion methodology (Supplementary Methods and Supplementary Table 5).^33^ To determine if *CYP-* activity scores were associated with symptom severity outcomes, logistic and linear regression models were fitted, with age, sex, dose-adjusted clozapine levels (i.e. concentration-to-dose ratio, one measurement per participant), and duration of clozapine therapy included as covariates.^33^ In addition, linear regression models were fitted to estimate the amount of variance in dose-adjusted clozapine levels that was explained by *CYP2C19, CYP1A2*, and *CYP2D6*. Levels of N-desmethylclozapine were unavailable. The Bonferroni-corrected significance level was *p*<0.017, as we corrected for three independent regression analyses performed. *Post-hoc* models that included results from the PRS-analyses were fitted to test the independency of PRS and *CYP* associations.

## Results

### Genome-wide association analysis

684 individuals and 5,506,411 SNPs passed the QC and were included in the GWAS. There were 330 participants with low and 354 with high symptom severity (see Supplementary Table 6 for demographic and clinical characteristics). Linkage disequilibrium (LD)-intercept scores and genomic inflation correction factors (λGC) pre- and post-imputation were all <1.02, suggesting no inflation of the test statistics (Supplementary Figure 2).

No genome-wide significant hits were identified. The most significantly associated locus was detected between quantitative outcome and intronic rs1923778 on chromosome 9 (*p=*3.78×10^−7^; Figure 1A, Supplementary Figure 3A & Supplementary Table 7), within *nuclear factor 1 B-type (NFIB)*. We did not detect dose-adjusted clozapine level differences between genotype groups at this locus, rendering it unlikely that the association we found is merely a proxy for association with dose-adjusted clozapine levels (Supplementary Figure 4 & Supplementary Table 8). The most significant locus for binary outcome was intronic rs4742565 on chromosome 9 (*p=*1.64×10^−6^; Figure 1B, Supplementary Figure 3B & Supplementary Table 7), within *protein tyrosine phosphatase receptor type D (PTPRD)*. In sensitivity analyses, results remained similar (Supplementary Results). LD between the top quantitative locus and the top binary locus was R^2^=0.0010, D’=0.092 and the loci were 5 Mb apart.

**Figure 1A&B.**
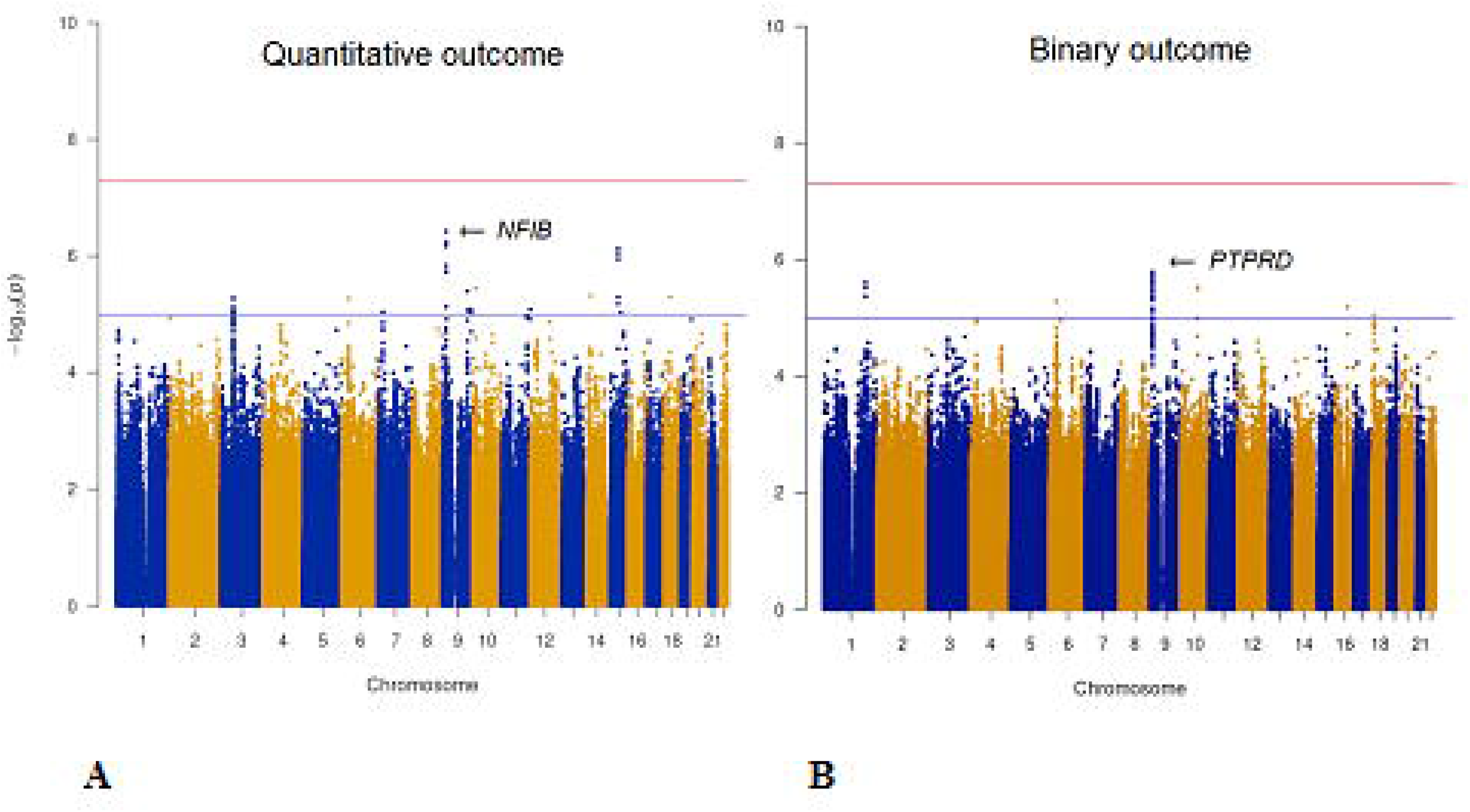
Manhattan plots depicting the genome-wide association results of symptom severity while on clozapine for quantitative (A) and binary outcome (B). The X-axis shows the chromosomal positions. The Y-axis shows –log10 (*p*-values). The red line illustrates the genome-wide significance level of *p=*5×10^−8^, and the blue line illustrates the suggestive level of significance of *p=*5×10^−5^. The arrows indicate the top loci and the closest genes.

Post-GWAS analyses using FUMA indicated significantly enriched differentially expressed gene sets for the hypothalamus and hippocampus for quantitative outcome (Supplementary Results, Supplementary Figure 5A) and expression in the brain of the prioritized genes for binary outcome (Supplementary Figure 5B). We did not find any significantly associated target genes using H-MAGMA (Supplementary Figure 6A&B).

### Polygenic risk score analysis

The same 684 individuals were included in PRS-analyses. Schizophrenia-PRS was significantly associated with binary outcome (*p*=1.03×10^−3^, R^2^ =1.85, p_t_=0.4, Figures 2A&3A). Patients in the highest schizophrenia-PRS tertile had 1.94 times (*p*=6.84×10^−4^, 95% confidence interval (CI)=1.33-2.81) increased chances of low symptom severity while on clozapine compared to patients in the lowest schizophrenia-PRS tertile (Figure 2B, Supplementary Table 9; Supplementary Table 10). Similarly, patients in the highest schizophrenia-PRS decile had 2.26 times (*p*=3.96×10^−3^, 95% CI=1.30-3.91) increased chances of low symptom severity while on clozapine compared to patients in the lowest schizophrenia-PRS decile (Figure 2B, Supplementary Table 9; Supplementary Table 10). Cross-disorder-PRS was significantly associated with quantitative outcome (*p*=0.01, R^2^ = 0.81, p_t_=0.3, Figure 3B). Other PRS association results were not significant (Supplementary Results, Supplementary Figure 7A-F). In sensitivity analyses results remained similar (Supplementary Results).

**Figure 2A&B.**
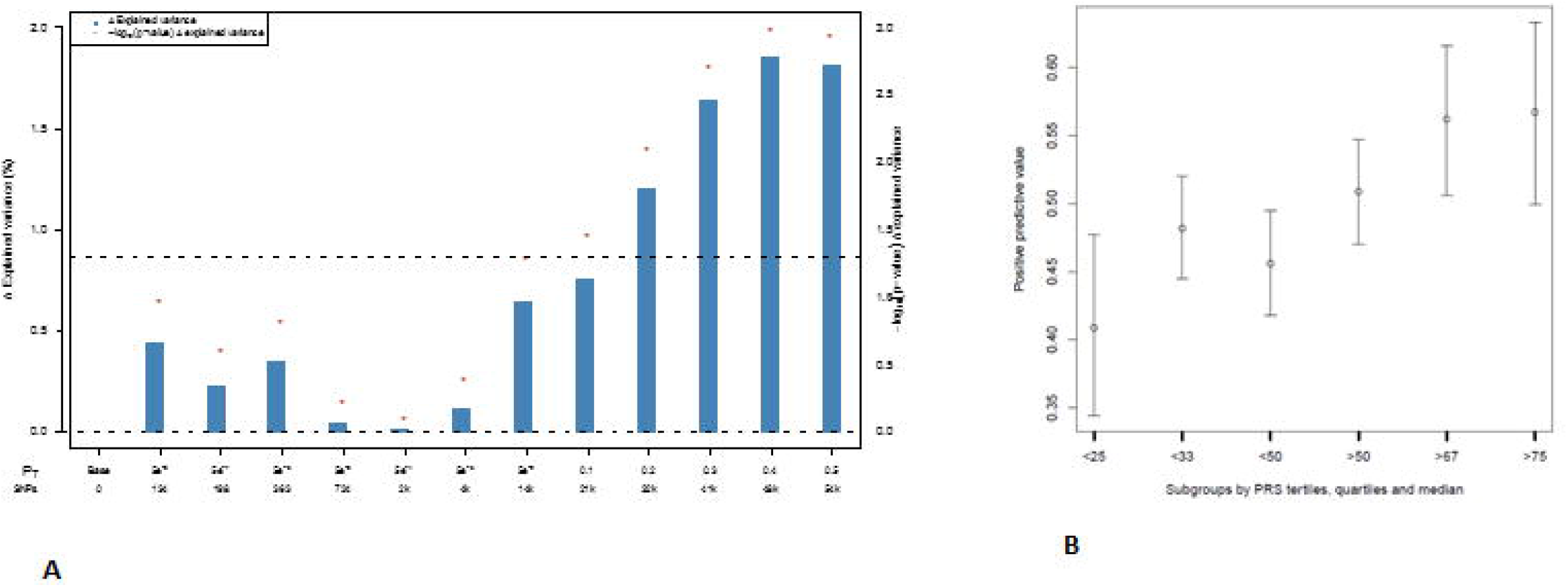
Bar plot illustrating the explained variance for the association of schizophrenia-PRS with binary outcome at several p_t_, adjusted for sex, age, and 10 PCs. p_t_ are displayed on the X-axis, where the number of included SNPs increases with more lenient p_t_. Δ Explained variance represents the Nagelkerke R^2^ (shown as %). The red dots represent the significance of the association results (-Log 10 *p*-value). The dashed line represents a nominal significance-level of *p*<0.05 (A). Individual risk prediction: higher schizophrenia-PRS was associated with higher positive predictive value for low symptom severity. Whiskers represent confidence intervals (±1.96×standard error) around the central positive predictive value estimate (B).

**Figure 3A&B.**
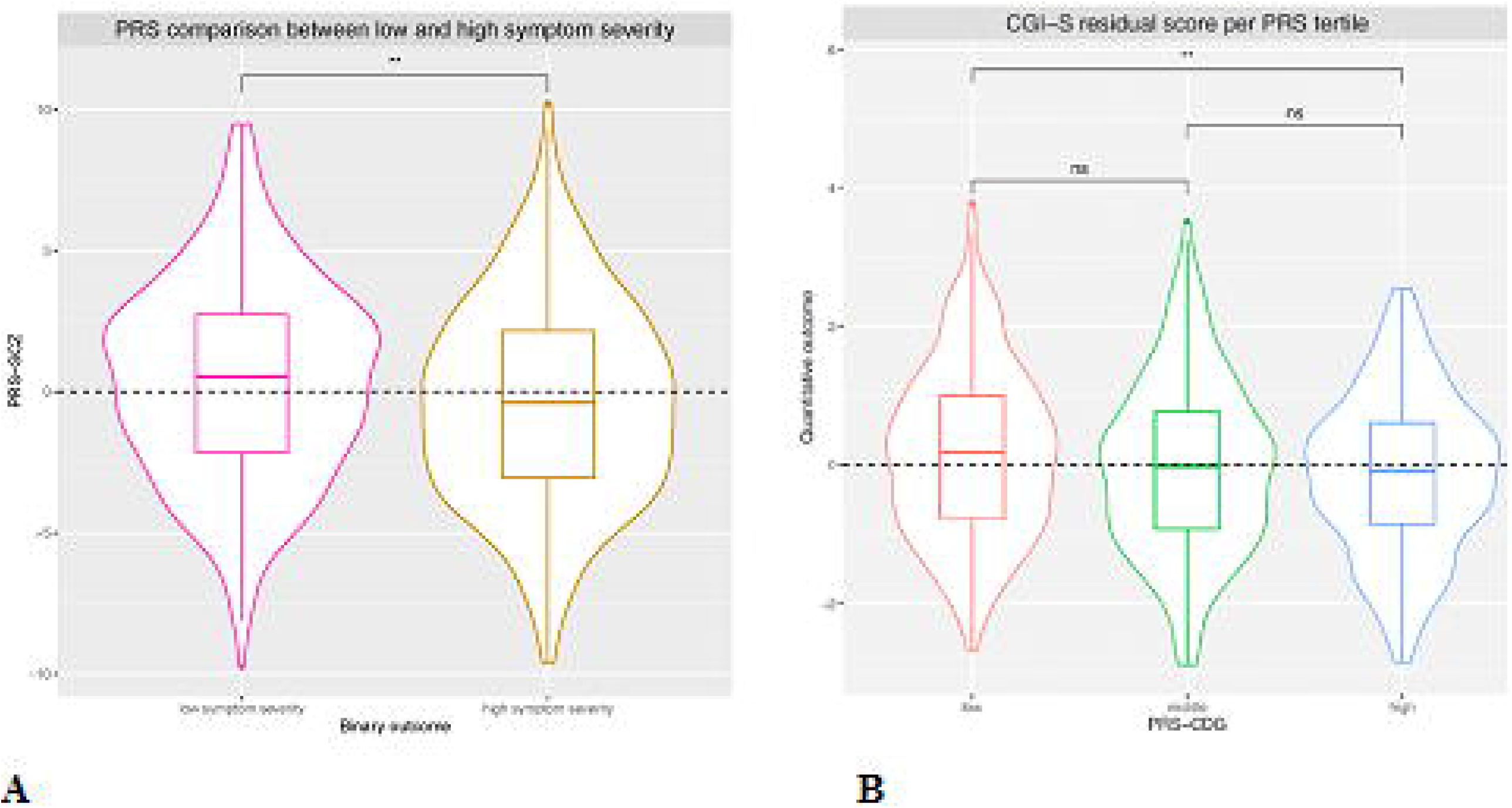
Violin plots of schizophrenia-PRS (PRS-SCZ) comparison for binary outcome (A), and cross-disorder-PRS (PRS-CDG) tertile comparison for quantitative outcome (B). For both analyses the best fitting p_t_ was used. The dashed line illustrates the mean PRS-SCZ (A) and the mean residual CGI-S score (B) in all participants. Differences were determined by linear regression of quantitative outcome on PRS tertile and using a T-test for binary outcome, corrected for sex, age, and 10 PCs (Supplementary Methods). \*\**p*<5.0×10^−3^. *Abbreviation:* ns=non-significant.

### Genotype-predicted enzyme activity score analyses

Higher *CYP2C19* activity score was significantly associated with greater probability of low symptom severity (odds ratio (OR)=1.59, 95% CI=1.13-2.24, *p=*8.44×10^−3^, N=291, Supplementary Table 11, Figure 4A) and was associated in the same direction at a higher *p*-value with quantitative outcome (beta=-0.10, *p=*0.10; Supplementary Table 12, Figure 4B) but was not associated with dose-adjusted clozapine levels (Supplementary Table 13, Figure 4C). The association between *CYP2C19* activity score and the probability of low symptom severity did not change when including schizophrenia-PRS (OR=1.59, 95% CI=1.13-2.24, *p*=8.51×10^−3^, Supplementary Table 14), schizophrenia-PRS and the first 10 PCs (OR=1.63, 95% CI=1.97-2.43, *p*=0.02, Supplementary Table 15), or the top two GWAS hits (*NFIB* rs1923778, *PTPRD* rs4742565) (OR=1.60, 95% CI=1.06-2.42, *p*=0.02, Supplementary Table 16).

**Figure 4A - I.**
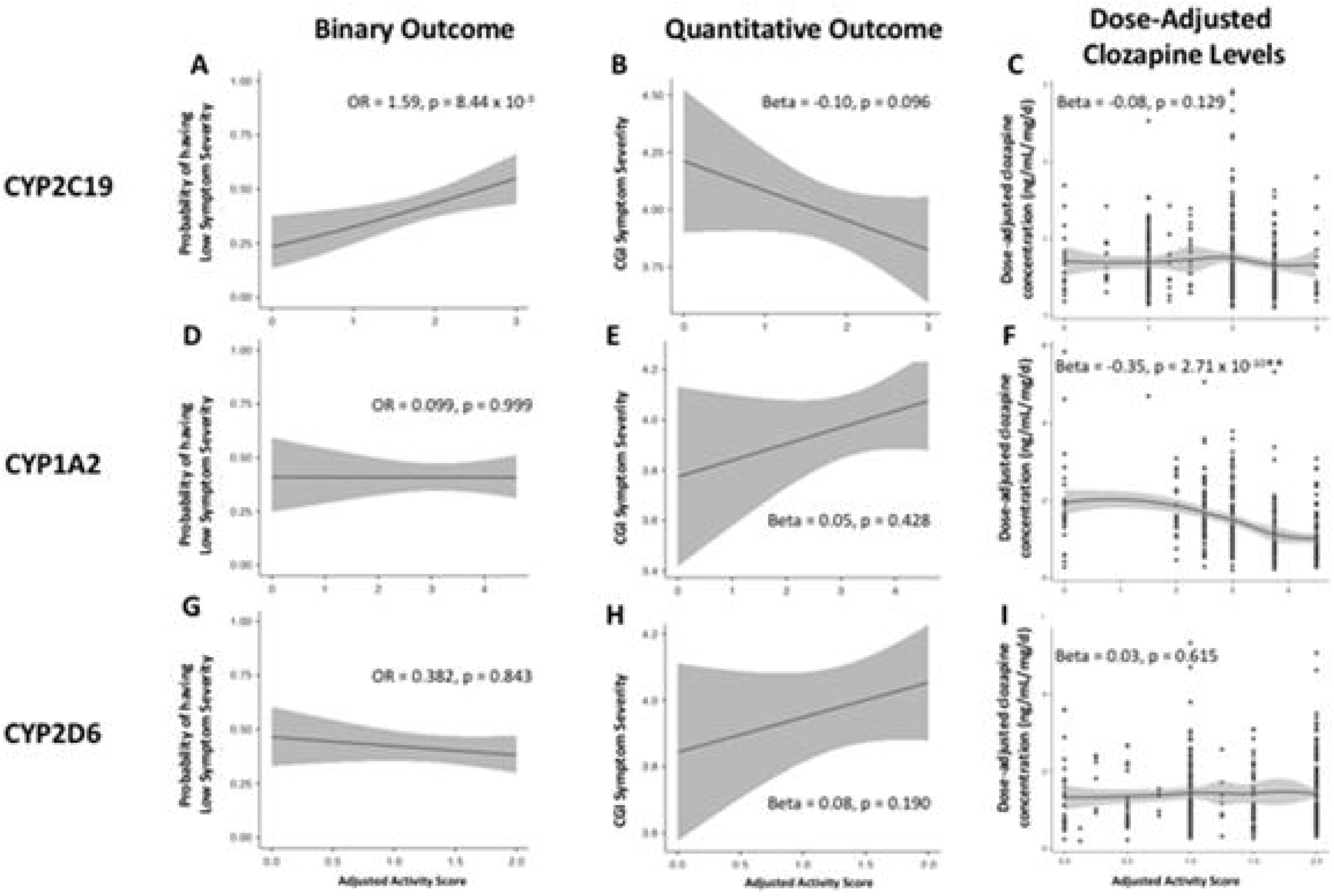
Association of corrected *CYP2C19, CYP1A2*, and *CYP2D6* genotype-predicted activity scores with symptom severity while on clozapine and dose-adjusted clozapine levels.

*CYP1A2* activity score was not associated with our binary (Figure 4D) or quantitative (Figure 4E) outcomes but, as expected, was inversely correlated with dose-adjusted clozapine levels (beta=-0.35, *p=*2.71×10^−10^; Supplementary Table 13, Figure 4F). *CYP2D6* activity score was not associated with either symptom severity or dose-adjusted clozapine levels (Figure 4G-I). Neither *NFIB* rs1923778 (beta=-0.08, *p*=0.47) nor *PTPRD* rs4742565 (beta=0.07, *p*=0.26) were associated with dose-adjusted clozapine levels. No interaction between *CYP2C19* and schizophrenia-PRS was found for binary outcome (beta=0.04, *p*=0.17). It is important to note that the C/D ratios were significantly higher in the Hacettepe cohort, compared to the other cohorts (Supplementary Table 8). In sensitivity analyses results remained similar, indicating that ancestry does not influence the results and conclusions (Supplementary Results). Furthermore, as expected, there was no difference in the dose-adjusted clozapine concentrations between people with low symptom severity (mean=1.37, SD=0.86) and high symptom severity (mean=1.42, SD=0.91; t=0.57, *p*=0.57; Supplementary Results, Supplementary Table 17&18, Supplementary Figure 13).

## Discussion

To our knowledge, this is the first study using a comprehensive, genome-wide approach to examine the genomic underpinnings of symptom severity among individuals with SSD treated with clozapine (N=804). Using a novel approach of integrating genome-wide, PRS, and *CYP* analyses, we demonstrate that higher schizophrenia-PRS and higher genotype-predicted *CYP2C19* enzyme activity are independently associated with lower symptom severity while on clozapine.

Although no significant genome-wide hit was discovered, the loci on *NFIB* (rs1923778, *p=*3.78×10^−7^) and *PTPRD* (rs4742565, *p=*1.64×10^−6^) are of interest given previous literature. *NFIB* is a protein coding gene associated with embryonic development, tumor growth, and brain development.^56^ A recent GWAS of clozapine levels found a different locus on *NFIB* (rs28379954; Supplementary Table 19) to be associated with clozapine levels (*p*=1.68×10^−8^) and risk of subtherapeutic serum concentrations.^17^ *PTPRD* is a protein coding gene associated with several brain phenotypes and disorders, such as obsessive-compulsive disorder, autism spectrum disorder, and substance use disorders.^57–62^ Moreover, a GWAS of response to the atypical antipsychotic lurasidone found *PTPRD* to be suggestively associated with treatment response in European and African participants (*p=*6.03×10^−5^ and *p=*4.29×10^−5^, respectively).^23^

Our PRS analyses revealed that higher schizophrenia-PRS was associated with lower symptom severity as a binary trait and that higher cross-disorder-PRS was associated with lower symptom severity as a quantitative trait. To our knowledge, no previous study has examined the association between cross-disorder-PRS and antipsychotic treatment outcome. However, several studies have evaluated the association between schizophrenia-PRS and antipsychotic treatment outcome,^22–26^ although only one of these studies examined clozapine response.^22^ In line with our results, that study reported higher schizophrenia-PRS in clozapine responders compared to clozapine non-responders, although this was not significant (*p=*0.06).^22^ Differences in phenotyping (here we used PANSS and CGI-S scales; in the previous study a non-validated, 4-level ordinal scale was used) and power (N=684 vs. N=123) possibly explain differences in statistical significance (*p*=1.03×10^−3^ vs. *p*=0.06). The same direction of association was found in a GWAS of lurasidone response (N=429)^23^ and a risperidone response GWAS.^24^ However, a study with first-episode psychosis patients using several antipsychotics (but not clozapine) found that higher schizophrenia-PRS was associated with lower response rates (i.e. higher symptom severity).^63^ Several studies indicate that schizophrenia-PRS increases when comparing first-episode psychosis to schizophrenia, and schizophrenia to TRS^22,63–65^, although there is also conflicting evidence for the latter.^66,67^ Bearing those former observations in mind, higher schizophrenia-PRS may characterize a subset of TRS-patients more likely to respond well to clozapine. Speculatively, people with low schizophrenia-PRS may thus have a better prognosis with shorter duration of illness, whereas in advanced stages of illness those with high schizophrenia-PRS may respond better to clozapine.^65^ Alternatively, it is also plausible that people with higher schizophrenia-PRS are genetically more close to schizophrenia and therefore more responsive to clozapine, while people who are more genetically distant and/or where other environmental factors exert an influence are less responsive to clozapine.

Furthermore, we found a positive association between genotype-predicted *CYP2C19* enzyme activity and symptom severity. This finding aligns with a previous study of 137 clozapine-treated patients that reported *CYP2C19* ultrarapid metabolizers (*\*17/*17*) were five times more likely to show clinical improvements,^68^ although smaller studies have shown no association^33, 69^ or an inverse association^70^ between *CYP2C19*17* carriers and clozapine-related symptomatic outcomes. Although *CYP2C19* is involved in the demethylation of clozapine to N-desmethylclozapine, a pharmacologically active metabolite that binds to an array of receptors including dopamine D2 and D3 receptors, muscarinic receptors and serotonergic receptors^27^, current evidence suggests *CYP2C19* genetic variation has limited effect on clozapine metabolism.^33,71,72^ As such, the association between *CYP2C19* enzymatic activity and improvement in clinical outcome is unlikely to be explained by differential clozapine or norclozapine blood concentrations. In fact, we did not detect an association between *CYP2C19* enzyme activity and clozapine concentrations. Alternatively, our findings may reflect *CYP2C19*’s role in metabolizing endogenous compounds in the brain.^73^ Human studies have shown increased *CYP2C19* genotype-predicted activity was associated with smaller hippocampi volumes and greater suicidality,^74^ although attempts to replicate these associations have been unsuccessful.^75^ Thus, we cannot rule out that our finding represents a type 1 error. A weakness of our study is the lack of a replication cohort and consequently our findings await replication in larger datasets with diverse ancestries to more firmly guard against the risk of type-I error that is inherent in research projects (such as ours) where several statistical tests are performed.

Therefore, future investigations of the relationships between genotype-predicted *CYP2C19* enzyme activity and symptomatic outcomes are warranted. In such future studies, norclozapine should also be taken into account to allow a separate genetic study on such levels and ratios with clozapine. Additionally, it is interesting that in the current study no association between *CYP1A2* and symptom severity is found, as this was suggested from a previous study.^33^ Perhaps this difference in findings is explained by the use of different measurements of symptom severity, as the previous study used PANSS,^33^ while the current study used CGI-S which is less specifically about psychotic symptoms. It is good to note, there is still little understanding of the relationship between *CYP1A2* genotype and symptom severity, and this should be investigated in more detail.

Strengths of our study include our unique sample of over 800 multi-ethnic patients with SSD using clozapine, and the use of a very recent GWAS-platform, and genotype-predicted activity of relevant metabolizing enzymes. Furthermore, although no longitudinal data on clozapine response rates was available for our participants, the distribution of low and high symptom severity while on clozapine in our cohort (N=330 vs N=354, respectively) aligns with reported response rates to clozapine.^7^ Nonetheless, several limitations should be considered when interpreting our results. First, participants were cross-sectionally ascertained using two symptom scales, which made the use of a conversion table necessary. The lack of inflation of our GWAS test statistic as well as the consistency between the results of the main and sensitivity analyses’ nonetheless support the robustness of the results. Second, our study population is derived from multiple cohorts and due to incomplete covariate information, we were unable to enter some into our GWAS. The possible effect of previous antipsychotic medication and concomitant therapies should be of interest to future GWASs. In such GWASs, patients stopping clozapine due to poor clinical effect should be targeted as a separate group, as this could provide highly valuable genetic information on ultra-TRS. Third, we estimate that about 3% of our cohort were incorrectly classified as *CYP2D6* normal metabolizers due to the absence of copy number variation data required to detect *CYP2D6* ultrarapid metabolizers (Supplementary Methods). However, to our knowledge, there is no evidence suggesting this misclassification error would have a meaningful impact on our study findings. Fourth, as the GROUP cohort is an older cohort, there might be sample overlap between the current and the PGC-schizophrenia study population and consequently the PGC-cross-disorder study population. We therefore repeated our analyses excluding the samples from GROUP: all PRS association patterns remained similar with the same p_t_, albeit less significant, as expected due to loss of power (Supplementary Figure 9A&B). Furthermore, there were no clozapine concentrations available from GROUP. Therefore, this cohort was not represented in the *CYP*-analyses. Another limitation is that we could not definitely determine for all cohorts whether clozapine concentrations represented steady state and trough levels nor could we verify adherence. Nonetheless, as our observed clozapine C/D ratios follow the expected pattern of steady-state and trough levels (Supplementary Figure 14A-D), it is likely that these levels are reliable approximations. Inflammation (e.g., C-reactive protein) and caffeine consumption data, which can inhibit *CYP1A2* activity, were not available and could therefore be over-estimated for some participants. Finally, caution in interpretation of our GWAS results is warranted given the lack of a replication cohort.

In conclusion, we demonstrate for the first time that higher schizophrenia-PRS and *CYP2C19* predicted activity are independently associated with low symptom severity among clozapine-treated schizophrenia patients. For future clinical translation, if these findings are replicated, schizophrenia-PRS and genotype-predicted *CYP2C19* activity may be used in conjunction with non-genetic factors to help predict clozapine response, ultimately allowing early identification of individuals more likely responding to clozapine. Such future studies should also incorporate deeper phenotyping information collected (e.g. clinical symptomatology in further detail, steady-state clozapine levels, adherence) in a longitudinal design. Timely prescribing may improve patients’ prognosis, given clozapine’s superiority over other antipsychotics in early disease stages.^15^

## Data Availability

Data is available on request

## Conflicts of interest

Dr. Bousman reports he has received in-kind testing kits from Myriad Neuroscience, CNSDose, Genomind, and AB-Biotics for research purposes but has not received payments or received any equity, stocks, or options in these companies or any other pharmacogenetic companies. Dr. Lähteenvuo reports being a shareholder and board member at Genomi Solutions Ltd and Nursie Health Ltd; receiving research grants or awards from the Finnish Medical Foundation and Emil Aaltonen Foundation; receiving travel grants or speakers’ honoraria from Sunovion Ltd, Janssen-Cilag, Otsuka Pharmaceutical Ltd, Lundbeck Ltd, Orion Pharma ltd; and working as a coordinator for a research project funded by the Stanley Foundation. Dr. A.E. Anil Yagcioglu has recently received travel grants or speakers’ honoraria from Janssen, Otsuka; is a member of the advisory board for Janssen, Otsuka; is currently participating in an international clinical trial funded by Janssen. Dr. Tiihonen reports personal fees from the Finnish Medicines Agency (Fimea), European Medicines Agency (EMA), Eli Lilly, Janssen, Lundbeck, and Otsuka; is a member of the advisory board for Lundbeck; and has participated in research projects funded by grants from Eli Lilly and Janssen to his employing institution. All other authors have declared that there are no conflicts of interest in relation to the subject of this study.

## Funding

Dr. Bousman reports a grant from Alberta Innovates Strategic Research Project G2018000868, during the conduct of the study. Dr. Pardiñas’ work is supported by a “Springboard” award by the Academy of Medical Sciences (SBF005\1083). Dr Luykx’ work is supported by a personal grant from the Rudolf Magnus Young Talent Fellowship program of the University Medical Center Utrecht.

## Acknowledgments

We would like to thank R.L. Cummins, R. Kahn, M.K. Bakker, E. Bekema, H. Gijsman, A. Jongkind and P. Kleymann for their valuable input (see Supplementary Acknowledgements for details).

## Notes

### Clinical Trial

NCT03253367

### Author Declarations

This paper concerns data of 5 cohorts, that received separate approvals. CLOZIN cohort & GROUP cohort: Ethical Review Board Utrecht (local name: Medisch Ethische Toetsingscommissie Utrecht (submitted as and approved with numbers 15-306/15-312 & 04-003) CRC: Melbourne Health Human Research Ethics Committee (approved with number MHREC ID 2012.069) Hacettepe cohort: Hacettepe University Faculty of Medicine Research Ethics Committee (approved with number FON 11/21) MHS: Ethical Review Board Free University Amsterdam Medical Center (local name: Medisch Ethische Toetsingscommisue Vrije UUniversiteit Medisch Centrum (VUmc)) (approved with number D3-201)

### Summary of Updates

We got comments from reviewers to clarify some minor points.

## References

1. Lally J, MacCabe JH. Antipsychotic medication in schizophrenia: A review. Br Med Bull. Published online 2015. doi:10.1093/bmb/ldv017

2. Howes OD, McCutcheon R, Agid O, et al. Treatment-Resistant Schizophrenia: Treatment Response and Resistance in Psychosis (TRRIP) Working Group Consensus Guidelines on Diagnosis and Terminology. Am J Psychiatry. 2017;174(3):216–229. doi:10.1176/appi.ajp.2016.16050503

3. Kane JM, Leucht S, Carpenter D, Docherty JP. Expert consensus guideline series. Optimizing pharmacologic treatment of psychotic disorders. Introduction: methods, commentary, and summary. In: The Journal of Clinical Psychiatry. Vol 64 Suppl 1. ; 2003:5–19.

4. Kreyenbuhl J, Buchanan RW, Dickerson FB, Dixon LB. The schizophrenia patient outcomes research team (PORT): Updated treatment recommendations 2009. Schizophr Bull. 2010;36(1):94–103. doi:10.1093/schbul/sbp130

5. Luykx JJ, Stam N, Tanskanen A, Tiihonen J, Taipale H. In the aftermath of clozapine discontinuation: comparative effectiveness and safety of antipsychotics in patients with schizophrenia who discontinue clozapine. Br J Psychiatry. Published online January 2020:1-8. doi:10.1192/bjp.2019.267

6. Miyamoto S, Miyake N, Jarskog LF, Fleischhacker WW, Lieberman JA. Pharmacological treatment of schizophrenia: A critical review of the pharmacology and clinical effects of current and future therapeutic agents. Mol Psychiatry. Published online 2012. doi:10.1038/mp.2012.47

7. Siskind D, Siskind V, Kisely S. Clozapine Response Rates among People with Treatment-Resistant Schizophrenia: Data from a Systematic Review and Meta-Analysis. Can J Psychiatry. 2017;62(11):772–777. doi:10.1177/0706743717718167

8. Taylor DM, Young C, Paton C. Prior antipsychotic prescribing in patients currently receiving clozapine: A case note review. J Clin Psychiatry. 2003;64(1):30–34. doi:10.4088/JCP.v64n0107

9. Doyle R, Behan C, OLJKeeffe D, et al. Clozapine Use in a Cohort of First-Episode Psychosis. J Clin Psychopharmacol. Published online 2017:1. doi:10.1097/JCP.0000000000000734

10. Howes OD, Vergunst F, Gee S, McGuire P, Kapur S, Taylor D. Adherence to treatment guidelines in clinical practice: Study of antipsychotic treatment prior to clozapine initiation. Br J Psychiatry. 2012;201(6):481–485. doi:10.1192/bjp.bp.111.105833

11. Shah P, Iwata Y, Plitman E, et al. The impact of delay in clozapine initiation on treatment outcomes in patients with treatment-resistant schizophrenia: A systematic review. Psychiatry Res. Published online 2018. doi:10.1016/j.psychres.2018.06.070

12. John AP, Ko EKF, Dominic A. Delayed Initiation of Clozapine Continues to Be a Substantial Clinical Concern. Can J Psychiatry. 2018;63(8):526–531. doi:10.1177/0706743718772522

13. Jin H, Tappenden P, MacCabe JH, Robinson S, McCrone P, Byford S. Cost and health impacts of adherence to the National Institute for Health and Care Excellence schizophrenia guideline recommendations. Br J Psychiatry. Published online December 2020:1-6. doi:10.1192/bjp.2020.241

14. Vermeulen JM, van Rooijen G, van de Kerkhof MPJ, Sutterland AL, Correll CU, de Haan L. Clozapine and Long-Term Mortality Risk in Patients With Schizophrenia: A Systematic Review and Meta-analysis of Studies Lasting 1.1-12.5 Years. Schizophr Bull. 2019;45(2):315–329. doi:10.1093/schbul/sby052

15. Okhuijsen-Pfeifer C, Huijsman EAH, Hasan A, et al. Clozapine as a first- or second-line treatment in schizophrenia: a systematic review and meta-analysis. Acta Psychiatr Scand. 2018;138(4):281–288. doi:10.1111/acps.12954

16. Siskind D, McCartney L, Goldschlager R, Kisely S. Clozapine v. first-and second-generation antipsychotics in treatment-refractory schizophrenia: systematic review and meta-analysis. Br J Psychiatry. 2016;209(5):385–392. doi:10.1192/bjp.bp.115.177261

17. Smith RL, O’Connell K, Athanasiu L, et al. Identification of a novel polymorphism associated with reduced clozapine concentration in schizophrenia patients-a genome-wide association study adjusting for smoking habits. Transl Psychiatry. 2020;10(1):198. doi:10.1038/s41398-020-00888-1

18. Pardiñas AF, Nalmpanti M, Pocklington AJ, et al. Pharmacogenomic Variants and Drug Interactions Identified Through the Genetic Analysis of Clozapine Metabolism. Am J Psychiatry. 2019;176(6):477–486. doi:10.1176/appi.ajp.2019.18050589

19. de With SAJ, Pulit SL, Staal WG, Kahn RS, Ophoff RA. More than 25 years of genetic studies of clozapine-induced agranulocytosis. Pharmacogenomics J. 2017;17(4):304–311. doi:10.1038/tpj.2017.6

20. Legge SE, Hamshere ML, Ripke S, et al. Genome-wide common and rare variant analysis provides novel insights into clozapine-associated neutropenia. Mol Psychiatry. 2018;23(1):162–163. doi:10.1038/mp.2017.214

21. Legge SE, Pardiñas AF, Helthuis M, et al. A genome-wide association study in individuals of African ancestry reveals the importance of the Duffy-null genotype in the assessment of clozapine-related neutropenia. Mol Psychiatry. 2019;24(3):328–337. doi:10.1038/s41380-018-0335-7

22. Frank J, Lang M, Witt SH, et al. Identification of increased genetic risk scores for schizophrenia in treatment-resistant patients. Mol Psychiatry. Published online 2015. doi:10.1038/mp.2014.56

23. Li J, Yoshikawa A, Brennan MD, Ramsey TL, Meltzer HY. Genetic predictors of antipsychotic response to lurasidone identified in a genome wide association study and by schizophrenia risk genes. Schizophr Res. 2018;192:194–204. doi:10.1016/j.schres.2017.04.009

24. Santoro ML, Ota V, de Jong S, et al. Polygenic risk score analyses of symptoms and treatment response in an antipsychotic-naive first episode of psychosis cohort. Transl Psychiatry. 2018;8(1):174. doi:10.1038/s41398-018-0230-7

25. Amare AT, Schubert KO, Hou L, et al. Association of Polygenic Score for Schizophrenia and HLA Antigen and Inflammation Genes With Response to Lithium in Bipolar Affective Disorder: A Genome-Wide Association Study. JAMA psychiatry. 2018;75(1):65–74. doi:10.1001/jamapsychiatry.2017.3433

26. Zhang J-P, Robinson D, Yu J, et al. Schizophrenia Polygenic Risk Score as a Predictor of Antipsychotic Efficacy in First-Episode Psychosis. Am J Psychiatry. 2019;176(1):21–28. doi:10.1176/appi.ajp.2018.17121363

27. Thorn CF, Müller DJ, Altman RB, Klein TE. PharmGKB summary: clozapine pathway, pharmacokinetics. Pharmacogenet Genomics. 2018;28(9):214–222. doi:10.1097/FPC.0000000000000347

28. Wagner E, Kane JM, Correll CU, et al. Clozapine Combination and Augmentation Strategies in Patients With Schizophrenia-Recommendations From an International Expert Survey Among the Treatment Response and Resistance in Psychosis (TRRIP) Working Group. Schizophr Bull. Published online May 2020. doi:10.1093/schbul/sbaa060

29. Jaquenoud Sirot E, Knezevic B, Morena GP, et al. ABCB1 and cytochrome P450 polymorphisms: clinical pharmacogenetics of clozapine. J Clin Psychopharmacol. 2009;29(4):319–326. doi:10.1097/JCP.0b013e3181acc372

30. Brennan MD. Pharmacogenetics of second-generation antipsychotics. Pharmacogenomics. 2014;15(6):869–884. doi:10.2217/pgs.14.50

31. Lee S-T, Ryu S, Kim S-R, et al. Association study of 27 annotated genes for clozapine pharmacogenetics: validation of preexisting studies and identification of a new candidate gene, ABCB1, for treatment response. J Clin Psychopharmacol. 2012;32(4):441–448. doi:10.1097/JCP.0b013e31825ac35c

32. Pouget JG, Shams TA, Tiwari AK, Müller DJ. Pharmacogenetics and outcome with antipsychotic drugs. Dialogues Clin Neurosci. 2014;16(4):555–566.

33. Lesche D, Mostafa S, Everall I, Pantelis C, Bousman CA. Impact of CYP1A2, CYP2C19, and CYP2D6 genotype-and phenoconversion-predicted enzyme activity on clozapine exposure and symptom severity. Pharmacogenomics J. 2020;20(2):192–201. doi:10.1038/s41397-019-0108-y

34. Wagner E, Oviedo-Salcedo T, Pelzer N, et al. Effects of Smoking Status on Remission and Metabolic and Cognitive Outcomes in Schizophrenia Patients Treated with Clozapine. Pharmacopsychiatry. Published online 2020. doi:10.1055/a-1208-0045

35. Okhuijsen-Pfeifer C, Ayhan Y, Lin BD, et al. Genetic Susceptibility to Clozapine-Induced Agranulocytosis/Neutropenia Across Ethnicities: Results From a New Cohort of Turkish and Other Caucasian Participants, and Meta-Analysis. Schizophr Bull Open. Published online 2020. doi:10.1093/schizbullopen/sgaa024

36. Okhuijsen-Pfeifer C, Cohen D, Bogers JPAM, et al. Differences between physicians’ and nurse practitioners’ viewpoints on reasons for clozapine underprescription. Brain Behav. Published online 2019. doi:10.1002/brb3.1318

37. Huisman, R., Okhuijsen-Pfeifer, C., Mulder, E.Y.H., Jongkind, A., Cohen, D., Bogers, J.P.A.M., Van der horst, M.Z., Luykx J. [Validatie van de Nederlandstalige Glasgow Antipsychotica Bijwerkingen Schaal voor Clozapine]. Tijdschr Psychiatr. Published online 2020.

38. World Medical Association. Declaration of Helsinki. Ethical Principles for Medical Research Involving Human Subjects. 64th WMA General Assembly, Fortaleza, Brazil, October 2013.; 2013.

39. Kay SR, Fiszbein A, Opler LA. The positive and negative syndrome scale (PANSS) for schizophrenia. Schizophr Bull. Published online 1987. doi:10.1093/schbul/13.2.261

40. National Institute of Mental Health. CGI. Clinical Global Impressions. ECDEU Assess Man Psychopharmacol Revis. Published online 1976.

41. Luykx JJ, Bakker SC, Visser WF, et al. Genome-wide association study of NMDA receptor coagonists in human cerebrospinal fluid and plasma. Mol Psychiatry. 2015;20(12):1557–1564. doi:10.1038/mp.2014.190

42. Luykx JJ, Bakker SC, Lentjes E, et al. Genome-wide association study of monoamine metabolite levels in human cerebrospinal fluid. Mol Psychiatry. 2014;19(2):228–234. doi:10.1038/mp.2012.183

43. Nievergelt CM, Maihofer AX, Klengel T, et al. International meta-analysis of PTSD genome-wide association studies identifies sex-and ancestry-specific genetic risk loci. Nat Commun. 2019;10(1):4558. doi:10.1038/s41467-019-12576-w

44. Watanabe K, Taskesen E, van Bochoven A, Posthuma D. Functional mapping and annotation of genetic associations with FUMA. Nat Commun. 2017;8(1):1826. doi:10.1038/s41467-017-01261-5

45. Sey NYA, Hu B, Mah W, et al. A computational tool (H-MAGMA) for improved prediction of brain-disorder risk genes by incorporating brain chromatin interaction profiles. Nat Neurosci. 2020;23(4):583–593. doi:10.1038/s41593-020-0603-0

46. Wray NR, Goddard ME, Visscher PM. Prediction of individual genetic risk to disease from genome-wide association studies. Genome Res. 2007;17(10):1520–1528. doi:10.1101/gr.6665407

47. Pardiñas AF, Holmans P, Pocklington AJ, et al. Common schizophrenia alleles are enriched in mutation-intolerant genes and in regions under strong background selection. Nat Genet. 2018;50(3):381–389. doi:10.1038/s41588-018-0059-2

48. Genomic Relationships, Novel Loci, and Pleiotropic Mechanisms across Eight Psychiatric Disorders. Cell. 2019;179(7):1469-1482.e11. doi:10.1016/j.cell.2019.11.020

49. Li Z, Huang M, Ichikawa J, Dai J, Meltzer HY. N-desmethylclozapine, a major metabolite of clozapine, increases cortical acetylcholine and dopamine release in vivo via stimulation of M1 muscarinic receptors. Neuropsychopharmacology. Published online 2005. doi:10.1038/sj.npp.1300768

50. Sur C, Mallorga PJ, Wittmann M, et al. N-desmethylclozapine, an allosteric agonist at muscarinic 1 receptor, potentiates N-methyl-D-aspartate receptor activity. Proc Natl Acad Sci U S A. Published online 2003. doi:10.1073/pnas.1835612100

51. Lee S-B, Wheeler MM, Thummel KE, Nickerson DA. Calling Star Alleles With Stargazer in 28 Pharmacogenes With Whole Genome Sequences. Clin Pharmacol Ther. 2019;106(6):1328–1337. doi:10.1002/cpt.1552

52. Gaedigk A, Ingelman-Sundberg M, Miller NA, Leeder JS, Whirl-Carrillo M, Klein TE. The Pharmacogene Variation (PharmVar) Consortium: Incorporation of the Human Cytochrome P450 (CYP) Allele Nomenclature Database. Clin Pharmacol Ther. 2018;103(3):399–401. doi:10.1002/cpt.910

53. Mrazek DA, Biernacka JM, O’Kane DJ, et al. CYP2C19 variation and citalopram response. Pharmacogenet Genomics. 2011;21(1):1–9. doi:10.1097/fpc.0b013e328340bc5a

54. Saiz-Rodríguez M, Ochoa D, Belmonte C, et al. Polymorphisms in CYP1A2, CYP2C9 and ABCB1 affect agomelatine pharmacokinetics. J Psychopharmacol. 2019;33(4):522–531. doi:10.1177/0269881119827959

55. Whirl-Carrillo M, McDonagh EM, Hebert JM, et al. Pharmacogenomics knowledge for personalized medicine. Clin Pharmacol Ther. 2012;92(4):414–417. doi:10.1038/clpt.2012.96

56. Steele-Perkins G, Plachez C, Butz KG, et al. The transcription factor gene Nfib is essential for both lung maturation and brain development. Mol Cell Biol. 2005;25(2):685–698. doi:10.1128/MCB.25.2.685-698.2005

57. Uhl GR, Martinez MJ. PTPRD: neurobiology, genetics, and initial pharmacology of a pleiotropic contributor to brain phenotypes. Ann N Y Acad Sci. 2019;1451(1):112–129. doi:10.1111/nyas.14002

58. Liu X, Shimada T, Otowa T, et al. Genome-wide Association Study of Autism Spectrum Disorder in the East Asian Populations. Autism Res. 2016;9(3):340–349. doi:10.1002/aur.1536

59. Choucair N, Mignon-Ravix C, Cacciagli P, et al. Evidence that homozygous PTPRD gene microdeletion causes trigonocephaly, hearing loss, and intellectual disability. Mol Cytogenet. 2015;8:39. doi:10.1186/s13039-015-0149-0

60. Revealing the complex genetic architecture of obsessive-compulsive disorder using meta-analysis. Mol Psychiatry. 2018;23(5):1181–1188. doi:10.1038/mp.2017.154

61. Uhl GR, Martinez MJ, Paik P, et al. Cocaine reward is reduced by decreased expression of receptor-type protein tyrosine phosphatase D (PTPRD) and by a novel PTPRD antagonist. Proc Natl Acad Sci U S A. 2018;115(45):11597–11602. doi:10.1073/pnas.1720446115

62. Cox JW, Sherva RM, Lunetta KL, et al. Genome-Wide Association Study of Opioid Cessation. J Clin Med. 2020;9(1). doi:10.3390/jcm9010180

63. Jonas KG, Lencz T, Li K, et al. Schizophrenia polygenic risk score and 20-year course of illness in psychotic disorders. Transl Psychiatry. 2019;9(1):300. doi:10.1038/s41398-019-0612-5

64. Werner MCF, Wirgenes KV, Haram M, et al. Indicated association between polygenic risk score and treatment-resistance in a naturalistic sample of patients with schizophrenia spectrum disorders. Schizophr Res. 2020;218:55–62. doi:10.1016/j.schres.2020.03.006

65. Vassos E, Di Forti M, Coleman J, et al. An Examination of Polygenic Score Risk Prediction in Individuals With First-Episode Psychosis. Biol Psychiatry. 2017;81(6):470–477. doi:10.1016/j.biopsych.2016.06.028

66. Wimberley T, Gasse C, Meier SM, Agerbo E, MacCabe JH, Horsdal HT. Polygenic risk score for schizophrenia and treatment-resistant schizophrenia. Schizophr Bull. Published online 2017. doi:10.1093/schbul/sbx007

67. Legge SE, Dennison CA, Pardiñas AF, et al. Clinical indicators of treatment-resistant psychosis. Br J Psychiatry. Published online 2020. doi:10.1192/bjp.2019.120

68. Piatkov I, Caetano D, Assur Y, et al. CYP2C19*17 protects against metabolic complications of clozapine treatment. world J Biol psychiatry Off J World Fed Soc Biol Psychiatry. 2017;18(7):521–527. doi:10.1080/15622975.2017.1347712

69. van de Bilt MT, Prado CM, Ojopi EPB, et al. Cytochrome P450 genotypes are not associated with refractoriness to antipsychotic treatment. Schizophr Res. 2015;168(1-2):587–588. doi:10.1016/j.schres.2015.08.002

70. Rodrigues-Silva C, Semedo AT, Neri HF da S, et al. The CYP2C19*2 and CYP2C19*17 Polymorphisms Influence Responses to Clozapine for the Treatment of Schizophrenia. Neuropsychiatr Dis Treat. 2020;16:427–432. doi:10.2147/NDT.S228103

71. Tóth K, Csukly G, Sirok D, et al. Potential Role of Patients’ CYP3A-Status in Clozapine Pharmacokinetics. Int J Neuropsychopharmacol. 2017;20(7):529–537. doi:10.1093/ijnp/pyx019

72. Kirchheiner J, Nickchen K, Bauer M, et al. Pharmacogenetics of antidepressants and antipsychotics: the contribution of allelic variations to the phenotype of drug response. Mol Psychiatry. 2004;9(5):442–473. doi:10.1038/sj.mp.4001494

73. Hedlund E, Gustafsson JA, Warner M. Cytochrome P450 in the brain; a review. Curr Drug Metab. 2001;2(3):245–263. doi:10.2174/1389200013338513

74. Ingelman-Sundberg M, Persson A, Jukic MM. Polymorphic expression of CYP2C19 and CYP2D6 in the developing and adult human brain causing variability in cognition, risk for depression and suicide: the search for the endogenous substrates. Pharmacogenomics. 2014;15(15):1841–1844. doi:10.2217/pgs.14.151

75. Savadlou A, Arnatkeviciute A, Tiego J, et al. Impact of CYP2C19 genotype-predicted enzyme activity on hippocampal volume, anxiety, and depression. Psychiatry Res. 2020;288:112984. doi:10.1016/j.psychres.2020.112984

