## Supplements for "Genome-wide association analyses of symptom severity among clozapine-treated patients with schizophrenia spectrum disorder"

Supplementary Information**

**Supplementary Methods**…………………………………………………………………………..……….……..3

**Supplementary Table 1.** Conversion Table of PANSS Scores and CGI-S Scores…………………...……….….7

**Supplementary Figure 1.** Principal Components Plots for Primary and Sensitivity Analyses……..………….....8

**Supplementary Table 2.** Number of Samples and SNPs Included in Each Step of the Quality Control……..…..9

**Supplementary Table 3.** Complex-LD Regions and Long-Range LD Regions that Were Excluded from PRS-Analyses……………………………………………………………………………..……………….…………...10

**Supplementary Table 4.** Activity Scoring Assignments for Each of the Observed Alleles……………..……...11

**Supplementary Table 5.** List of Inhibitors and Inducers by Enzyme…………………………….…..……..…..13

**Supplementary Table 6.** Descriptive Statistics of the Study Population………………………….…..………...14

**Supplementary Figure 2.** Q-Q plots of the Genome-wide Association Analyses…………………………...….15

**Supplementary Figure 3.** Regional Associations Plots for the Top SNPs identified in GWA Analysis………..16

**Supplementary Table 7.** Top SNPs Identified in the GWA Analyses…………………………………………..17

**Supplementary Results**………………………………………………………………………………………….18

**Supplementary Figure 4.** Dose-corrected clozapine concentrations in relation to the rs1923778 polymorphisms……………………………………………………………………………………………………19

**Supplementary Table 8.** Tukey post-hoc test for dose-adjusted clozapine concentrations of the rs1923778 polymorphism……………………………………………………………………………………………………..19

**Supplementary Figure 5.** Tissue specificity results for quantitative outcome and binary outcome…………….20

**Supplementary Figure 6.** Gene-based test as computed by MAGMA based on our genome-wide association analysis summary statistics……………………………………………………………………………………….22

**Supplementary Table 9.** Positive Predicted Values and Odds Ratios for Binary Outcome and Schizophrenia-PRS, based on various cut-offs……………………………………………………………….…………………..23

**Supplementary Table 10.** Positive Predicted Values and Odds Ratios for Binary Outcome and Schizophrenia-PRS, top vs tail tertile and decile..……………………………………………………………………….……….24

**Supplementary Figure 7.** Bar Plots Illustrating the Explained Variance of the Association between PRS and Symptom Severity………………………………………………………………………………………………...25

**Supplementary Table 11.** Logistic Regression Model for Genotype-Predictive Enzyme Activity Scores of Binary Outcome. …………………………………………………………………………………………………26

**Supplementary Table 12.** Linear Regression Model for Genotype-Predictive Enzyme Activity Scores of Quantitative Outcome. …………………………………………………………………………………………...27

**Supplementary Table 13.** Linear Regression Model for Genotype-Predictive Enzyme Activity Scores of Dose-Adjusted Clozapine Levels. …………………………………………………………………………………...…28

**Supplementary Table 14.** Combined Pharmacogenetic and PRS logistic Regression Model of Binary
Outcome. …………………………………………………………………………………………………………29

**Supplementary Table 15.** Combined Pharmacogenetic and PRS with PCs Logistic Regression Model of
Binary Outcome. …………………………………………………………………………………………………30

**Supplementary Table 16.** Combined Pharmacogenetic and Top GWA Hits with PCs Logistic Regression Model of Binary Outcome. ………………………………………………………………………………………32

**Supplementary Table 17**. Descriptive statistics on dose-adjusted clozapine concentrations and high vs. low symptom severity. Abbreviations: N=number of participants, SD=standard deviation, SE=standard error……..34

**Supplementary Table 18**. Results of an Independent Samples T-Test between dose-adjusted clozapine concentrations and high vs. low symptom severity. Abbreviations: df=degrees of freedom, p=*p*-value….……..34

**Supplementary Table 19.** Linkage Disequilibrium Statistics of Our Two Top Hits and Top Hits of Previous Performed Studies. ……………………………………………………………………………………………….34

**Supplementary Figure 8.** Dose-adjusted clozapine concentrations between cohorts…………………………...35

**Supplementary Figure 9.** Bar Plots Illustrating the Explained Variance of PRS and Symptom Severity, with Exclusion of the GROUP Cohort. ………………………………………………………………………………..36

**Supplementary Figure 10.** Heatmap plots using 54 tissues from GTEx for quantitative outcome and binary outcome. ………………………………………………………………………………………………………….37

**Supplementary Figure 11.** Gene-based association results from H-MAGMA using Hi-C interaction for all outcomes. …………………………………………………………………………………………………………38

**Supplementary Figure 12.** Gene ontologies enriched for symptom severity linked to genes for each
outcome.. …………………………………………………………………………………………………………40

**Supplementary Figure 13**. Visual plots of the mean and median of dose-adjusted clozapine concentrations and high vs. low symptom severity , and spread of dose-adjusted clozapine concentrations and Clinical Global impression (CGI) severity question. ……………………………………………………………………………...42

**Supplementary Figure 14.** Box Plots Illustrating the Association between Smoking Status and Dose-Adjusted Clozapine Concentrations and CGI-S Score. …………………………………………………………………….43

**Supplementary Acknowledgements**…………………………………………………………………………….44

**References**………………………………………………………………………………………………………..45

**Supplementary Methods**

**Detailed recruitment methods per cohort**

*Clozapine International (CLOZIN) consortium and Mental Health Services Rivierduinen*

The CLOZIN consortium recruited inpatient and outpatient participants in the Netherlands, Germany, Austria, and Finland. Mental Health Services Rivierduinen recruited participants in The Netherlands and followed the same methods and procedures as CLOZIN. Participants were enrolled when they were diagnosed by their treating physician with a schizophrenia spectrum disorder according to Diagnostic and Statistical Manual of Mental Disorders, Fourth or Fifth Edition (DSM-IV-TR or DSM-5) and were currently using clozapine or did use clozapine in the past. All participants were aged 18 years or older, were able to speak and read the local language, and were able and willing to provide written informed consent. Blood was collected for DNA extraction and the Clinical Global Impression (CGI)^1^ scale was obtained, to evaluate symptom severity and treatment response. Blood samples for measurement of clozapine plasma concentrations in clinical care were collected ~12h after the last clozapine dose intake. Clozapine plasma concentrations were measured using liquid chromatograph tandem mass spectrometry (LC-MS/MS) in a local accredited laboratory. Recruitment for all centers was approved by their respective local Institutional Review Boards.

*Cooperative Research Centre (CRC) Cohort Recruitment*

Schizophrenia patients were recruited from inpatient and outpatient clinics around Melbourne, Australia. The Mini International Neuropsychiatric Interview (MINI)^2^ was used to confirm the primary diagnosis from the treating physician. Only patients with a confirmed diagnosis were included in the current analyses. All participants were aged between 18-65 years and were currently prescribed and taking clozapine. They were considered as ‘treatment-resistant’ as they failed to respond to two or more previous trials of antipsychotics with persistent symptoms and poor functioning.^3^ After an overnight fast, whole blood was collected and processed. Blood samples for DNA extraction and measurement of clozapine plasma concentrations were collected after overnight fasting (~12h after last clozapine dose intake). Clozapine plasma concentrations were measured using a liquid chromatograph tandem mass spectrometry (LC-MS/MS) method in a National Association of Testing Authorities, Australia accredited laboratory. Participants were provided with written information and written consent was sought from all eligible individuals prior to participation. This recruitment was approved by the Melbourne Health Human Research Ethics Committee (MHREC ID 2012.069).

*Genetic Risk and Outcome of Psychosis (GROUP) consortium*

The GROUP study was conducted by a consortium of four university psychiatric centers based in the Netherlands, and in total, thirty-six mental health care institutes participated in the study. Amongst other measurements, blood was collected for DNA extraction and the Positive and Negative Syndrome Scale (PANSS)^4^ was done to measure severity of a variety of symptoms. This project was specifically aimed at relatively young participants to allow for long-term follow-up. Patients were included if they were aged 16 to 50 years; diagnosed by their treating physician with a psychotic disorder according to DSM-IV-TR, had good command of the Dutch language, and were able and willing to give written informed consent. The study protocol was approved centrally by the Ethical Review Board of the University Medical Centre Utrecht and subsequently by local review boards of each participating institute.

*Hacettepe University*

The recruitment of patients at the Turkish site was previously described.^5^ In short, patients were recruited by Hacettepe University (HU), in collaboration with the members of the Schizophrenia and Other Psychotic Disorders Section of the Psychiatric Association of Turkey, working in various psychiatry clinics. Patients were included if they were aged 18 to 65 years, were diagnosed by their treating physician with schizophrenia or schizoaffective disorder according to DSM-IV-TR, and were using clozapine for at least 10 years or had developed clozapine-induced agranulocytosis (10 patients). Samples from HU were used since detailed clinical information and clozapine levels were available only for HU as a part of a research database at the time of the study.^6^ If previous clinical assessments were not readily accessible, a new assessment was conducted using clinical information from patient records. Blood samples for measurement of clozapine serum concentrations in clinical care were collected ~12h after the last clozapine dose intake. Clozapine serum concentrations were measured using using high-performance-liquid-chromatography coupled with ultraviolet detection (the Shimadzu Prominence device of Rotakim Analysis Services and Technical Equipment Company). The study protocol was approved by the Hacettepe University Faculty of Medicine Research Ethics Committee on March 24, 2011 (Project Number: FON 11/21). Only subjects who provided written informed consent were included in the study.

**Phenotypes**
Symptom severity was assessed by treating physicians or trained study raters using the Positive and Negative Syndrome Scale (PANSS) and/or the Clinical Global Impression-Severity (CGI-S) scale. PANSS is a semi-structured interview that assesses positive and negative symptoms, and general psychopathology, whereas the CGI-S is a single item assessment of the severity of disease on a 7-point Likert scale.^1,4^ CGI-S scores were available for participants from CLOZIN, Hacettepe University, and Mental Health Services Rivierduinen, and PANSS scores for participants from GROUP and CRC Australia. Previous studies have shown a correlation between PANSS scores and Brief Psychiatric Rating Scale (BPRS) scores, and between CGI and BPRS scores. ^7,8^ The authors of these studies were involved in the design of the current study and provided a table from their original research to convert PANSS scores to CGI scores and vice versa (Supplementary Table 1). Our main outcome measure was therefore continuous, allowing for linear regression analyses. For symptom severity defined as a binary outcome (hereafter referred to as ‘binary outcome’), we divided the participants into ‘low’ and ‘high’ symptom severity. Low symptom severity corresponded to a CGI-S score of 1 to 3 (‘normal’ to ‘mildly ill’) and high symptom severity corresponded to a CGI-S score of 4 to 7 (‘moderately ill’ to ‘among the most extremely ill patients’). ^1^ For participants for whom no CGI-S score was available, PANSS scores were converted to CGI-S scores using the aforementioned conversion table. Consequently, a PANSS score of 30 to 74 corresponded to low symptom severity, and a PANSS score ≥ 75 to high symptom severity. To define symptom severity as a quantitative phenotype (hereafter referred to as ‘quantitative outcome’), we used the CGI-S score. We chose CGI-S scores over PANSS scores because two-third of the participants were assessed using the CGI-S, it assesses both symptoms and global functioning, and it has high validity and is easy to use in clinical practice, resulting in higher potential for clinical translation of possible findings. ^9,10^

**Genotyping and Quality Control**

All participants were genotyped in a single batch on the most recent Illumina Infinium® Global Screening Array, version 3 (Illumina, San Diego, CA, USA). Genotyping was done in the Human Genotyping Facility of Erasmus Medical Center (Erasmus MC) Rotterdam. This platform contains a total number of 730,059 genetic markers. Samples were assigned randomly to plates to avoid cohort-specific batch effects. Approximately 200 ng of genomic DNA was used to genotype each sample. Calling was performed using Illumina Genomestudio software, without any filtering presets. We applied standard participant and single nucleotide polymorphism (SNP) level quality control (QC) using PLINK v1.90b3z 64-bit (22 Nov 2015; https://www.cog-genomics.org/plink2version) to ensure inclusion of only well performing SNPs in well genotyped individuals. ^11,12^ We first removed individuals with a genotype call rate<0.95. We selected a set of high-quality single nucleotide polymorphisms (SNPs) for heterozygosity, relatedness, and principal components (PCs) analysis by including SNPs with a call rate>0.99, a minor allele frequency (MAF)>0.1, a Hardy-Weinberg Equilibrium (HWE) *p*-value (*p*)*>*1x10^-4^, and Linkage Disequilibrium (LD)-pruned with an r²>0.2, a window size of 50, and window shifting per 5 SNPs. We removed individuals with discordance between genetic sex and reported sex in phenotype data and an excess heterozygosity or homozygosity rate (> 3 standard deviations from the mean). Additionally, we identified pairs of individuals with a relatedness coefficient (PLINK PIHAT)>0.1 and removed the individuals with the lowest calling rate. In addition, we calculated the first 10 PCs in PLINK. Outliers were identified by visual inspection and compared with their reported ethnicity (Supplementary Figure 1). In case of discrepancies, these samples were removed (Supplementary Table 2). Next, good quality SNPs were selected by removing non-autosomal genetic markers, SNPs with a call rate<0.95, SNPs out of HWE p<1×10^−6^, and SNPs with a strand-ambiguous A/T or C/G.

After this extensive QC, pre-imputation checks were done based on the reference panel that was used for imputation, namely the Haplotype Reference Consortium (HRC) release HRC r1.1 2016 (GRCh37/hg19)).^13^ SNPs that were not present in the reference panel or had a difference in MAF>0.15 compared to this reference panel, were removed. ^14^ Finally, the SNPs were imputed on the Michigan Imputation Server (https://imputationserver.sph.umich.edu) using Minimac4 after which additional post-imputation QC on the imputed data was performed: SNPs with a MAF<0.05 or an imputation score (R2)<0.3 were removed.^14^

**Gene set enrichment analysis using FUMA**

We used Functional Mapping and Annotation (FUMA) of genome-wide association studies (GWASs) to obtain gene-level summary statistics from our binary and quantitative GWAS summary statistics to perform extensive functional annotation, such as gene-set and cell/tissue type enrichment analysis.^15,16^ Then the gene-level summary statistics were used to create a differential gene expression analysis (DEG) (GTEx v8 and BrainSpan data) and expression heatmaps.

**H-MAGMA**

To identify genes associated with symptom severity, we applied Hi-C coupled Multi-marker Analysis of GenoMic Annotation (H-MAGMA), which aggregates SNP-level *p*-values into a gene-level association statistic with an additional assignment of non-coding SNPs to their chromatin-interacting target genes generated from fetal and adult brain Hi-C.

We thus conducted H-MAGMA analysis which predicts genes associated with the target phenotype by integrating long-range chromatin interaction with GWAS’ summary statistics.^17^ Together with existing eQTL resources in the adult and fetal cortex, it is possible to link variants associated with risk for symptom severity to target genes and functional pathways. More specifically, SNP to Ensemble gene annotation was carried out in H-MAGMA (https://github.com/thewonlab/H-MAGMA) by leveraging chromatin-interaction data generated from adult brain^18^ and fetal brain^19^ Hi-C. We used Gencode v26 for assigning exonic SNPs and promoter SNPs (2kb upstream to the transcription start sites) to genes based on genomic location. Intronic and intergenic SNPs were mapped to their target genes based on chromatin interactions to promoters and exons generated by fetal brain and adult brain Hi-C. Using this gene-SNP relationship as input, we ran MAGMA (v1.0.8) to aggregate SNP-based *p*-values to gene-based *p*-values. We set *p*<0.005 as the significance threshold. H-MAGMA gene list was used for further gene ontology enrichment analysis.

Gene ontology enrichment analysis was performed using g:Profiler (v0.6.7)^20^ (https://biit.cs.ut.ee/gprofiler/ ) with the “ordered list” option in which all genes were ranked based on *p*-value from H-MAGMA gene-based test. We selected 19,222 protein-coding genes that were detected in the H-MAGMA gene list and not located within the MHC region (chr6:25M-35M) as the background. We tested enrichment within the Gene Ontology Molecular Functions (MF) and Biological Process (BP) categories.

**PRS analysis**

PRSs were constructed using various p-thresholds (pt): 5x10-8, 5x10-7, 5x10-6, 5x10-5, 5x10-4, 5x10-3, 0.05, 0.1, 0.2, 0.3, 0.4, and 0.5. Complex linkage disequilibrium (LD) regions and long-range LD regions were excluded from PRS analyses (Supplementary Table 3). The PRSs for the three traits were regressed against the two symptom severity phenotypes for both primary and sensitivity analyses, using linear regression for quantitative outcome and logistic regression for binary outcome in PRSice2, correcting for sex, age, and 10 PCs. ^21^ The Bonferroni-corrected significance level was p<0.017 as quantitative and binary outcomes were derived from the same variable. If only odds ratios (ORs) were reported as effect estimates in the summary statistics, they were log-converted to beta values. To that end, the beta values, effective allele, and *p*-values were extracted from all summary statistics. SNPs that overlapped between the summary statistics GWASs (training datasets), 1000 genomes, and our dataset (target) were extracted.^22^ Then, insertions or deletions, ambiguous SNPs, SNPs with MAF <0.01 and LD R2<0.8 in both training (if the information is available) and target datasets were excluded. We also excluded all SNPs in complex-LD regions and long-range LD regions in the genome (Supplementary Table 3). Clumping was performed with a cutoff of R2=0.1 using a 250-kb window.

**Genotype-predicted enzyme activity score calculation and analysis**

Genotype imputation was performed for chromosomes 10, 15, and 22, which include *CYP2C19, CYP1A2, CYP2D6*, respectively. Separate imputation was required because the imputation used for the GWAS analysis did not impute indels, which are required for calling *CYP2C19, CYP1A2, CYP2D6* haplotypes. The 1000 Genomes Phase 3 (version 5) reference panel (build hg19, population: mixed) was used to phase and impute SNPs as well as indels on the Michigan Imputation Server, with a post-imputation filter to select high quality calls (R2>0.5). Imputed data was subjected to Stargazer v1.08^23^ to call *CYP2C19, CYP1A2, CYP2D6* haplotypes (star alleles). Corresponding activity scores for *CYP2D6* were based on translation tables maintained by the Pharmacogene Variation (PharmVar) Consortium^24^ and the Pharmacogenomics Knowledgebase (PharmGKB)^25^ (Supplementary Table 4). Whereas, activity scores for *CYP1A2* and *CYP2C19* followed previous scoring methods.^26,27^ Importantly, the genome-wide association study (GWAS) platform that we used did not capture structural variations (including copy number variations). As a result, *CYP2D6* ultrarapid metabolizers could not be detected and were incorrectly classified as normal metabolizers. Based on the ancestry composition of the study participants, we estimate this misclassification would have occurred in 3% of our samples. However, to our knowledge, there is no evidence suggesting this misclassification error would have a meaningful impact on our study findings.

Prior to analysis, activity scores for *CYP2C19*, *CYP1A2*, and *CYP2D6* were corrected for concomitant inhibitors or inducers of each of the corresponding genes using established methodology.^28^ In the presence of a strong inhibitor the activity score was multiplied by zero, while in the presence of a moderate inhibitor the activity score was multiplied by 0.5. If an inducer was present the activity score was multiplied by 1.5 (see Supplementary Table 5 for a list of inhibitors and inducers). In cases where both an inhibitor and inducer of the same enzyme were present, the activity score for the corresponding gene was retained, as consensus on how to correct activity scores in this situation has not been established.**Supplementary Table 1**. Conversion table of Positive And Negative Syndrome Scale (PANSS) scores and Clinical Global Impression-Severity (CGI-S) scores, provided by prof. dr. Leucht and colleagues.^7^

| **PANSS** | **CGI** |  | **PANSS** | **CGI** |
| --- | --- | --- | --- | --- |
| 30 | 0,6 |  | 91 | 4,82 |
| 31 | 0,72 |  | 92 | 4,88 |
| 32 | 0,88 |  | 93 | 4,93 |
| 33 | 1,05 |  | 94 | 4,98 |
| 34 | 1,18 |  | 95 | 5,03 |
| 35 | 1,3 |  | 96 | 5,08 |
| 36 | 1,41 |  | 97 | 5,12 |
| 37 | 1,53 |  | 98 | 5,16 |
| 38 | 1,63 |  | 99 | 5,2 |
| 39 | 1,72 |  | 100 | 5,24 |
| 40 | 1,8 |  | 101 | 5,28 |
| 41 | 1,89 |  | 102 | 5,32 |
| 42 | 1,97 |  | 103 | 5,35 |
| 43 | 2,06 |  | 104 | 5,38 |
| 44 | 2,14 |  | 105 | 5,42 |
| 45 | 2,22 |  | 106 | 5,46 |
| 46 | 2,29 |  | 107 | 5,5 |
| 47 | 2,37 |  | 108 | 5,54 |
| 48 | 2,44 |  | 109 | 5,59 |
| 49 | 2,5 |  | 110 | 5,65 |
| 50 | 2,55 |  | 111 | 5,7 |
| 51 | 2,6 |  | 112 | 5,76 |
| 52 | 2,65 |  | 113 | 5,82 |
| 53 | 2,7 |  | 114 | 5,87 |
| 54 | 2,76 |  | 115 | 5,93 |
| 55 | 2,81 |  | 116 | 5,97 |
| 56 | 2,87 |  | 117 | 6,02 |
| 57 | 2,93 |  | 118 | 6,07 |
| 58 | 2,99 |  | 119 | 6,11 |
| 59 | 3,06 |  | 120 | 6,14 |
| 60 | 3,14 |  | 121 | 6,18 |
| 61 | 3,21 |  | 122 | 6,21 |
| 62 | 3,28 |  | 123 | 6,24 |
| 63 | 3,35 |  | 124 | 6,27 |
| 64 | 3,42 |  | 125 | 6,29 |
| 65 | 3,48 |  | 126 | 6,32 |
| 66 | 3,54 |  | 127 | 6,34 |
| 67 | 3,59 |  | 128 | 6,35 |
| 68 | 3,64 |  | 129 | 6,37 |
| 69 | 3,69 |  | 130 | 6,39 |
| 70 | 3,73 |  | 131 | 6,4 |
| 71 | 3,78 |  | 132 | 6,42 |
| 72 | 3,83 |  | 133 | 6,43 |
| 73 | 3,88 |  | 134 | 6,45 |
| 74 | 3,93 |  | 135 | 6,46 |
| 75 | 3,98 |  | 136 | 6,48 |
| 76 | 4,03 |  | 137 | 6,5 |
| 77 | 4,08 |  | 138 | 6,53 |
| 78 | 4,13 |  | 139 | 6,55 |
| 79 | 4,18 |  | 140 | 6,59 |
| 80 | 4,22 |  | 141 | 6,64 |
| 81 | 4,27 |  | 142 | 6,68 |
| 82 | 4,32 |  | 143 | 6,73 |
| 83 | 4,38 |  | 144 | 6,78 |
| 84 | 4,43 |  | 145 | 6,83 |
| 85 | 4,48 |  | 146 | 6,89 |
| 86 | 4,54 |  | 147 | 6,96 |
| 87 | 4,6 |  | 148 | 7,01 |
| 88 | 4,65 |  | 149 | 7,07 |
| 89 | 4,71 |  | 150 | 7,14 |
| 90 | 4,77 |  |  |  |

**Supplementary Figure 1.** Population structures identified by MDS (multidimensional scaling). The first and second (A1 & B1), and the third and fourth (A2 & B2) components are plotted against each other, for primary analysis (A), and sensitivity analysis (B). The HapMap3 population is shown in one color for overview purposes. The black dots, identified as ethnical outliers, were excluded from analyses.

**A1**
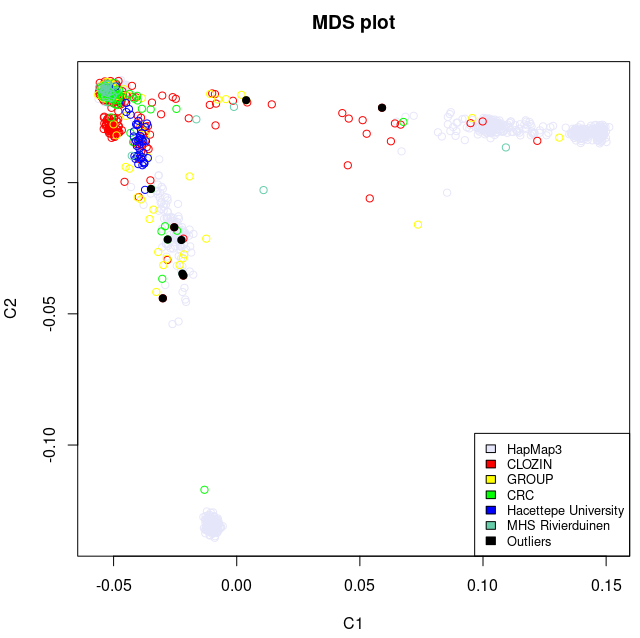
**A2**
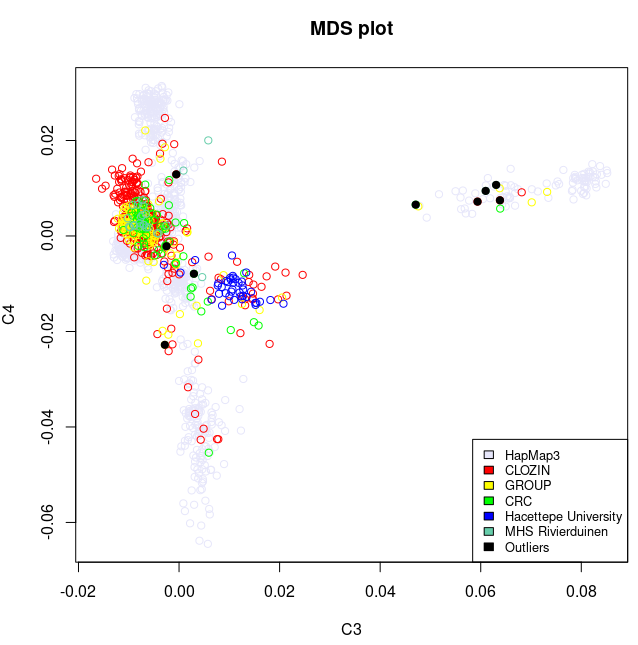

**B1**
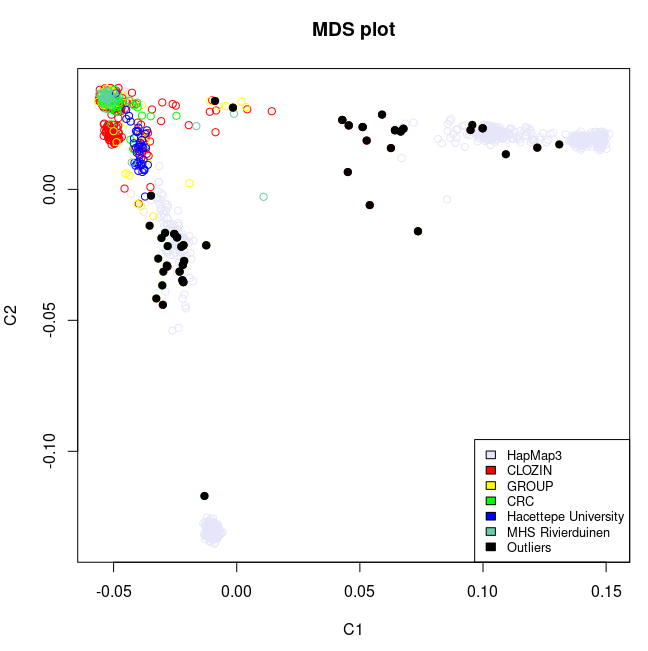
**B2**
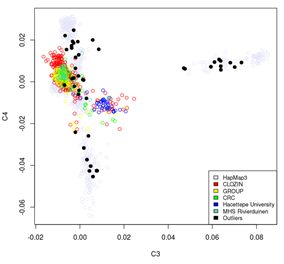

**Supplementary Table 2**. Number of samples and SNPs included in each step of the quality control.
*Abbreviation:* SNPs=Single Nucleotide Polymorphisms, HWE= Hardy-Weinberg Equilibrium, MAF=Minor Allele Frequency
*In the main text, N=804 is mentioned. This is the number of samples without duplicates.

| **Step** | **Number of participants** | **Number of SNPs** |
| --- | --- | --- |
| Start | 839* | 725,831 |
| Check missingness in SNPs and samples | 818 | 725,506 |
| Sexcheck | 799 | 725,831 |
| Pruning (get independent samples) genotype call rate>0.99, HWE>1x10^-4^, MAF> 0.1 | 799 | 238,143 |
| Check heterozygosity/homozygosity (for sample mix-up/contamination/inbreeding) | 776 | 131,722 |
| Exclude related samples & duplicates (PLINK PIHAT >0.1) | 734 | 131,722 |
| Remove ethnical outliers from dataset | 725 | 131,722 |
| Remove all failing samples and normal SNP quality control (removing non-autosomal genetic markers, SNPs with a call rate<0.95, SNPs out of HWE *p*<1×10^−6^, and SNPs with a strand-ambiguous A/T or C/G) | 725 | 688,618 |
| Remove samples with missing phenotype | 684 | 688,618 |
| Imputation with imputation score (R2)>0.3 | 684 | 5,506,411 |

**Supplementary Table 3.** 20 complex-LD (linkage disequilibrium) regions and long-range LD regions that were excluded from Polygenic Risk Score-analysis.^29^

| Chromosome | Base pair position (start point to end point) |
| --- | --- |
| 1 | 48000000-52000000 |
| 2 | 86000000-100500000 |
| 2 | 183000000-190000000 |
| 3 | 47500000-50000000 |
| 3 | 83500000-87000000 |
| 5 | 44500000-50500000 |
| 5 | 129000000-132000000 |
| 6 | 25500000-33500000 |
| 6 | 57000000-64000000 |
| 6 | 140000000-142500000 |
| 7 | 55000000-66000000 |
| 8 | 8000000-12000000 |
| 8 | 43000000-50000000 |
| 8 | 112000000-115000000 |
| 10 | 37000000-43000000 |
| 11 | 87500000-90500000 |
| 12 | 33000000-40000000 |
| 20 | 32000000-34500000 |
| 8 | 8135000-12000000 |
| 17 | 40900000-45000000 |

**Supplementary Table 4.** Activity scoring assignments for each of the observed alleles in the current study.
**^a^** Percentages may not equate to 100 due to rounding.

**^b^** Activity scores for *CYP2D6* were assigned based on the gene-specific information tables created by the Pharmacogenomics Knowledgebase (PharmGKB) and Clinical Pharmacogenetics Implementation Consortium (CPIC). Activity scores for CYP1A2 and CYP2C19 followed previous scoring methods.^26,27^

^c^ Predicted phenotypes for CYP1A2 and CYP2C19 are based on definitions provided by the Clinical Pharmacogenetics Implementation Consortium (CPIC). CYP2D6 phenotypes were based on the standardized activity score definitions developed jointly by CPIC and the Dutch Pharmacogenetics Working Group.

IM=intermediate metabolizer, NM=normal (extensive) metabolizer, PM=poor metabolizer, RM=rapid metabolizer, UM=ultrarapid metabolizer, IND=indeterminant.

| **Diplotype** | **Frequency** | | **Activity Score^b^** | **Predicted Phenotype^c^** |
| --- | --- | --- | --- | --- |
|  | **N** | **%^a^** |  |  |
| ***CYP1A2*** |  |  |  |  |
| ***1F/*1F** | 298 | 43.4 % | 3 | UM |
| ***1A/*1F** | 275 | 40.0 % | 2.5 | UM |
| ***1A/*1A** | 76 | 11.1 % | 2 | NM |
| ***1A/*1K** | 1 | 0.1 % | 1.5 | NM |
| ***1A/*1L** | 10 | 1.5 % | IND | IND |
| ***1F/*1L** | 24 | 3.5 % | IND | IND |
| ***1F/*1K** | 2 | 0.3 % | IND | IND |
| ***1L/*1L** | 1 | 0.1 % | IND | IND |
| ***CYP2D6*** |  |  |  |  |
| ***1/*1** | 104 | 15.1 % | 2 | NM |
| ***1/*2** | 107 | 15.6 % | 2 | NM |
| ***1/*33** | 4 | 0.6 % | 2 | NM |
| ***1/*35** | 35 | 5.1 % | 2 | NM |
| ***2/*2** | 21 | 3.1 % | 2 | NM |
| ***2 / *33** | 3 | 0.4 % | 2 | NM |
| ***2 / *35** | 12 | 1.7 % | 2 | NM |
| ***2 / *45** | 1 | 0.1 % | 2 | NM |
| ***33/*35** | 2 | 0.3 % | 2 | NM |
| ***35 / *35** | 4 | 0.6 % | 2 | NM |
| ***35 / *39** | 1 | 0.1 % | 2 | NM |
| ***1 / *9** | 11 | 1.6 % | 1.5 | NM |
| ***1/*41** | 45 | 6.6 % | 1.5 | NM |
| ***1/*17** | 3 | 0.4 % | 1.5 | NM |
| ***1 / *29** | 2 | 0.3 % | 1.5 | NM |
| ***2 / *9** | 4 | 0.6 % | 1.5 | NM |
| ***2/*17** | 1 | 0.1 % | 1.5 | NM |
| ***2 / *29** | 2 | 0.3 % | 1.5 | NM |
| ***2/*41** | 13 | 1.9 % | 1.5 | NM |
| ***2 / *59** | 2 | 0.3 % | 1.5 | NM |
| ***9 / *35** | 3 | 0.4 % | 1.5 | NM |
| ***33 / *41** | 2 | 0.3 % | 1.5 | NM |
| ***35 / *41** | 10 | 1.5 % | 1.5 | NM |
| ***1/*10** | 11 | 1.6 % | 1.5 | NM |
| ***1 / *59** | 1 | 0.1 % | 1.5 | NM |
| ***2/*10** | 5 | 0.7 % | 1.25 | NM |
| ***10 / *33** | 1 | 0.1 % | 1.25 | NM |
| ***10 / *35** | 1 | 0.1 % | 1.25 | NM |
| ***1/*3** | 11 | 1.6 % | 1 | IM |
| ***1/*4** | 95 | 13.8 % | 1 | IM |
| ***1/*6** | 4 | 0.6 % | 1 | IM |
| ***2 / *3** | 3 | 0.4 % | 1 | IM |
| ***2/*4** | 39 | 5.7 % | 1 | IM |
| ***2 / *6** | 5 | 0.7 % | 1 | IM |
| ***2/*31** | 1 | 0.1 % | 1 | IM |
| ***2 / *40** | 1 | 0.1 % | 1 | IM |
| ***3/*35** | 7 | 1.0 % | 1 | IM |
| ***4 / *33** | 2 | 0.3 % | 1 | IM |
| ***4 / *35** | 10 | 1.5 % | 1 | IM |
| ***9 / *41** | 2 | 0.3 % | 1 | IM |
| ***41 / *41** | 4 | 0.6 % | 1 | IM |
| ***9 / *10** | 1 | 0.1 % | 0.75 | IM |
| ***10 / *41** | 4 | 0.6 % | 0.75 | IM |
| ***4 / *9** | 6 | 0.9 % | 0.5 | IM |
| ***4 / *17** | 1 | 0.1 % | 0.5 | IM |
| ***4 / *29** | 1 | 0.1 % | 0.5 | IM |
| ***4/*41** | 30 | 4.4 % | 0.5 | IM |
| ***3 / *10** | 2 | 0.3 % | 0.25 | IM |
| ***4 / *10** | 11 | 1.6 % | 0.25 | IM |
| ***10 / *131** | 1 | 0.1 % | 0.25 | IM |
| ***4 / *131** | 1 | 0.1 % | 0 | PM |
| ***3/*3** | 1 | 0.1 % | 0 | PM |
| ***3/*4** | 4 | 0.6 % | 0 | PM |
| ***3 / *6** | 2 | 0.3 % | 0 | PM |
| ***4/*4** | 17 | 2.5 % | 0 | PM |
| ***4 / *6** | 2 | 0.3 % | 0 | PM |
| ***1 / *28** | 2 | 0.3 % | IND | IND |
| ***1 / *117** | 2 | 0.3 % | IND | IND |
| ***2 / *117** | 1 | 0.1 % | IND | IND |
| ***6 / *117** | 1 | 0.1 % | IND | IND |
| ***4 / *117** | 2 | 0.3 % | IND | IND |
| ***CYP2C19*** |  |  |  |  |
| ***17/*17** | 36 | 5.2 % | 3 | UM |
| ***1/*17** | 177 | 25.8 % | 2.5 | RM |
| ***1/*1** | 258 | 37.6 % | 2 | NM |
| ***2/*17** | 39 | 5.7 % | 1.5 | IM |
| ***8/*17** | 1 | 0.1 % | 1.5 | IM |
| ***1/*2** | 138 | 20.1 % | 1 | IM |
| ***1/*3** | 3 | 0.4 % | 1 | IM |
| ***1/*8** | 2 | 0.3 % | 1 | IM |
| ***1/*22** | 1 | 0.1 % | 1 | IM |
| ***1/*35** | 2 | 0.3 % | 1 | IM |
| ***2/*2** | 28 | 4.1 % | 0 | PM |
| ***2/*8** | 2 | 0.3 % | 0 | PM |

**Supplementary Table 5**. List of inhibitors and inducers by enzyme based on the Flockhart Table.^30^
m=moderate, s=strong.

|  | CYP1A2 | Frequency | CYP2C19 | Frequency | CYP2D6 | Frequency |
| --- | --- | --- | --- | --- | --- | --- |
| Inhibitors | fluvoxamine (s)  ciprofloxacin (s) | 7%  0% | fluoxetine (m)  esomeprazole (m)  ethinylestradiol (m)  voriconazole (m) | 2%  12%  1%  0% | fluoxetine (s)  bupropion (s)  paroxetine (s)  (es)citalopram (m)  levomepromazine (m)  sertraline (m)  duloxetine (m) | 2%  1%  1%  9%  1%  4%  0% |
| Inducers | smoking  carbamazepine  rifampcin  phenytoin  phenobarbital | 54%  0%  0%  0%  0% | carbamazepine  St john’s wort | 0%  0% |  |  |

**Supplementary Table 6.** Descriptive statistics of individuals included in the GWA and PRS analyses, after quality control.
*Abbreviations*: N=number of individuals, SD=Standard Deviation, Psychosis NOS=Psychosis Not Otherwise Specified, CLZ=clozapine, NA=Not Available.
CLOZIN consortium, the Netherlands, Germany, Austria, and Finland; GROUP consortium, the Netherlands; CRC: Cooperative Research Centre, Australia; Hacettepe University Ankara, Turkey; MHS: Mental Health Services Rivierduinen Leiden, The Netherlands. *Age data was available for all individuals. **See ‘Supplementary Methods - Detailed recruitment methods per cohort’ for explanation of the relatively young age in GROUP.

| Cohort | Total | CLOZIN | GROUP | CRC | Hacettepe University | MHS Rivierduinen |
| --- | --- | --- | --- | --- | --- | --- |
| N (% male) | 684 (73,0) | 407 (70,5) | 152 (83,4) | 67 (71,6) | 34 (44,1) | 24 (91,7) |
| Mean age  ± SD (years)* | 39,5 ±12,2 | 43,2 ± 11,9 | 27,6 ± 5,6** | 41,6 ± 9,8 | 42,5 ± 7,7 | 40,9 ± 12,6 |
| Diagnosis | Schizophrenia (N=552)  Schizoaffective disorder (N=95)  Schizophreniform disorder (N=1)  Psychosis NOS (N=36) | Schizophrenia (N=303)  Schizoaffective disorder (N=79)  Schizophreniform disorder (N=1)  Psychosis NOS (N=24) | Schizophrenia (N=126)  Schizoaffective disorder (N=16)  Psychosis NOS (N=10) | Schizophrenia (N=67) | Schizophrenia (N=34) | Schizophrenia (N=22)  Psychosis NOS (N=2) |
| Mean dosage  ± SD (mg) | 342,3 ± 195,4  (N=659) | 312,2 ± 184,0  (N=389) | 336,0 ± 190,1  (N=146) | 414,4 ± 168,3  (N=66) | 439,7 ± 204,8  (N=34) | 533,3 ± 265,6  (N=24) |
| N smoking (%) | 393 (58,7)  (N=669) | 249 (56,6)  (N=407) | 101 (66,0)  (N=152) | 31 (45,6)  (N=61) | 9 (22,5)  (N=25) | 21 (87,5)  (N=24) |
| Mean Age at initiation of CLZ  ± SD (years) | 34,8 ± 11,1  (N=462) | 35,6 ± 11,4  (N=375) | NA | 32,7 ± 9,5  (N=54) | 29,5 ± 7,5  (N=33) | NA |
| Mean duration of CLZ therapy  ± SD (years) | 7,8 ± 7,5  (N=461) | 7,4 ± 7,7  (N=375) | NA | 8,0 ± 6,2  (N=53) | 12,6 ± 5,4  (N=33) | NA |
| Mean CLZ levels  ± SD (ng/mL) | 424,0 ± 248,5  (N=376) | 377,5 ± 185,9  (N=261) | NA | 449,2 ± 224,6  (N=67) | 874,3 ± 449,8  (N=24) | 407,9 ± 176,3  (N=24) |

**Supplementary Figure 2A&B.** Q-Q plots of the genome-wide association scan for quantitative outcome (A), and binary outcome (B).

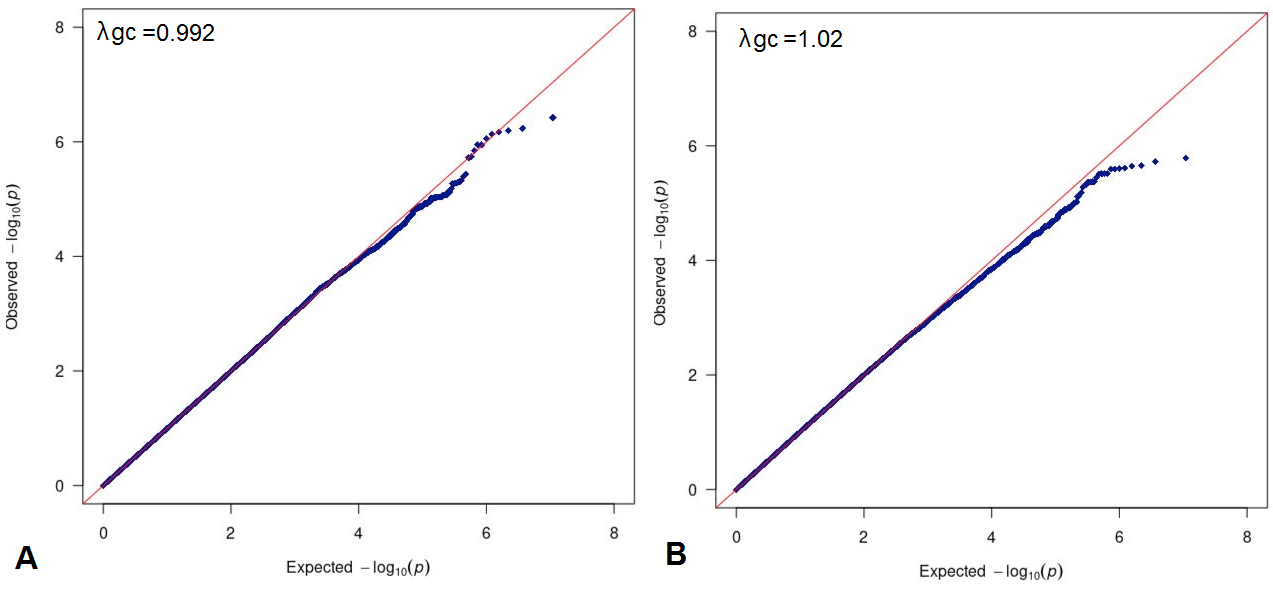
*Abbreviations:* Q-Q=quantile-quantile; λGC=genomic inflation correction factor.

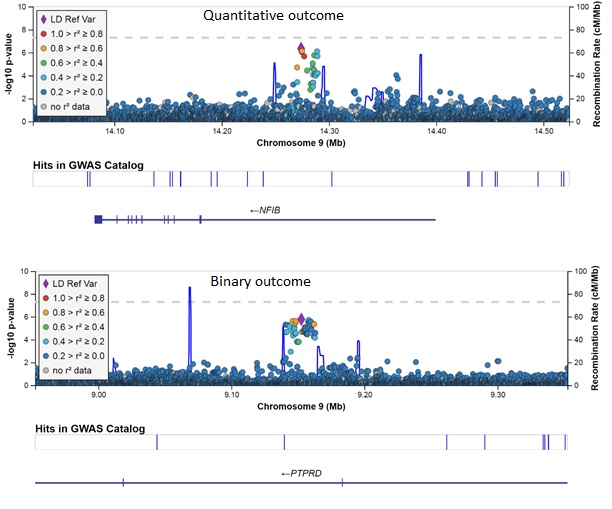
**Supplementary Figure 3A&B.** Regional association plots for the top SNPs (Single Nucleotide Polymorphisms) of quantitative outcome (rs1470431) and binary outcome (rs4742565), generated using LocusZoom (<http://csg.sph.umich.edu/locuszoom/>). The left Y-axis refers to the –Log 10 *p*-value corresponding to the association test between each SNP and symptom severity while on clozapine. SNPs are colored based on the level of linkage disequilibrium between each SNP and the index SNP. The diamond (shown in purple) is the most statistically significant SNP in the region.

**Supplementary Table 7.** Top SNPs identified in the genome-wide association analyses.
*Abbreviations:* SNP=Single Nucleotide Polymorphism, Chr=Chromosome, Pos=Position, A1=effect allele, A2=other allele, MAF=Minor Allele Frequency, β=Beta, OR=Odds Ratio.

| Outcome measure | Locus Name | SNP | Chr | Pos | A1/A2 | MAF (own datasat) | β/  OR | *P*-value | Function Reference Gene |
| --- | --- | --- | --- | --- | --- | --- | --- | --- | --- |
| Quantitative | *NFIB* | rs1923778 | 9 | 14274071 | T/C | 0.12 | β=-0.60 | 3.78x10^-7^ | Intron |
| Binary | *PTPRD* | rs4742565 | 9 | 9152508 | T/C | 0.47 | OR=0.56 | 1.64x10^-6^ | Intron |

**Supplementary Results**

**Other PRS-traits**
Schizophrenia-PRS was not significantly associated with quantitative outcome (*p*=0.12, R^2^=0.32, optimal p_t_=5.0x10^-6^, Supplementary Figure 9A).

Cross-disorder-PRS was only nominally significantly associated with binary outcome (*p*=0.04, R^2^=0.72, optimal p_t_=5x10^-7^, Supplementary Figure 9D).

Clozapine-levels-PRS was either not significantly or nominally significantly associated with quantitative or binary outcome (*p*=0.11, R^2^=0.34, p_t_=5x10^-5^, Supplementary Figure 8E, and *p*=0.03, R^2^=0.79, optimal p_t_=0.3, Supplementary Figure 9F, respectively).

**Sensitivity analyses**

For the GWAS, in sensitivity analyses results remained similar, with *p*=6.60x10^-6^ for the top locus (rs1923778) of quantitative outcome, and *p*=9.33x10^-7^ for the top locus (rs4742565) of binary outcome.
For PRS, in the sensitivity analyses results remained similar. For schizophrenia-PRS for quantitative outcome, the best fitting *p*-value threshold (p_t_) was 5x10^-6^, with *p=*0.07, and an explained variance (R^2^) of 0.45. Schizophrenia-PRS binary outcome: *p*=2.97x10^-3^, R^2^=1.58, p_t_=0.4. Cross-disorder-PRS quantitative outcome: *p*=0.03, R^2^=0.69, p_t_=0.2. Cross-disorder-PRS binary outcome: *p*=0.02, R^2^=0.92, p_t_=5x10^-7^. Clozapine-levels-PRS quantitative outcome: *p*=0.10, R^2^=0.37, p_t_=5x10^-7^. Clozapine-levels-PRS binary outcome: *p*=0.05, R^2^=0.71, p_t_=0.3.
For genotype-predicted enzyme activity score analyses, in the sensitivity analyses results remained similar. Higher *CYP2C19* activity score was significantly associated after multiple testing correction with a greater probability of low symptom severity (odds ratio (OR)=1.58, 95% confidence intervals (CI)=1.11-2.25, *p=*0.01, N=266 but not for quantitative outcome (beta=-0.15, *p=*0.12). *CYP2C19* activity score was not associated with dose-adjusted clozapine levels (beta=-0.11, *p*=0.08).
 **FUMA & H-MAGMA**
No gene or gene-set passed the genome-wide significance threshold of *p*<5x10^-8^ in our gene-based test. In addition, the top 5 genes associated with our phenotypes of interest were shown in Supplementary Figure 6A&B. Then, the gene-level summary statistics were used to create a differential gene expression analysis (DEG) (GTEx v8 and BrainSpan data) (Supplementary Figure 6A&B) and expression heatmaps (Supplementary Figure 10A&B). Tissue enrichment analysis did not yield any consistently associated tissues. For our quantitative GWAS, significantly enriched differentially expressed gene (DEG) sets were detected for hypothalamus and hippocampus for both down- and up- regulated DEG sites (Supplementary Figure 5A). Furthermore, tissue-specific expression pattern analysis based on GTEx v6 RNA-seq data^31^ for each gene show that for quantitative outcome the genes *FGP2, FGF1, NRCAM, KCNIP4,* and *PPFIA4* were highly specific expressed in brain regions (see heatmap in Supplementary Figure 10A).

Regarding fetal brain Hi-C, we identified 198 genes (including 58 protein coding genes) associated with binary phenotype in adult (*p*<5.0x10^-3^), and 269 genes (including 92 protein coding genes) associated with quantitative phenotype. Regarding adult brain Hi-C, we identified 158 genes (including 62 protein coding genes) associated with binary phenotype in adult, and 259 genes (including 86 protein coding genes) associated with quantitative phenotype (Supplementary Figure 11A-D). Five genes were common in 4 H-MAGMA analyses, namely, MCAT, LDHAL68, SUMO4, C19orf81, L1TD1. Rank-based gene ontology enrichment analysis suggested that symptom severity risk genes were enriched in the pathways such as Interleukin-36 pathway and cytokine receptor binding (Supplementary Figure 12A-D).

**Genotype-predicted enzyme activity score analyses**

Confirming previous findings,^32,33^ we found that examination of smoking status, independent of activity scores, showed that smokers had significantly lower dose-adjusted clozapine levels (Cohen’s d=0.76, *p=*1.65x10^-11^) and significantly greater CGI symptom severity scores relative to non-smokers (Cohen’s d=0.21; *p*=9.07x10^-3^; Supplementary Figure 13). Examination of concomitant use of an inhibitor of clozapine metabolism, independent of activity scores, showed those taking an inhibitor had significantly higher dose-adjusted clozapine levels (Cohen’s d=0.26, *p*=0.02) but they did not differ in symptom severity scores relative to those that were not taking an inhibitor (Cohen’s d=0.03, *p*=0.75).

**Supplementary Figure 4.** Dose-corrected clozapine concentrations in relation to the rs1923778 polymorphisms

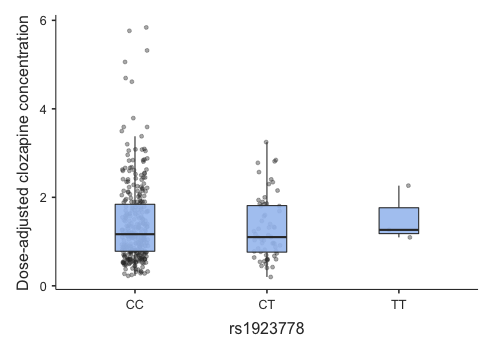

**Supplementary Table 8.** Tukey post-hoc test for dose-adjusted clozapine concentrations of the rs1923778 polymorphism.

|  | |  | | **CC** | | **CT** | | **TT** |
| --- | --- | --- | --- | --- | --- | --- | --- | --- |
| CC |  | Mean difference |  | — |  | 0.116 |  | -0.123 |
|  |  | p-value |  | — |  | 0.626 |  | 0.969 |
| CT |  | Mean difference |  |  |  | — |  | -0.239 |
|  |  | p-value |  |  |  | — |  | 0.892 |
| TT |  | Mean difference |  |  |  |  |  | — |
|  |  | p-value |  |  |  |  |  | — |

**Supplementary Figure 5A&B.** Tissue specificity results (enrichment test results of differentially expressed genes DEG sets for user-selected expression data sets) for quantitative outcome (A) and binary outcome (B). Significantly enriched DEG sets (*p*<0.05) are highlighted in red.

**A**
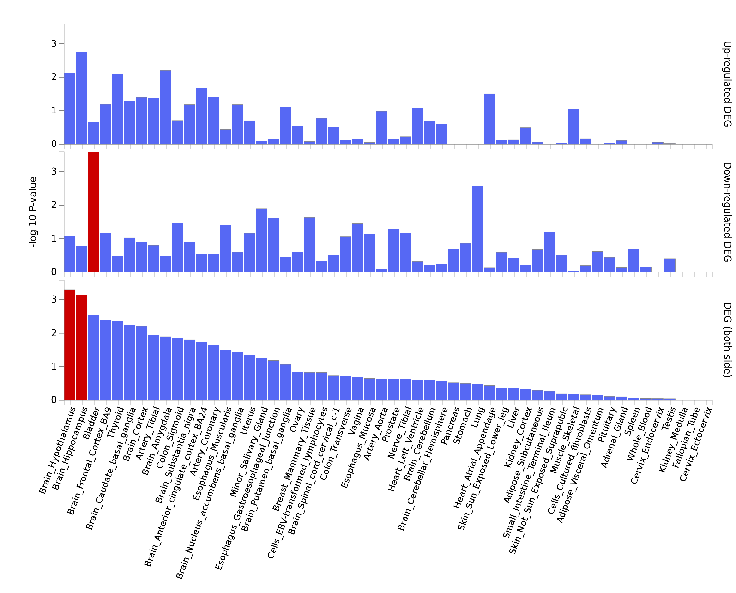

**B**
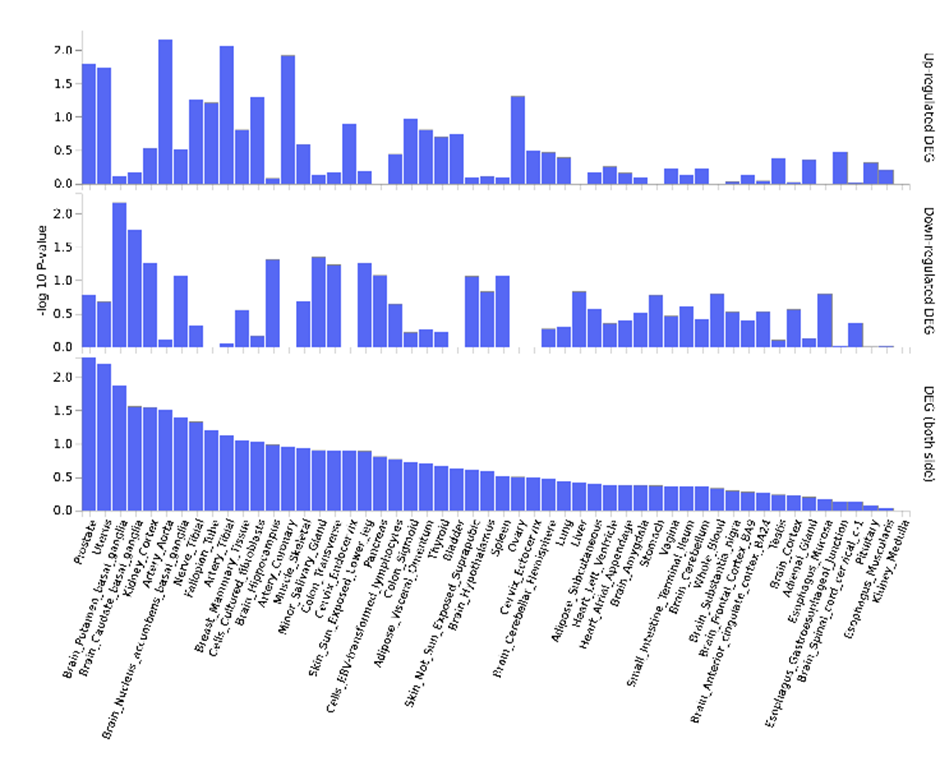

**Supplementary Figure 6A&B.** Gene-based test as computed by MAGMA based on our genome-wide association analysis summary statistics. Input SNPs were mapped to 17692 protein coding genes. Genome wide significance (red dashed line; no genes detected above this line) was defined at *p*=0.05/17692=2.82x10^-6^. For both quantitative outcome (A) as binary outcome (B) no genes were significant. Top 5 genes are labeled.

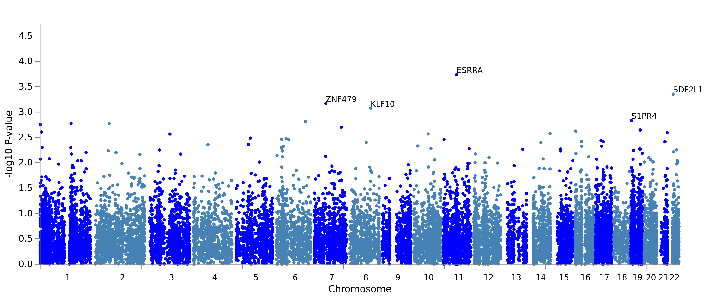

**A**

**B**
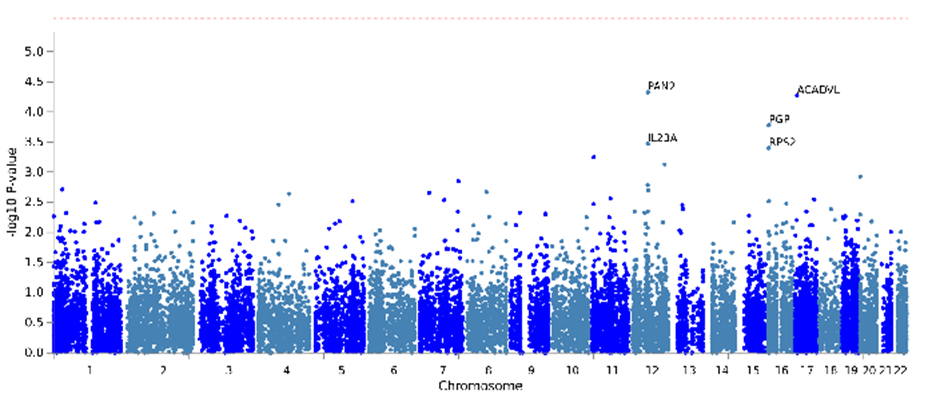

**Supplementary Table 9.** Positive predicted values and odds ratios with 95% confident intervals for binary outcome based on various cut-offs of schizophrenia-PRS.
*Abbreviations:* PPV=Positive Predicted Value, OR=Odds ratio, PRS=Polygenic Risk Score.

| Cut-off based on percentile | PPV | PPV-lower | PPV-upper | OR | OR-lower | OR-upper | *P*-value |
| --- | --- | --- | --- | --- | --- | --- | --- |
| <25 | 0.409 | 0.344 | 0.477 | 0.674 | 0.475 | 0.958 | **0.028** |
| <33 | 0.482 | 0.445 | 0.520 | 0.601 | 0.435 | 0.831 | **2.1x10^-3^** |
| >67 | 0.562 | 0.506 | 0.616 | 1.611 | 1.169 | 2.221 | **4.2x10^-3^** |
| >75 | 0.567 | 0.499 | 0.633 | 1.575 | 1.111 | 2.233 | **0.013** |

**Supplementary Table 10.** Positive predicted values and odds ratios for binary outcome (for high symptom severity) for the highest decile (>90) vs. lowest decile (<10) and highest tertile (>67) vs. lowest tertile (<33) of schizophrenia-PRS.
*Abbreviations:* PPV=Positive Predicted Value, OR=Odds ratio, PRS=Polygenic Risk Score.

| Cut-off | PPV | PPV-lower | PPV-upper | OR | OR-lower | OR-upper | *P*-value |
| --- | --- | --- | --- | --- | --- | --- | --- |
| Top 10 vs. tail 10 | 0.366 | 0.289 | 0.447 | 2.257 | 1.298 | 3.921 | 3.96x10^-3^ |
| Top 33 vs. tail 67 | 0.398 | 0.352 | 0.446 | 1.938 | 1.333 | 2.981 | 6.84x10^-4^ |

**Supplementary Figure 7A-F.** Bar plots illustrating the explained variance for the association of the three different PRS-traits at several *p*-value thresholds (p_t_) for quantitative outcome and binary outcome, adjusted for sex, age, and 10 PCs. p_t_ are displayed on the X axis, where the number of included SNPs increases with more lenient p_t._ ΔExplained variance represents the Nagelkerke R^2^ (shown as %). The red dots represent the strengths of the association results (-Log 10 *p-*value). The dashed line represents a significance-level of *p*-value*<*0.05.

*Abbreviations:* PRS=Polygenic Risk Score, PCs=Principal Components, SNPs=Single Nucleotide Polymorphisms.

**A.** Schizophrenia-PRS for quantitative outcome	**B.** Schizophrenia-PRS for binary outcome

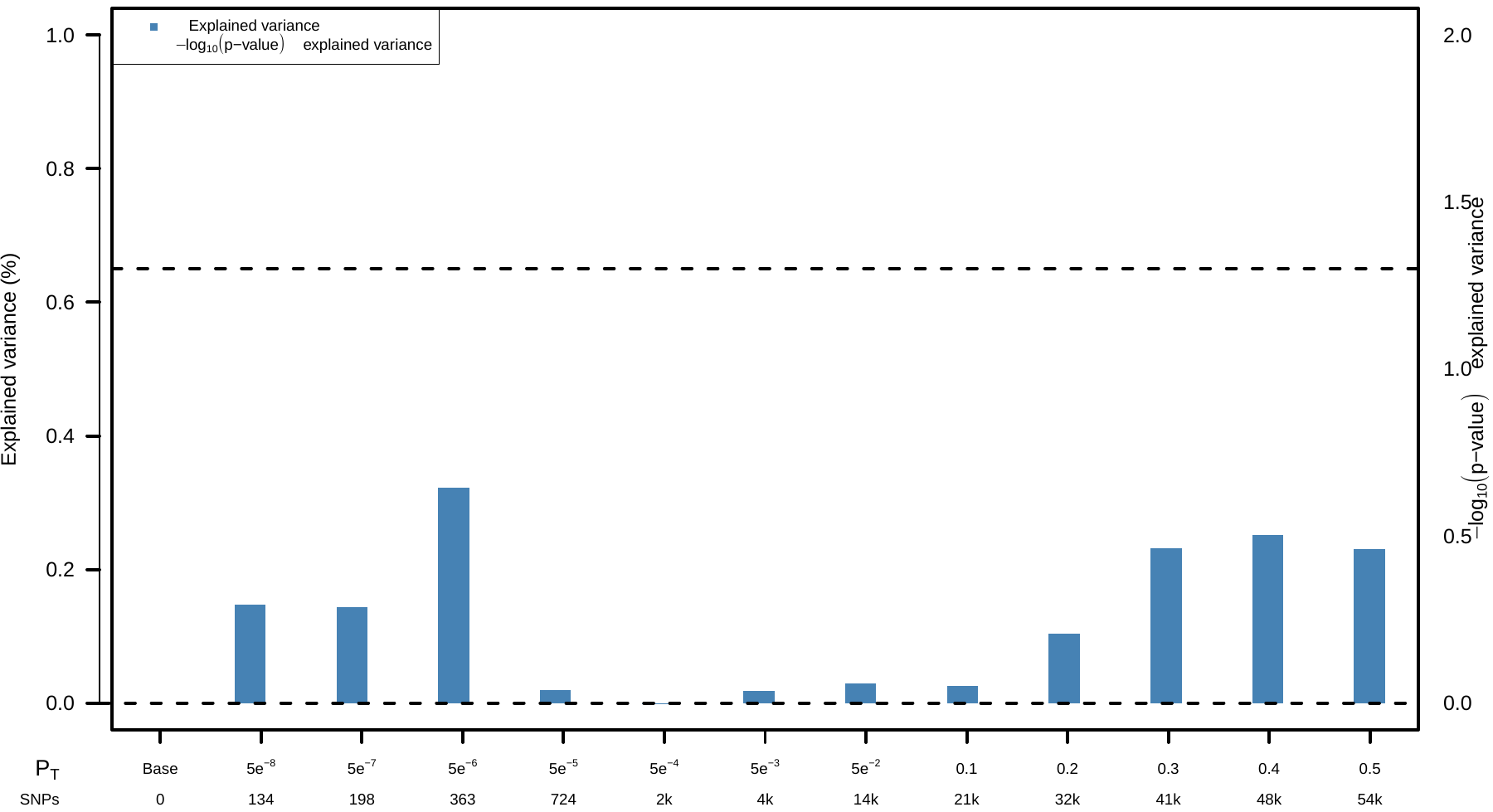

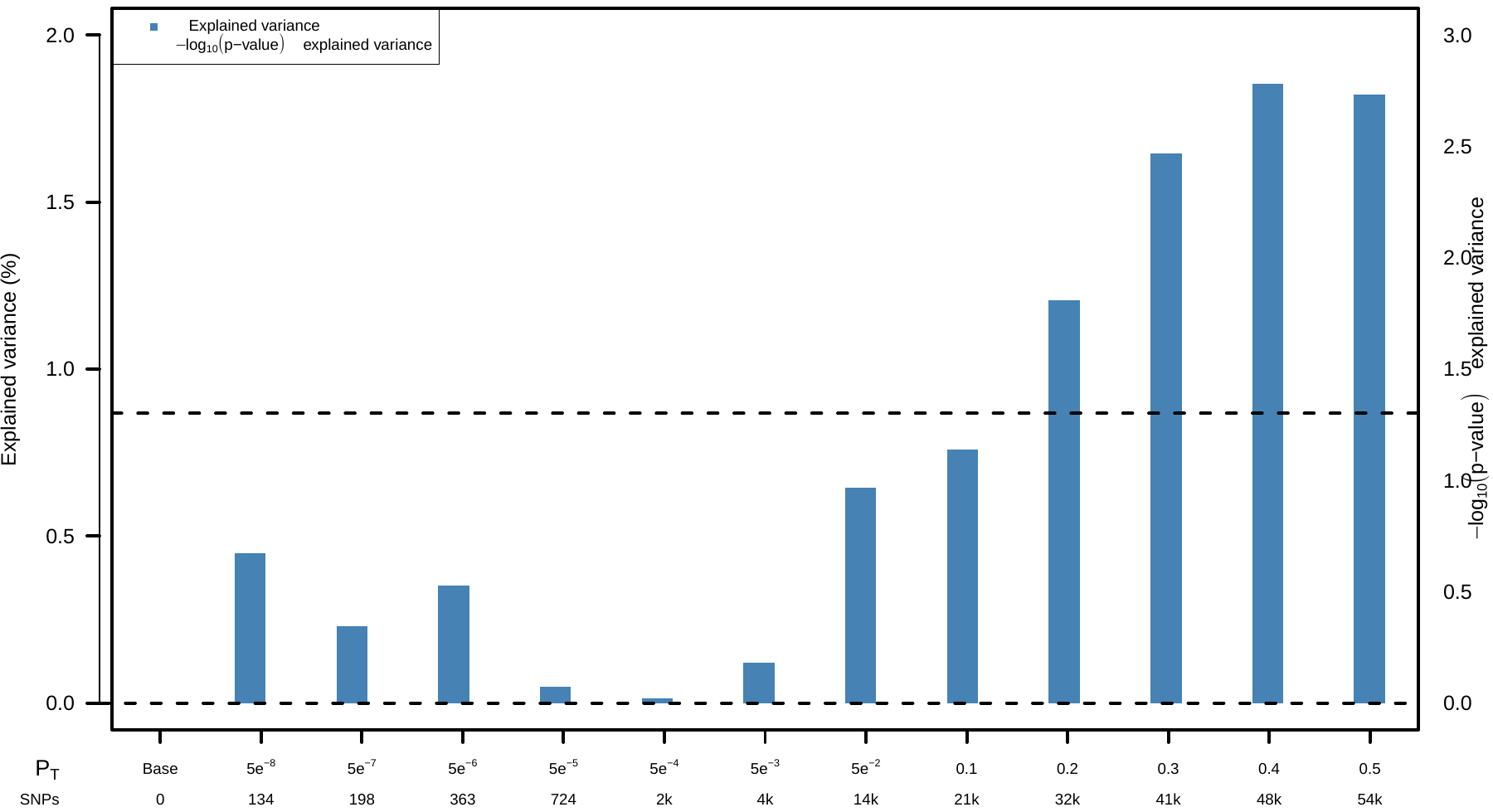

**C.** Cross-disorder-PRS for quantitative outcome	**D.** Cross-disorder-PRS for binary outcome
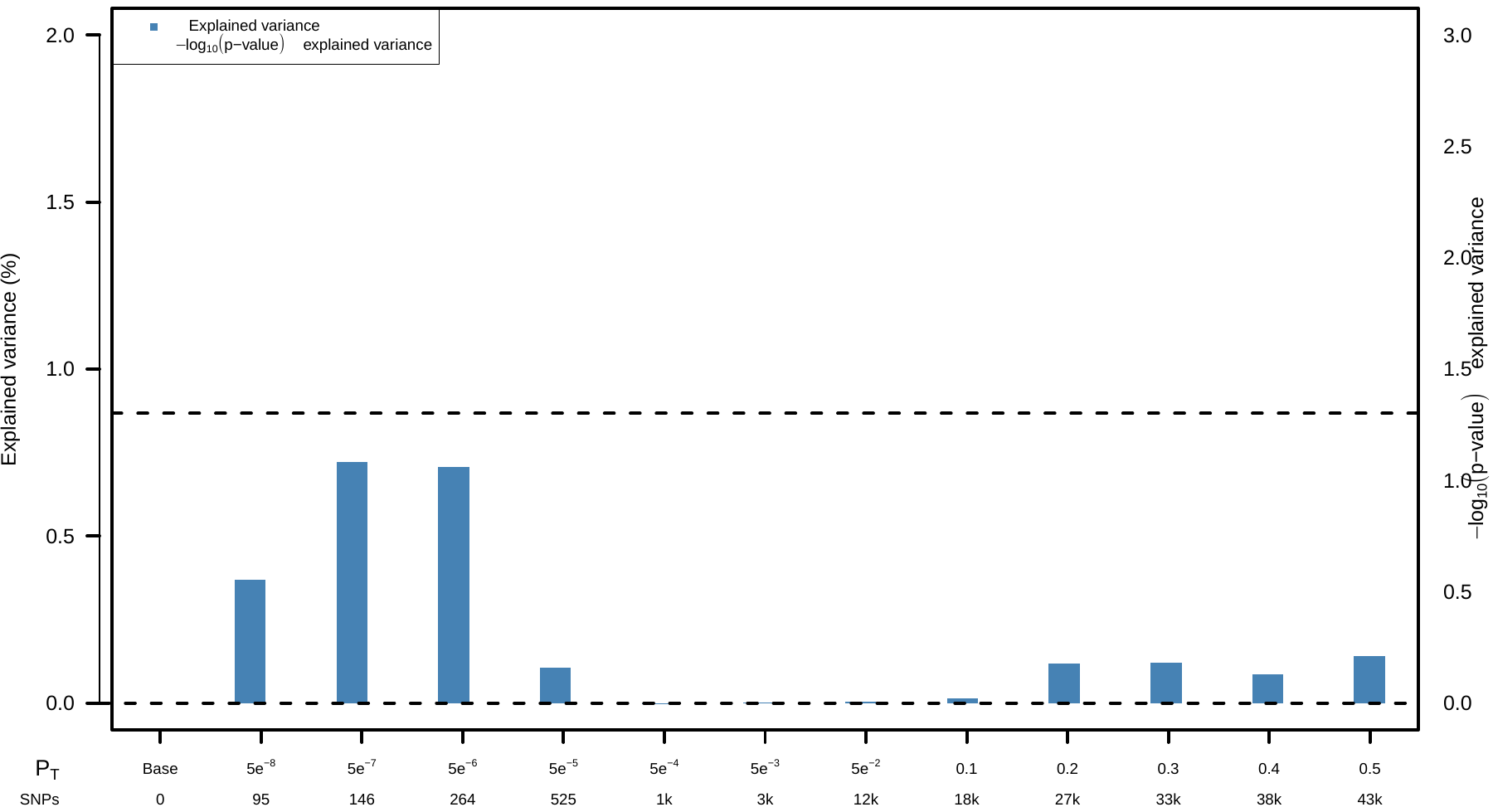

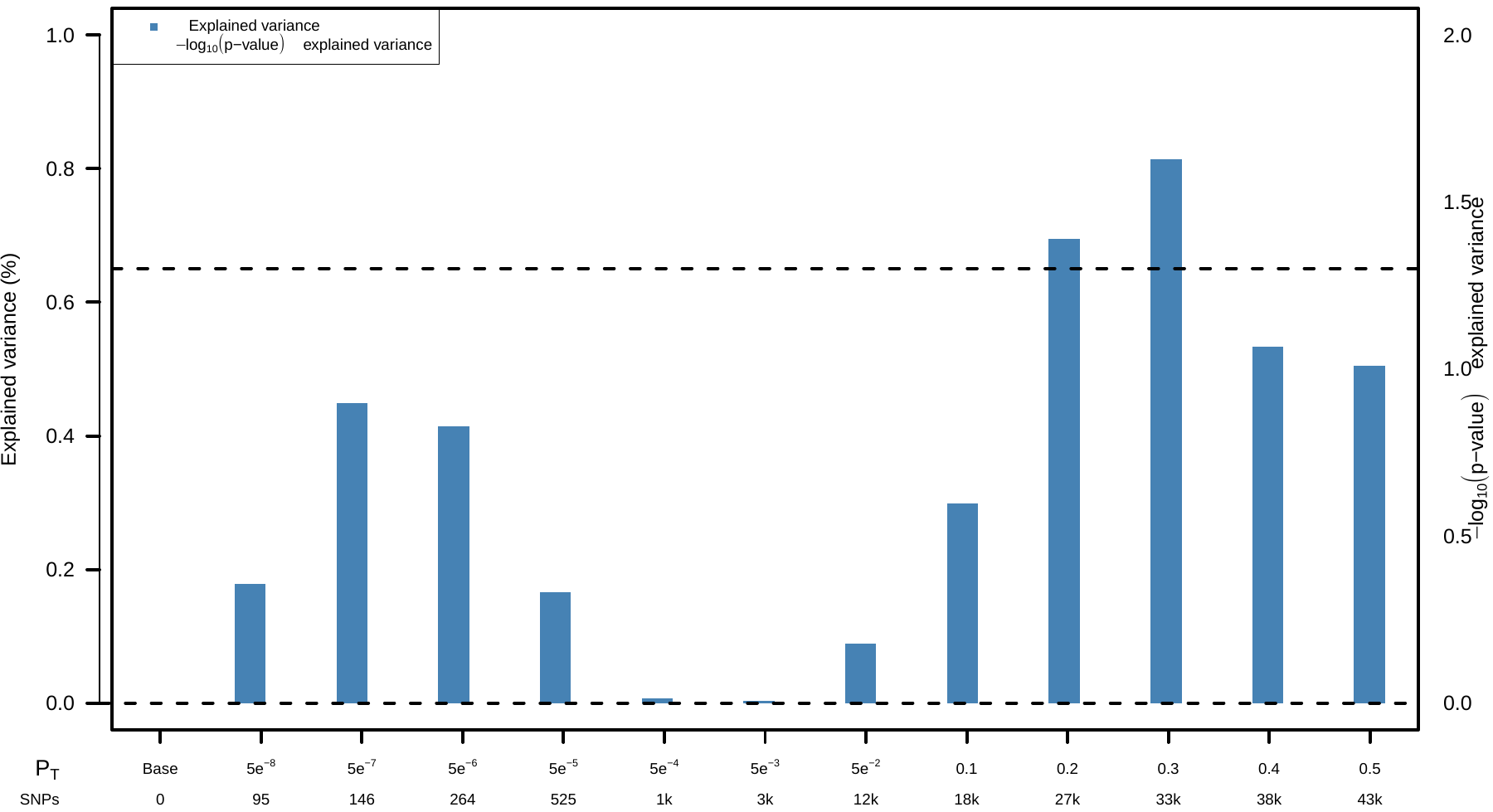

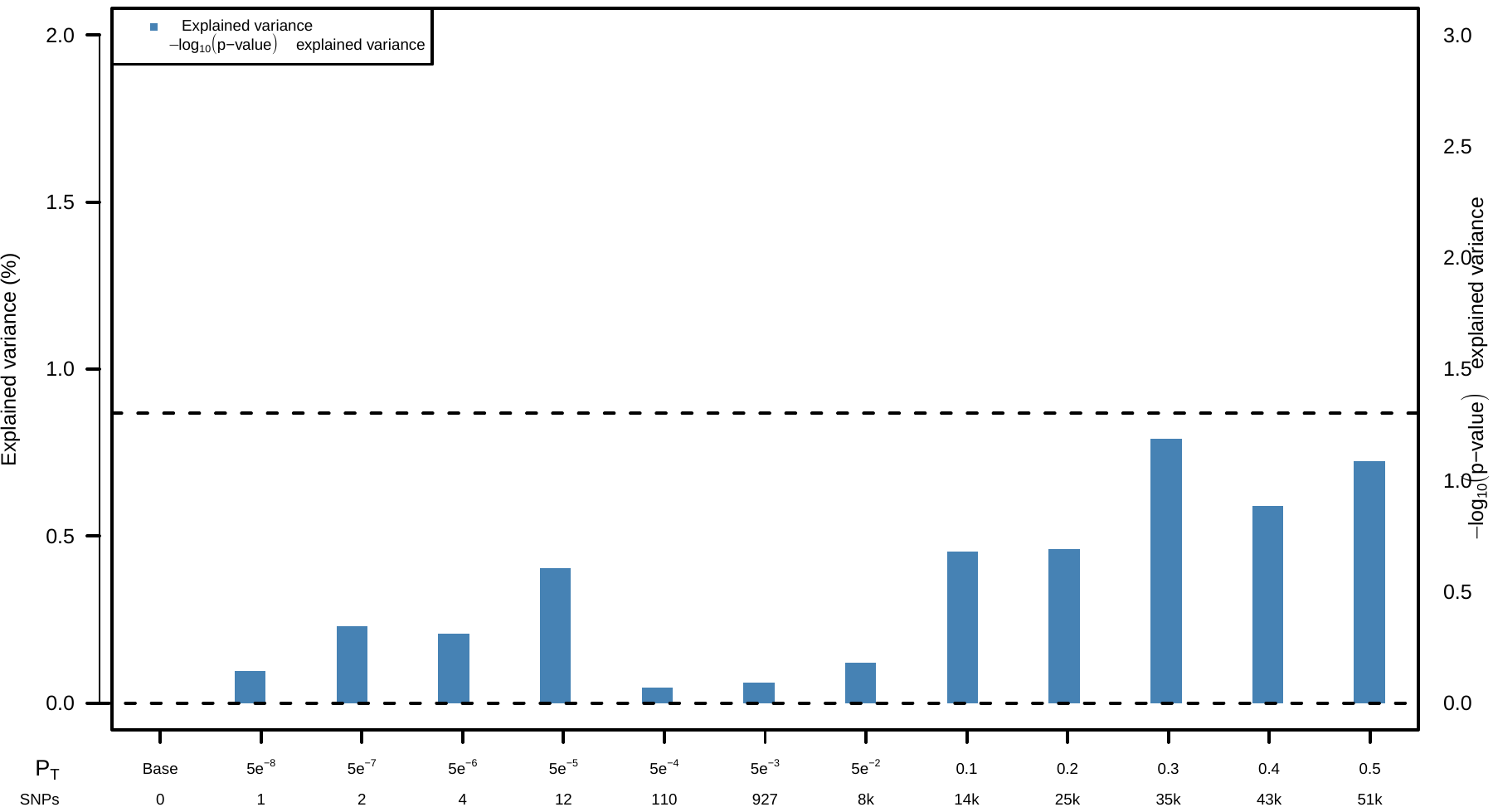

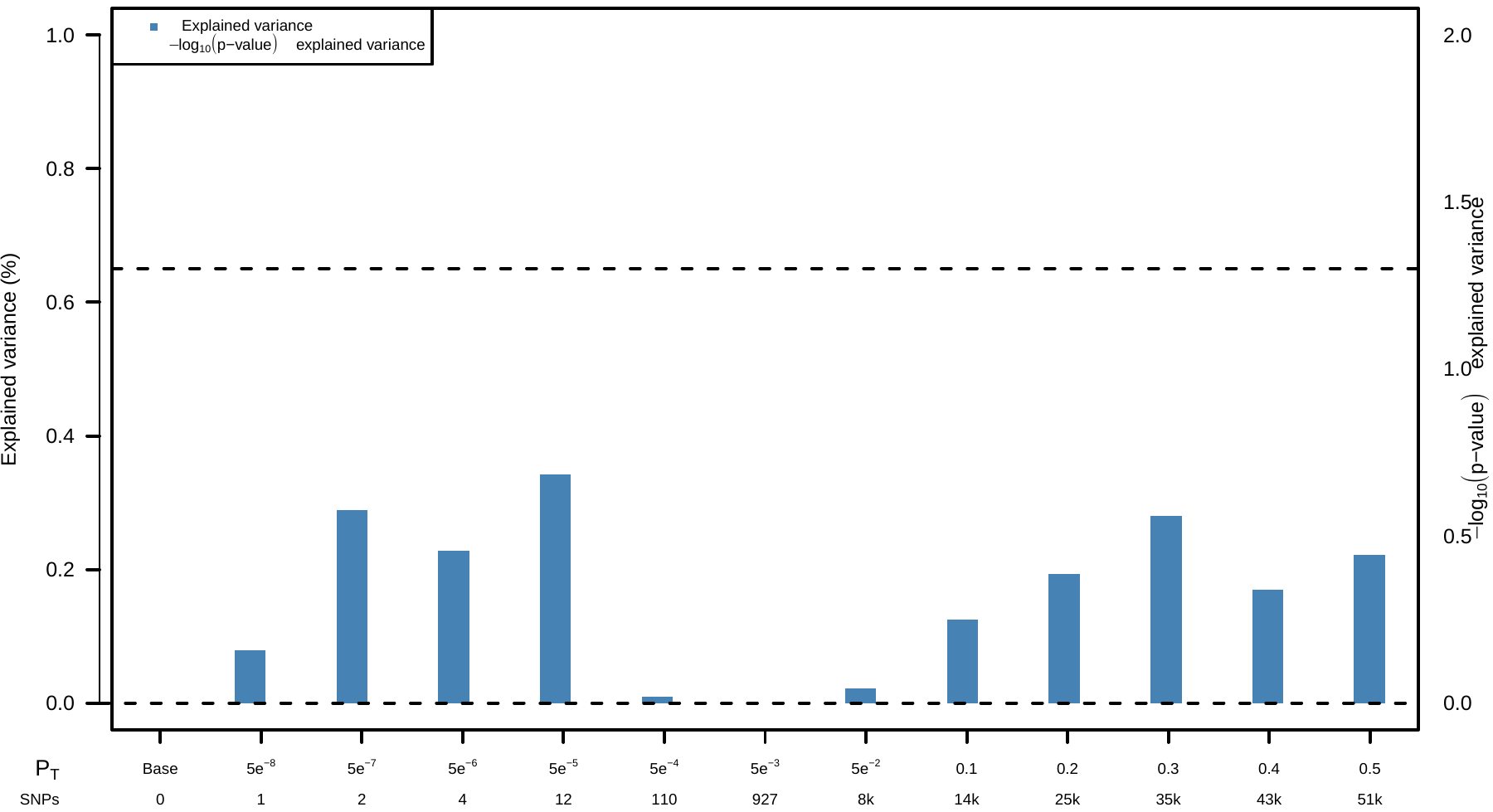
**E.** Clozapine-levels-PRS for quantitative outcome	**F.** Clozapine-levels-PRS for binary outcome

| **Supplementary Table 11.** Logistic regression model for genotype-predictive enzyme activity scores of binary outcome (low vs. high symptom severity among clozapine users; N=291). | | | | | | | | | | | | | | | | | | | |
| --- | --- | --- | --- | --- | --- | --- | --- | --- | --- | --- | --- | --- | --- | --- | --- | --- | --- | --- | --- |
|  | | | | **95% Confidence Interval** | | | |  | | | | | | | | | **95% Confidence Interval** | | |
| **Predictor** | | **Estimate** | | **Lower** | | **Upper** | | **SE** | | **Z** | | ***p*** | | **Odds ratio** | | | **Lower** | | **Upper** |
| Intercept |  | -0.67627 |  | -2.2813 |  | 0.9288 |  | 0.8189 |  | -0.8258 |  | 0.41 |  | | 0.509 |  | 0.102 |  | 2.53 |
| Sex: Female (male reference) |  | -0.18687 |  | -0.7385 |  | 0.3647 |  | 0.2814 |  | -0.6640 |  | 0.51 |  | | 0.830 |  | 0.478 |  | 1.44 |
| Age (years) |  | -0.01161 |  | -0.0358 |  | 0.0125 |  | 0.0123 |  | -0.9429 |  | 0.35 |  | | 0.988 |  | 0.965 |  | 1.01 |
| Dose-adjusted clozapine levels (ng/mL/mg/d) |  | -0.05760 |  | -0.3634 |  | 0.2482 |  | 0.1560 |  | -0.3692 |  | 0.71 |  | | 0.944 |  | 0.695 |  | 1.28 |
| Duration of clozapine (months) |  | 0.04832 |  | 0.0116 |  | 0.0851 |  | 0.0188 |  | 2.5766 |  | **0.01** |  | | 1.050 |  | 1.012 |  | 1.09 |
| CYP2C19 activity score |  | 0.46319 |  | 0.1185 |  | 0.8078 |  | 0.1758 |  | 2.6340 |  | **8.44x10^-3^** |  | | 1.589 |  | 1.126 |  | 2.24 |
| CYP1A2 activity score |  | -0.00141 |  | -0.2237 |  | 0.2209 |  | 0.1134 |  | -0.0125 |  | 0.99 |  | | 0.999 |  | 0.800 |  | 1.25 |
| CYP2D6 activity score |  | -0.17042 |  | -0.5523 |  | 0.2114 |  | 0.1948 |  | -0.8747 |  | 0.38 |  | | 0.843 |  | 0.576 |  | 1.24 |
| Note: activity scores were adjusted for concomitant use of known inhibitors or inducers.  *Abbreviations:* SE=Standard Error, Z=Z-score, *p*=*p*-value. | | | | | | | | | | | | | | | | | | | |

**Supplementary Table 12.** Linear regression model for genotype-predictive enzyme activity scores of quantitative outcome (N=291).

*Abbreviations:* SE=Standard Error, t=t-statistic, *p*=*p*-value, Stand. Estimate=standard estimate.

|  | | | **95% Confidence Interval** | |  | | | **95% Confidence Interval** | |
| --- | --- | --- | --- | --- | --- | --- | --- | --- | --- |
| **Predictor** | **Estimate** | **SE** | **Lower** | **Upper** | **t** | ***p*** | **Stand. Estimate** | **Lower** | **Upper** |
| Intercept | 3.85858 | 0.47424 | 2.92511 | 4.7920 | 8.13640 | 2.96x10^-13^ |  |  |  |
| Sex: Female (male reference) | 0.17616 | 0.15597 | -0.13084 | 0.4832 | 1.12946 | 0.26 | 0.0682 | -0.0506 | 0.1870 |
| Age (years) | 0.00538 | 0.00684 | -0.00809 | 0.0188 | 0.78620 | 0.43 | 0.0507 | -0.0762 | 0.1776 |
| Dose-adjusted clozapine levels (ng/mL/mg/d) | -3.37x10^-4^ | 0.08738 | -0.17233 | 0.1717 | -0.00385 | 1.00 | -2.45x10^-4^ | -0.1252 | 0.1247 |
| Duration of clozapine (months) | -0.03957 | 0.01038 | -0.06000 | -0.0191 | -3.81233 | 1.69x10^-4^ | -0.2424 | -0.3675 | -0.1172 |
| CYP2C19 Activity Score | -0.15857 | 0.09487 | -0.34531 | 0.0282 | -1.67150 | 0.10 | -0.0960 | -0.2092 | 0.0171 |
| CYP1A2 Activity Score | 0.05059 | 0.06375 | -0.07489 | 0.1761 | 0.79360 | 0.43 | 0.0492 | -0.0728 | 0.1712 |
| CYP2D6 Activity Score | 0.14289 | 0.10880 | -0.07128 | 0.3571 | 1.31326 | 0.19 | 0.0751 | -0.0375 | 0.1877 |

| **Supplementary Table 13.** Linear regression model for genotype-predictive enzyme activity scores of dose-adjusted clozapine levels (N=291).  Note: activity scores were adjusted for concomitant use of known inhibitors or inducers.  *Abbreviations:* B=Beta, SE=Standard Error, Z=Z-score, *p*=*p*-value. | | | | | | | | | | | | | | | | | | |
| --- | --- | --- | --- | --- | --- | --- | --- | --- | --- | --- | --- | --- | --- | --- | --- | --- | --- | --- |
|  | | | | | | **95% Confidence Interval** | | | |  | | | | | | **95% Confidence Interval** | | |
| **Predictor** | | **B** | | **SE** | | **Lower** | | **Upper** | | **t** | | ***p*** | | **Beta** | | **Lower** | | **Upper** |
| Intercept |  | 2.60936 |  | 0.28189 |  | 2.05451 |  | 3.16420 |  | 9.257 |  | 5.88x10^-11^ |  |  |  |  |  |  |
| Sex: Female (male reference) |  | 0.40061 |  | 0.10303 |  | 0.19781 |  | 0.60342 |  | 3.888 |  | 1.26x10^-4^ |  | 0.2134 |  | 0.1054 |  | 0.3215 |
| Age (years) |  | -0.00583 |  | 0.00463 |  | -0.01494 |  | 0.00327 |  | -1.261 |  | 0.21 |  | -0.0756 |  | -0.1937 |  | 0.0424 |
| Duration of clozapine (months) |  | 0.01209 |  | 0.00700 |  | -0.00168 |  | 0.02587 |  | 1.728 |  | 0.09 |  | 0.1019 |  | -0.0142 |  | 0.2181 |
| CYP2C19 activity score |  | -0.09752 |  | 0.06405 |  | -0.22359 |  | 0.02856 |  | -1.522 |  | 0.13 |  | -0.0813 |  | -0.1864 |  | 0.0238 |
| CYP1A2 activity score |  | -0.26383 |  | 0.04029 |  | -0.34313 |  | -0.18453 |  | -6.548 |  | **2.71x10^-10^** |  | -0.3531 |  | -0.4593 |  | -0.2470 |
| CYP2D6 activity score |  | 0.03713 |  | 0.07372 |  | -0.10799 |  | 0.18224 |  | 0.504 |  | 0.62 |  | 0.0269 |  | -0.0782 |  | 0.1319 |

| **Supplementary Table 14.** Combined pharmacogenetic and PRS logistic regression model of binary outcome (N=291).  *Abbreviations:* SE=Standard Error, Z=Z-score, *p*=*p*-value, SCZ-PRS=Schizophrenia Polygenic Risk Score. | | | | | | | | | | | | | | |
| --- | --- | --- | --- | --- | --- | --- | --- | --- | --- | --- | --- | --- | --- | --- |
|  | | | | | | | | | | | | **95% Confidence Interval** | | |
| **Predictor** | | **B** | | **SE** | | **Z** | | ***p*** | | **Odds ratio** | | **Lower** | | **Upper** |
| Intercept |  | -0.03242 |  | 1.5394 |  | -0.02106 |  | 0.98 |  | 0.968 |  | 0.0474 |  | 19.78 |
| Sex: Female (male reference) |  | -0.18393 |  | 0.2817 |  | -0.65297 |  | 0.51 |  | 0.832 |  | 0.4790 |  | 1.45 |
| Age (years) |  | -0.01114 |  | 0.0124 |  | -0.90091 |  | 0.37 |  | 0.989 |  | 0.9652 |  | 1.01 |
| Dose-adjusted clozapine levels (ng/mL/mg/d) |  | -0.05750 |  | 0.1561 |  | -0.36849 |  | 0.71 |  | 0.944 |  | 0.6953 |  | 1.28 |
| Duration of clozapine (months) |  | 0.04853 |  | 0.0188 |  | 2.58688 |  | **0.01** |  | 1.050 |  | 1.0118 |  | 1.09 |
| CYP2C19 Activity Score |  | 0.46247 |  | 0.1758 |  | 2.63120 |  | **8.51x10^-3^** |  | 1.588 |  | 1.1252 |  | 2.24 |
| CYP1A2 Activity Score |  | 5.91x10^-4^ |  | 0.1136 |  | 5.20x10^-3^ |  | 1.00 |  | 1.001 |  | 0.8008 |  | 1.25 |
| CYP2D6 Activity Score |  | -0.17617 |  | 0.1953 |  | -0.90207 |  | 0.37 |  | 0.838 |  | 0.5718 |  | 1.23 |
| SCZ-PRS |  | 8.53 x10^-4^ |  | 0.0173 |  | 0.49278 |  | 0.62 |  | 1.009 |  | 0.9749 |  | 1.04 |

| **Supplementary Table 15.** Combined pharmacogenetic and PRS with principal components (PCs) logistic regression model of binary outcome (N=291).  *Abbreviations:* PC= Principal Component, SE=Standard Error, Z=Z-score, p=p-value, SCZ-PRS=Schizophrenia Polygenic Risk Score | | | | | | | | | | | | | | |
| --- | --- | --- | --- | --- | --- | --- | --- | --- | --- | --- | --- | --- | --- | --- |
|  | | | | | | | | | | | | **95% Confidence Interval** | | |
| **Predictor** | | **B** | | **SE** | | **Z** | | ***p*** | | **Odds ratio** | | **Lower** | | **Upper** |
| Intercept |  | 5.1265 |  | 3.2281 |  | 1.588 |  | 0.11 |  | 168.429 |  | 0.30104 |  | 94233.85 |
| Sex: Female (male reference) |  | -0.1247 |  | 0.3108 |  | -0.401 |  | 0.69 |  | 0.883 |  | 0.48000 |  | 1.62 |
| Age (years) |  | -0.0182 |  | 0.0136 |  | -1.338 |  | 0.18 |  | 0.982 |  | 0.95622 |  | 1.01 |
| Dose-adjusted clozapine levels (ng/mL/mg/d) |  | 0.0461 |  | 0.1714 |  | 0.269 |  | 0.79 |  | 1.047 |  | 0.74838 |  | 1.47 |
| Duration of clozapine (months) |  | 0.0519 |  | 0.0206 |  | 2.514 |  | **0.01** |  | 1.053 |  | 1.01151 |  | 1.10 |
| CYP2C19 Activity Score |  | 0.4872 |  | 0.2038 |  | 2.391 |  | **0.02** |  | 1.628 |  | 1.09178 |  | 2.43 |
| CYP1A2 Activity Score |  | 0.1111 |  | 0.1246 |  | 0.892 |  | 0.37 |  | 1.118 |  | 0.87543 |  | 1.43 |
| CYP2D6 Activity Score |  | -0.0571 |  | 0.2166 |  | -0.264 |  | 0.79 |  | 0.944 |  | 0.61775 |  | 1.44 |
| SCZ-PRS |  | 0.0779 |  | 0.0404 |  | 1.927 |  | 0.05 |  | 1.081 |  | 0.99866 |  | 1.17 |
| PC1 |  | -7.6075 |  | 11.4711 |  | -0.663 |  | 0.51 |  | 4.97x10^-4^ |  | 8.55x10^-14^ |  | 2.89x10^6^ |
| PC2 |  | 16.5906 |  | 10.8711 |  | 1.526 |  | 0.13 |  | 1.60x10^7^ |  | 8.95x10^-3^ |  | 2.88x10^16^ |
| PC3 |  | 7.3990 |  | 12.2345 |  | 0.605 |  | 0.55 |  | 1634.398 |  | 6.30x10^-8^ |  | 4.24x10^13^ |
| PC4 |  | -87.2468 |  | 20.0057 |  | -4.361 |  | 1.30x10^-5^ |  | 1.29x10^-38^ |  | 1.20x10^-55^ |  | 1.37x10^-21^ |
| PC5 |  | 54.7994 |  | 18.0575 |  | 3.035 |  | 2.41x10^-3^ |  | 6.30x10^23^ |  | 2.68x10^8^ |  | 1.48x10^39^ |
| PC6 |  | -13.4428 |  | 17.8121 |  | -0.755 |  | 0.45 |  | 1.45x10^-6^ |  | 1.00x10^-21^ |  | 2.11x10^9^ |
| PC7 |  | 24.0574 |  | 15.3150 |  | 1.571 |  | 0.12 |  | 2.81x10^10^ |  | 2.58x10^-3^ |  | 3.05x10^23^ |
| PC8 |  | 6.9594 |  | 15.3252 |  | 0.454 |  | 0.65 |  | 1052.968 |  | 9.50x10^-11^ |  | 1.17x10^16^ |
| PC9 |  | 7.0306 |  | 14.6582 |  | 0.480 |  | 0.63 |  | 1130.691 |  | 3.77x10^-10^ |  | 3.39x10^15^ |
| PC10 |  | 36.6766 |  | 14.6954 |  | 2.496 |  | 0.01 |  | 8.48x10^15^ |  | 2628.22090 |  | 2.74x10^28^ |

| **Supplementary Table 16.** Combined pharmacogenetic and top GWA hits with principal components (PCs) logistic regression model of binary outcome (N=291).  *Abbreviations:* PC= Principal Component, SE=Standard Error, Z=Z-score, p=p-value, SCZ-PRS=Schizophrenia Polygenic Risk Score | | | | | | | | | | | | | | | | | | | | | |
| --- | --- | --- | --- | --- | --- | --- | --- | --- | --- | --- | --- | --- | --- | --- | --- | --- | --- | --- | --- | --- | --- |
|  | | | | | | | | | | | | | | | **95% Confidence Interval** | | | | | | |
| **Predictor** | **Estimate** | | | **SE** | | | **Z** | | | | **p** | | **OR** | | | **Lower** | | | **Upper** | | |
| Intercept |  | -1.7034 |  | | 0.9615 |  | | -1.772 |  | 0.076 | |  | | 0.182 | | |  | 0.0277 | |  | 1.20 |
| Sex: Female (male reference) |  | -0.1210 |  | | 0.3218 |  | | -0.376 |  | 0.707 | |  | | 0.886 | | |  | 0.4716 | |  | 1.66 |
| Age (years) |  | -0.0169 |  | | 0.0138 |  | | -1.224 |  | 0.221 | |  | | 0.983 | | |  | 0.9570 | |  | 1.01 |
| Dose-adjusted clozapine levels (ng/mL/mg/d) |  | 0.0340 |  | | 0.1797 |  | | 0.189 |  | 0.850 | |  | | 1.035 | | |  | 0.7275 | |  | 1.47 |
| Duration of clozapine (months) |  | 0.0465 |  | | 0.0213 |  | | 2.187 |  | **0.029** | |  | | 1.048 | | |  | 1.0048 | |  | 1.09 |
| CYP2C19 Activity Score |  | 0.4719 |  | | 0.2094 |  | | 2.254 |  | **0.024** | |  | | 1.603 | | |  | 1.0635 | |  | 2.42 |
| CYP1A2 Activity Score |  | 0.1492 |  | | 0.1276 |  | | 1.169 |  | 0.242 | |  | | 1.161 | | |  | 0.9040 | |  | 1.49 |
| CYP2D6 Activity Score |  | -0.0585 |  | | 0.2198 |  | | -0.266 |  | 0.790 | |  | | 0.943 | | |  | 0.6131 | |  | 1.45 |
| PC1 |  | 13.8485 |  | | 6.6642 |  | | 2.078 |  | 0.038 | |  | | 1.03x10^6^ | | |  | 2.1967 | |  | 4.86x10^11^ |
| PC2 |  | 7.4986 |  | | 10.9167 |  | | 0.687 |  | 0.492 | |  | | 1805.442 | | |  | 9.21x10^-7^ | |  | 3.54x10^12^ |
| PC3 |  | 16.7976 |  | | 12.4098 |  | | 1.354 |  | 0.176 | |  | | 1.97x10^7^ | | |  | 5.39x10^-4^ | |  | 7.22x10^17^ |
| PC4 |  | -78.0728 |  | | 19.8746 |  | | -3.928 |  | < .001 | |  | | 1.24x10^34^ | | |  | 1.50x10^-51^ | |  | 1.03x10^-17^ |
| PC5 |  | 58.1537 |  | | 18.7738 |  | | 3.098 |  | 0.002 | |  | | 1.80x10^25^ | | |  | 1.89x10^9^ | |  | 1.72x10^41^ |
| PC6 |  | -14.5085 |  | | 18.4950 |  | | -0.784 |  | 0.433 | |  | | 5.00x10^-7^ | | |  | 9.04x10^-23^ | |  | 2.77x10^9^ |
| PC7 |  | 17.9347 |  | | 15.3928 |  | | 1.165 |  | 0.244 | |  | | 6.15x10^7^ | | |  | 4.86x10^-6^ | |  | 7.79x10^20^ |
| PC8 |  | 7.8858 |  | | 15.7144 |  | | 0.502 |  | 0.616 | |  | | 2659.327 | | |  | 1.12x10^-10^ | |  | 6.32x10^16^ |
| PC9 |  | 3.2391 |  | | 15.1423 |  | | 0.214 |  | 0.831 | |  | | 25.511 | | |  | 3.29x10^-12^ | |  | 1.98x10^14^ |
| PC10 |  | 38.6448 |  | | 14.8678 |  | | 2.599 |  | 0.009 | |  | | 6.07x10^16^ | | |  | 13419.3970 | |  | 2.75x10^29^ |
| rs4742565 (PTPRD) |  | 0.7265 |  | | 0.1946 |  | | 3.734 |  | **1.86x10^-4^** | |  | | 2.068 | | |  | 1.4121 | |  | 3.03 |
| rs1923778 (NFIB) |  | 0.1186 |  | | 0.3613 |  | | 0.328 |  | 0.743 | |  | | 1.126 | | |  | 0.5546 | |  | 2.29 |

**Supplementary Table 17**. Descriptive statistics on dose-adjusted clozapine concentrations and high vs. low symptom severity. Abbreviations: N=number of participants, SD=standard deviation, SE=standard error.

|  | | | | | | | | | | | | |
| --- | --- | --- | --- | --- | --- | --- | --- | --- | --- | --- | --- | --- |
|  | | **Group** | | **N** | | **Mean** | | **Median** | | **SD** | | **SE** |
| Dose-adjusted clozapine concentration |  | High |  | 210 |  | 1.42 |  | 1.14 |  | 0.911 |  | 0.0628 |
|  | | Low |  | 159 |  | 1.37 |  | 1.16 |  | 0.858 |  | 0.0681 |
| **Supplementary Table 18**. Results of an Independent Samples T-Test between dose-adjusted clozapine concentrations and high vs. low symptom severity. Abbreviations: df=degrees of freedom, p=*p*-value. \| \| \| \| \| \| \| \| \|; \| \| \| \| **Statistic** \| \| **df** \| \| **p** \|; Dose-adjusted clozapine concentration \| \| Student's t \| \| 0.566 \| \| 367 \| \| 0.572 \| | | | | | | | | | | | | |

**Supplementary Table 19.** Linkage disequilibrium statistics calculated with Plink 1.9, to inspect the relation between our two top hits and the top hits of previous performed, relevant genome-wide association studies.

|  |  | **rs28379954**  Smith et al.^34^ | **rs2093483**  Li et al.^35^ | **rs1500318**  Li et al.^35^ |
| --- | --- | --- | --- | --- |
| **rs1923778** | Quantitative top locus | R^2^=9.44x10^-5^  D’=0.15 | R^2^=5.37x10^-5^  D’=0.03 | R^2^=9.95x10^-4^  D’=0.04 |
| **rs4742565** | Binary top locus | R^2^=4.83x10^-3^  D’=0.32 | R^2^=2.47x10^-3^  D’=0.06 | R^2^=2.68x10^-4^  D’=0.04 |

**Supplementary Figure 8.** Dose-adjusted clozapine concentrations between cohorts. *P* indicates the p-value of the dose-adjusted clozapine concentration between the Hacettepe cohort and the other cohorts.

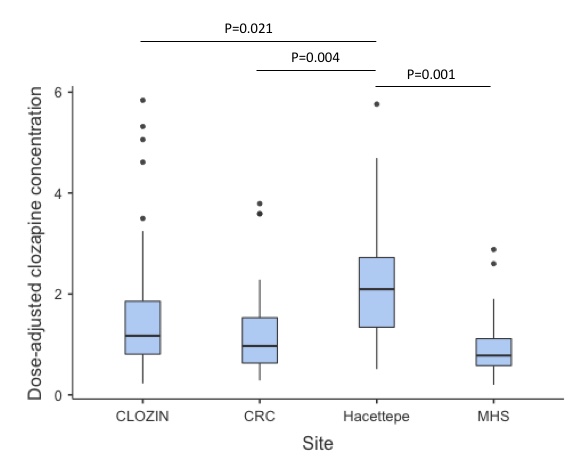

**Supplementary Figure 9A-B.** Bar plots illustrating the explained variance by PRSs, with exclusion of the GROUP cohort, at several *p*-value thresholds (p_t_), adjusted for sex, age, and 10 PCs. p_t_ are displayed on the X axis, where the number of included SNPs increases with more lenient p_t._ ΔExplained variance represents the Nagelkerke R^2^ (shown as %). The red dots represent the strengths of the association results (-Log 10 *p-*value). The dashed line represents a significance-level of *p*-value*<*0.05.

*Abbreviations:* PRS=Polygenic Risk Score, PCs=Principal Components, SNPs=Single Nucleotide Polymorphisms.

**A.** Binary outcome schizophrenia-PRS	**B.** Quantitative outcome cross-disorder-PRS

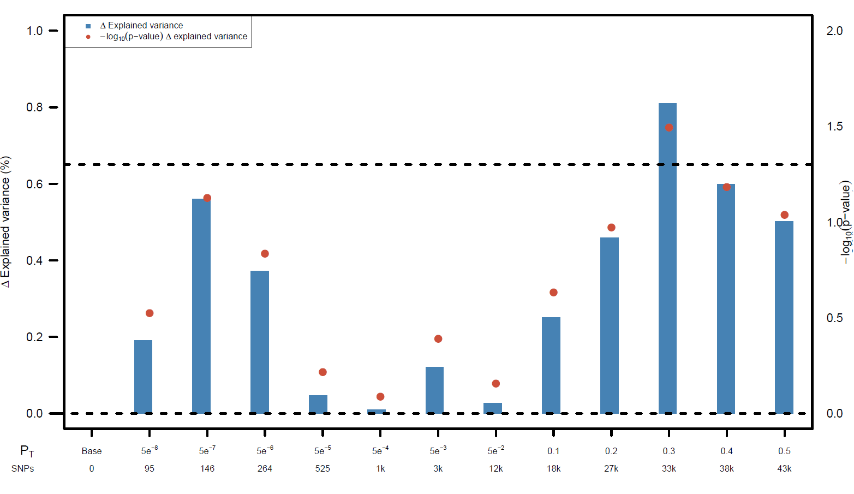

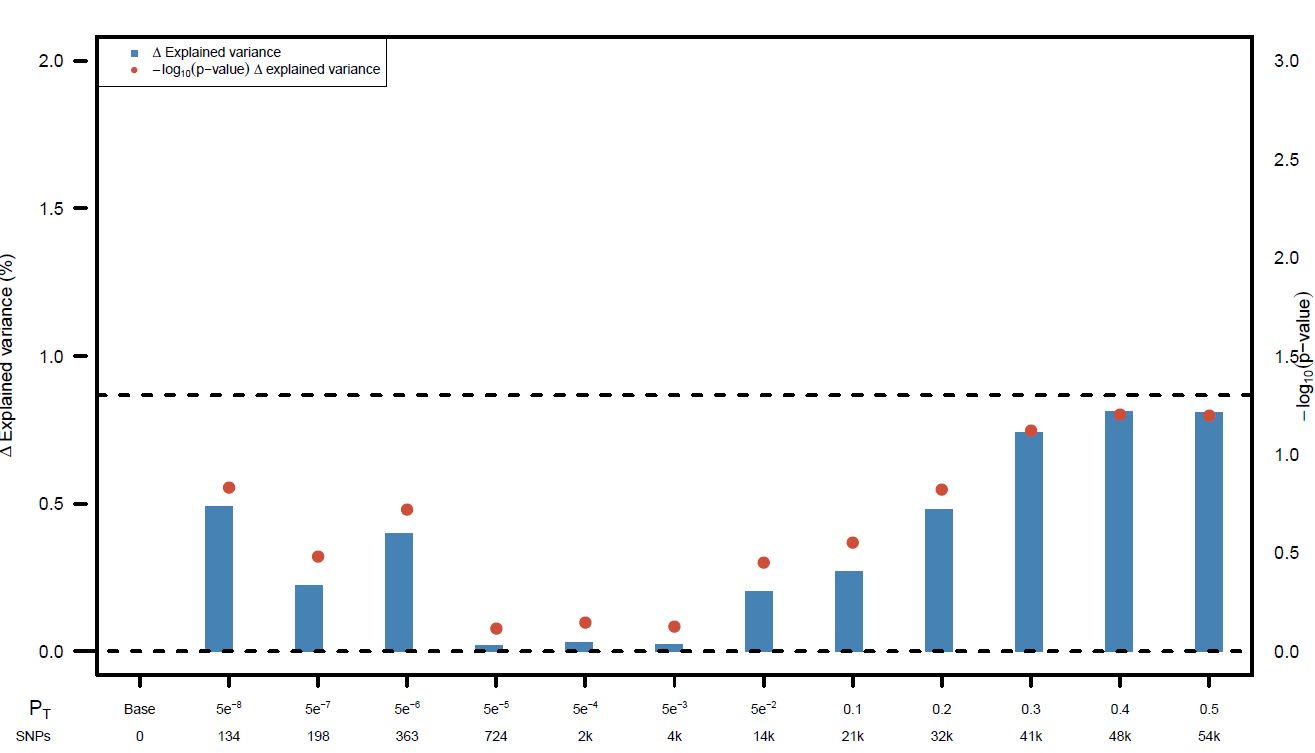

**Supplementary Figure 10A&B.** Heatmap plots using 54 tissues from GTEx for quantitative outcome (A) and binary outcome (B). The expression values were average of normalized expression per label (zero mean across samples).

**A**
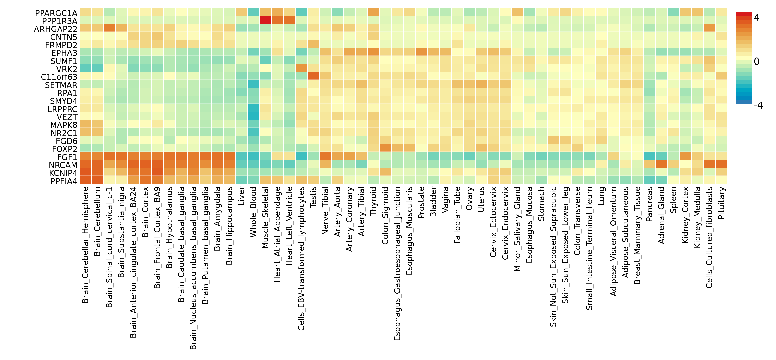

**B**
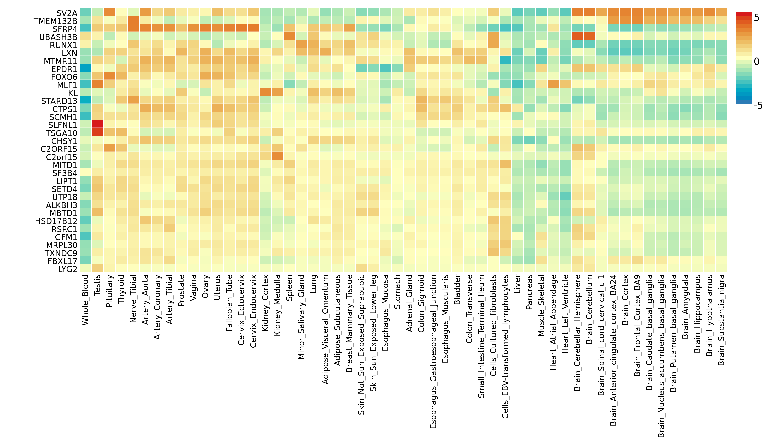

**Supplementary Figure 11A-D.** Gene-based association results from H-MAGMA using Hi-C interaction for all outcomes. The X-axis indicates the start position of genes (hg19).

**A.** Quantitative outcome in fetal brain

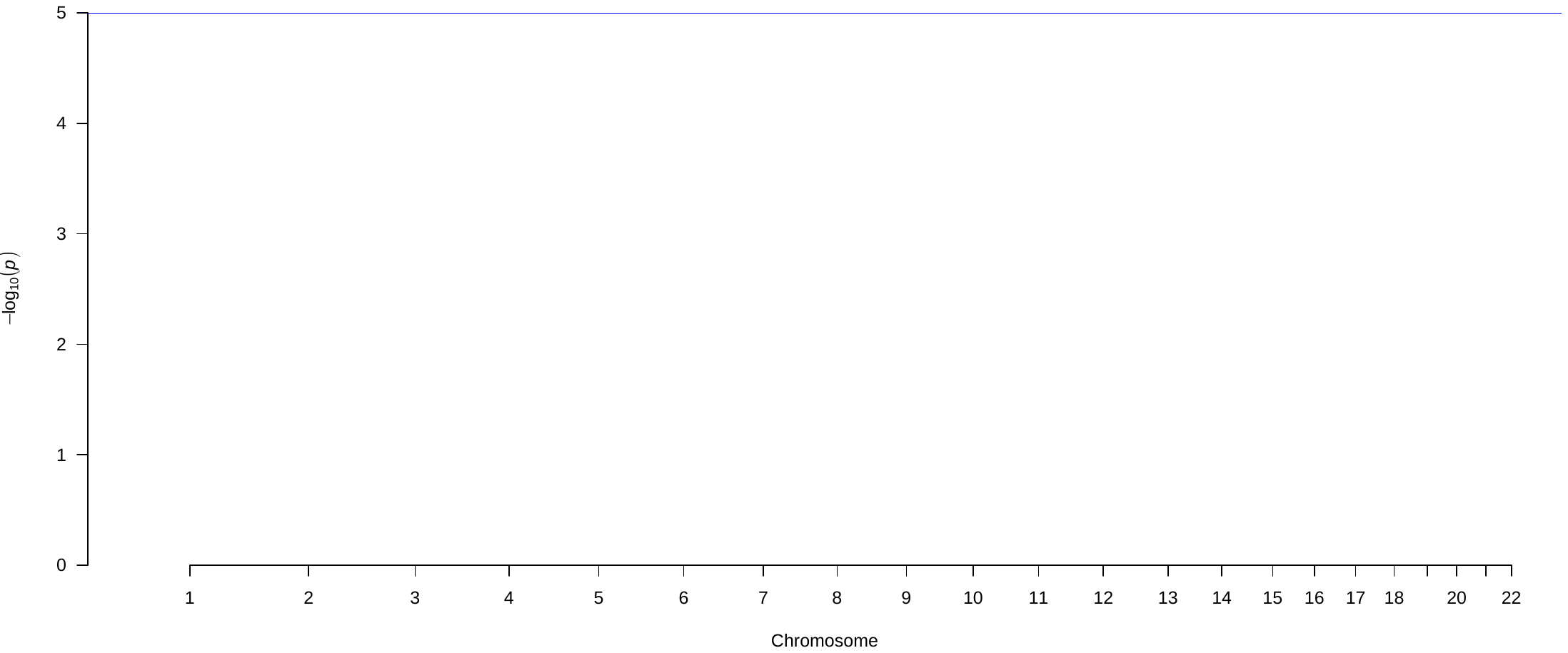

**B.** Quantitative outcome in adult brain

**C.** Binary outcome in fetal brain

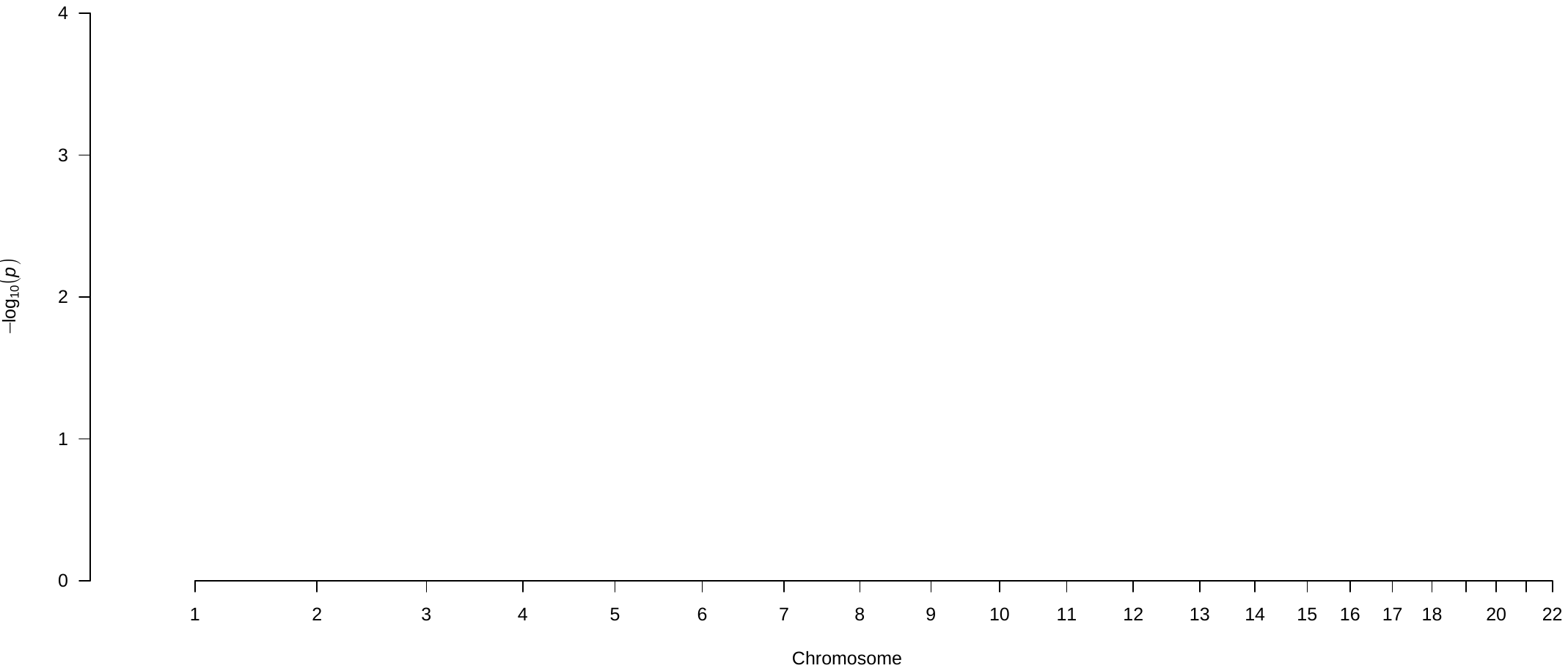

**D.** Binary outcome in adult brain

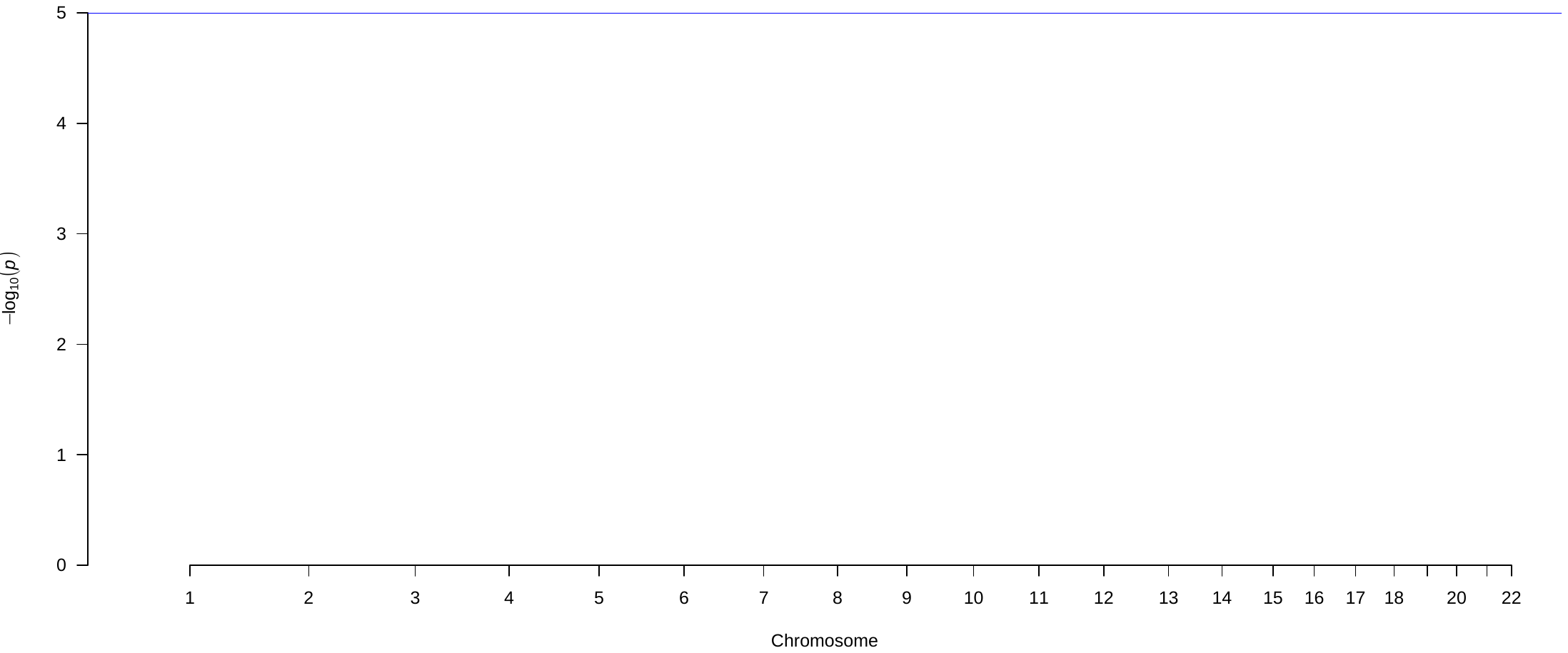

**Supplementary Figure 12A-D.** Gene ontologies enriched for symptom severity linked to genes for each outcome.

**A.** Quantitative outcome for fetal brain

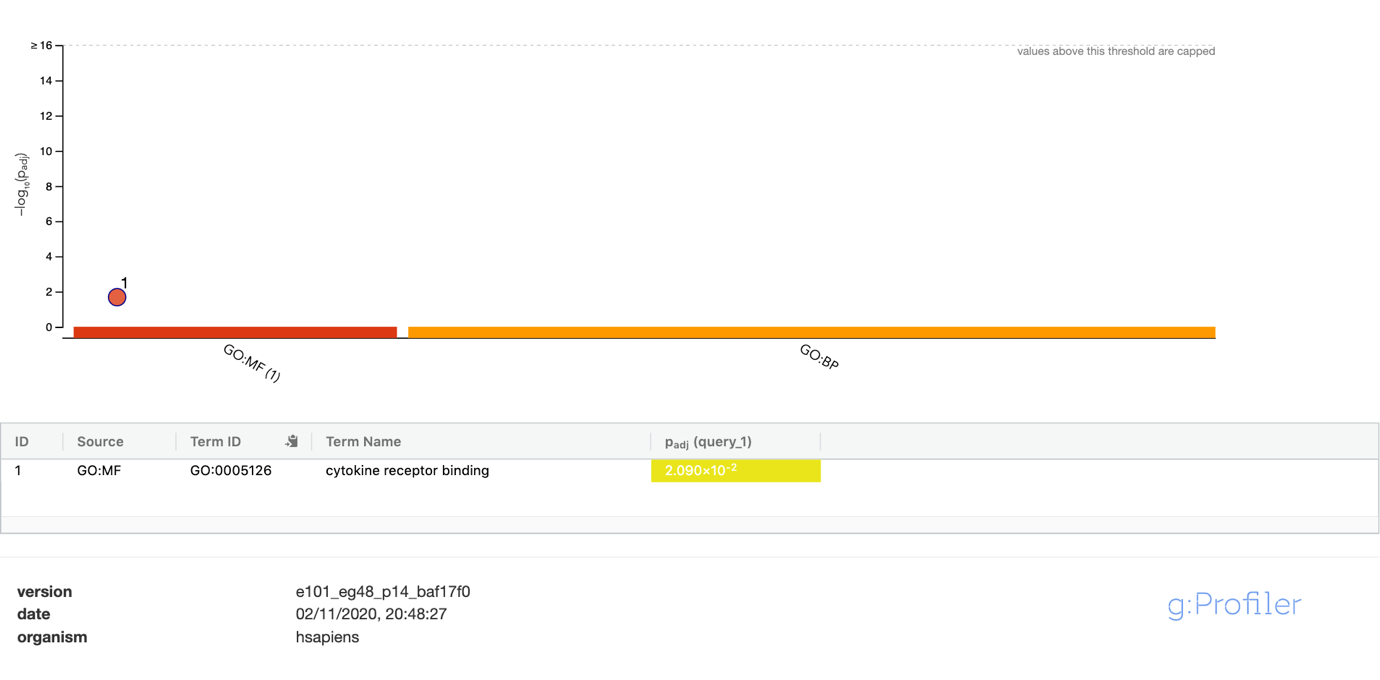

**B.** Quantitative outcome for adult brain

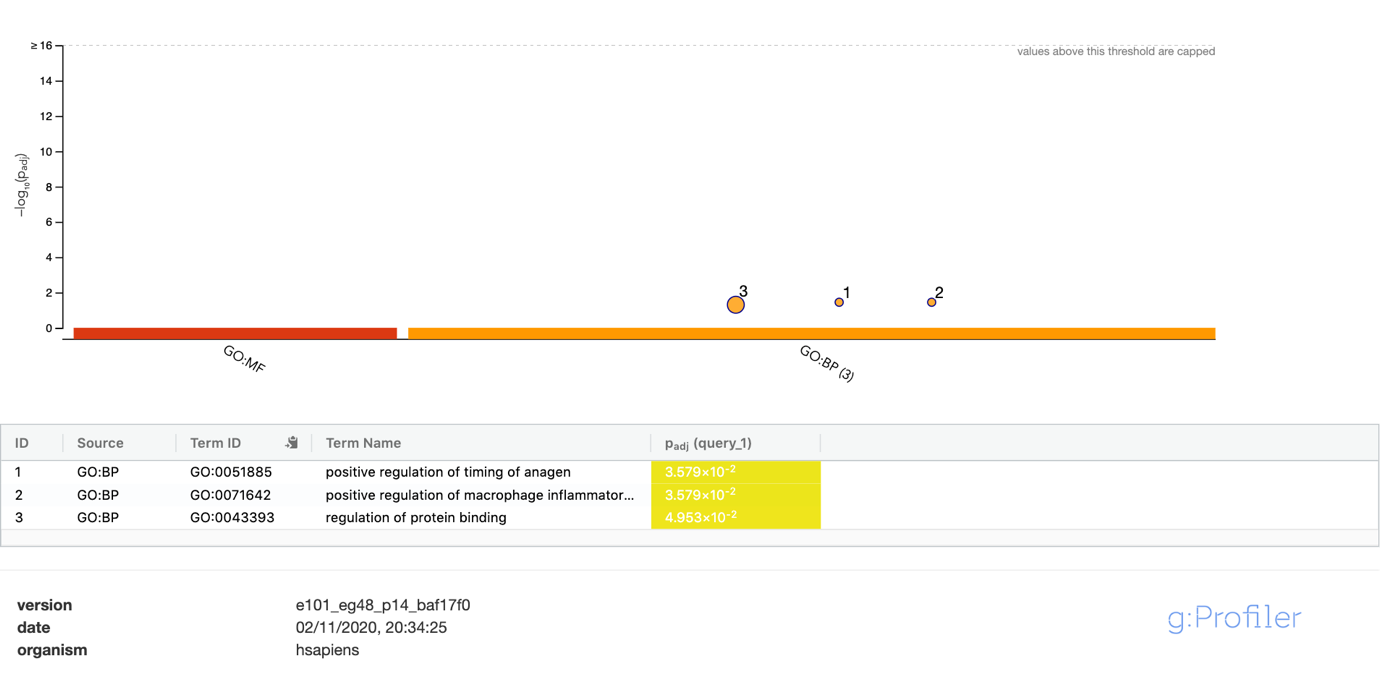

**C.** Binary outcome for fetal brain

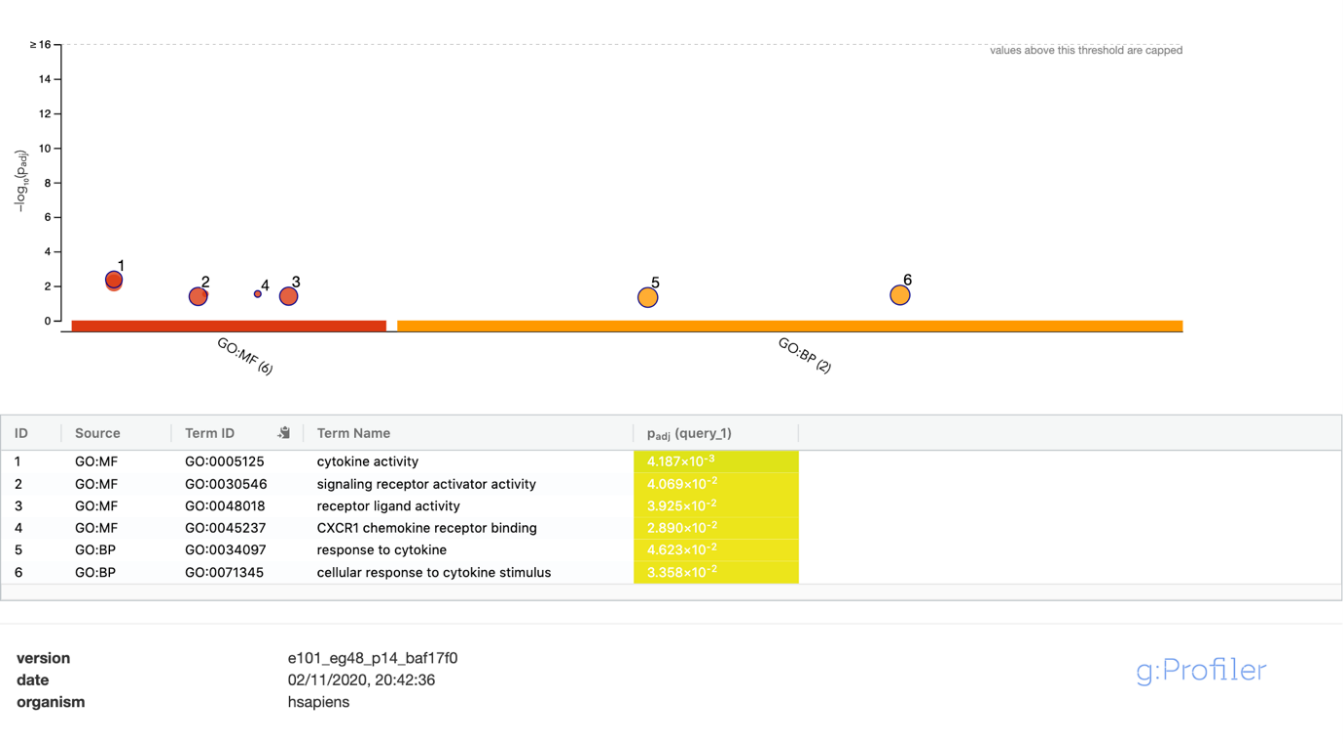

**D.** Binary outcome for adult brain

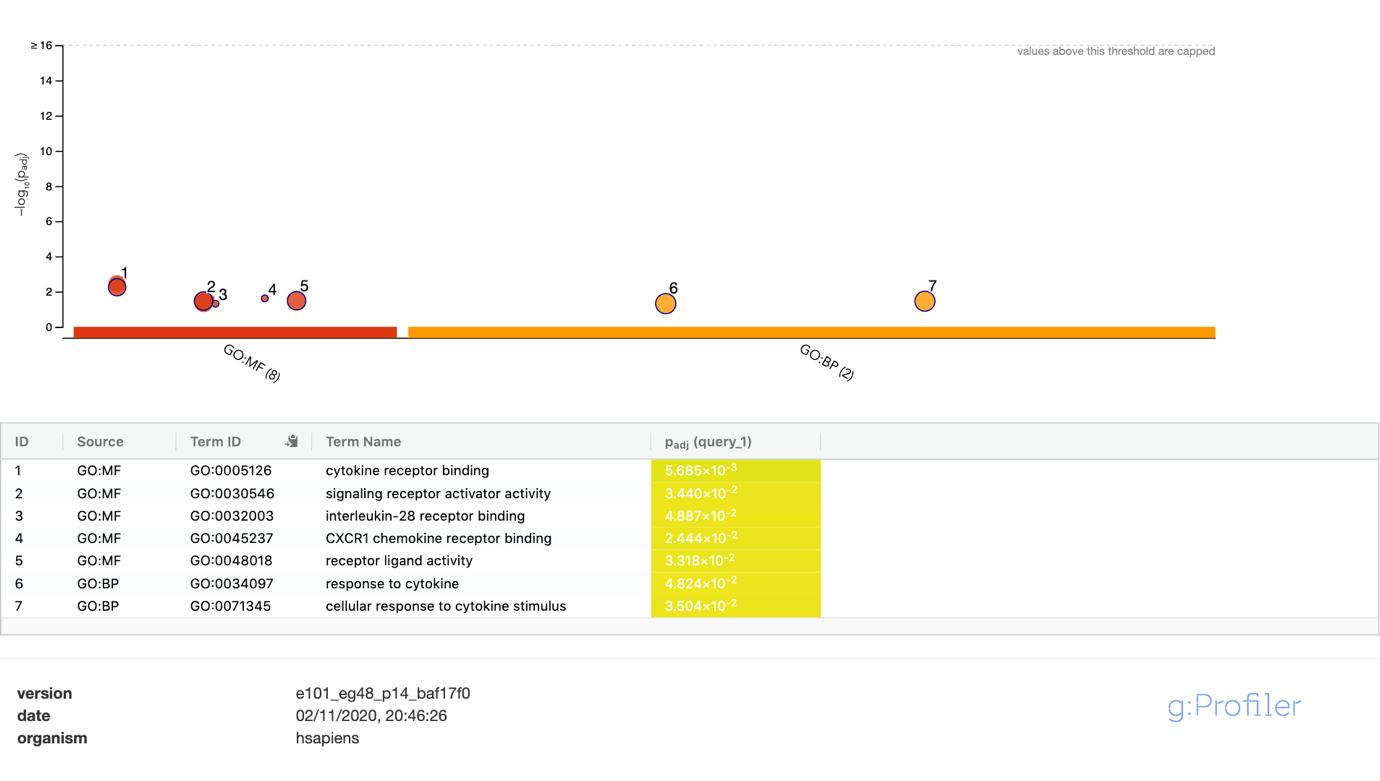

**Supplementary Figure 13**. A) Visual plot of the mean (including 95% confidence interval (CI) and median of dose-adjusted clozapine concentrations and high vs. low symptom severity; no differences were detected between those with high and low symptom burden, B) Visual plot of the spread of dose-adjusted clozapine concentrations and Clinical Global impression-Severity (CGI-S) score.

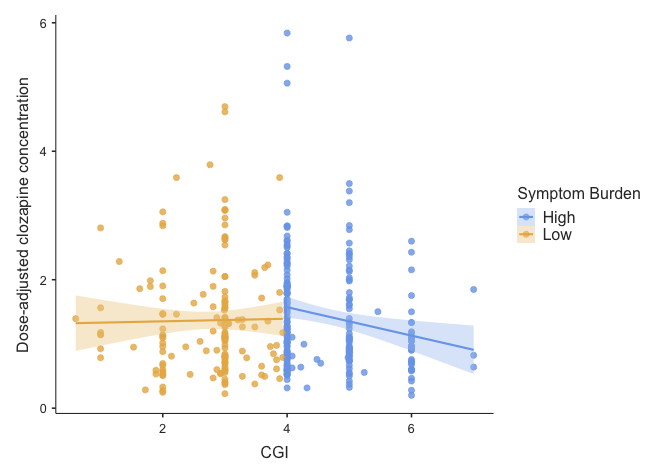

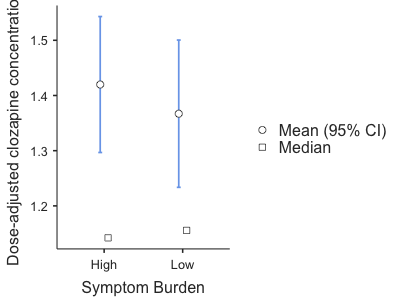

A)	B)

**Supplementary Figure 14A-D**. Dose-adjusted clozapine concentrations (Cohen’s d**=**0.76; R^2^**=**0.122, *p***=**1.65x10^-11^) and CGI-S score (Cohen’s d**=**0.21; R^2^**=**0.008, *p***=**9.07x10^-3^) by smoking status (A-B), sex (C; Cohen’s d**=**0.51; *p***=**1.10x10^-5^) and smoking status and sex (D; Male Smoker: mean(SD)=1.08(0.72), Female Smoker: mean(SD)=1.22(0.68), Male Non-smoker: mean(SD)=1.56(0.93), Female Non-smoker: mean=2.03(0.95).

*Abbreviations: p*=*p*-value, CGI-S=Clinical Global Impression-Severity.

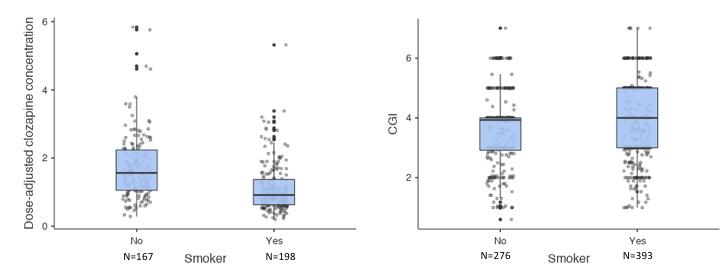

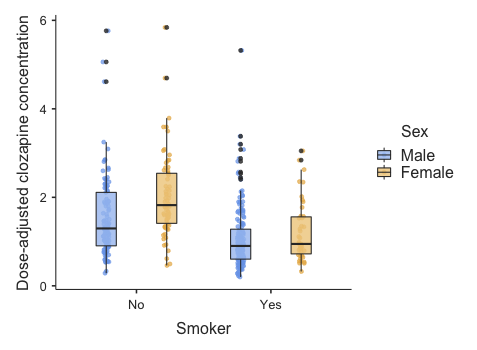
**A	B**

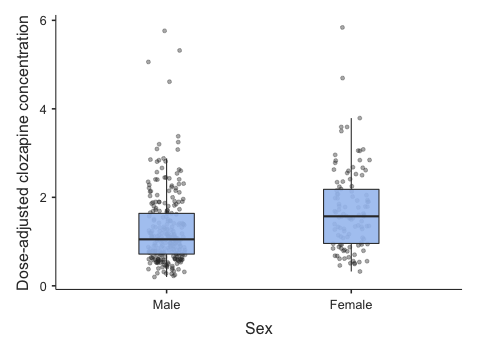

**C	D**

**Supplementary Acknowledgements**

First of all, we would like to thank Russel Lawrence Cummins, experienced expert, who we asked to comment on the importance of this manuscript. He commented: *‘I favor a mental healthcare where "cure and care" go more hand in hand. When reading this article, I realized this research is bound to prevent and alleviate much suffering among the hardest hurt by schizophrenia as it may help such people in the future to receive appropriate treatment more timely.’*

Furthermore, would like to thank prof. dr. R. Kahn for his feedback on our methods and first results, and for partly sponsoring the CLOZIN study. We also would like to thank M.K. Bakker for his statistical input, and E. Bekema for handling the samples and arranging shipments. Last, we would like to thank H. Gijsman, A. Jongkind, P. Kleymann for their help with recruiting participants.

**References**

1.	National Institute of Mental Health. CGI. Clinical Global Impressions. ECDEU Assess Man Psychopharmacol Revis. 1976;

2.	Sheehan D V, Lecrubier Y, Sheehan KH, Amorim P, Janavs J, Weiller E, et al. The Mini-International Neuropsychiatric Interview (M.I.N.I.): the development and validation of a structured diagnostic psychiatric interview for DSM-IV and ICD-10. J Clin Psychiatry. 1998;59 Suppl 2:22–57.

3.	Howes OD, McCutcheon R, Agid O, de Bartolomeis A, van Beveren NJM, Birnbaum ML, et al. Treatment-Resistant Schizophrenia: Treatment Response and Resistance in Psychosis (TRRIP) Working Group Consensus Guidelines on Diagnosis and Terminology. Am J Psychiatry. 2017 Mar;174(3):216–29.

4.	Kay SR, Fiszbein A, Opler LA. The positive and negative syndrome scale (PANSS) for schizophrenia. Schizophr Bull. 1987;

5.	Anıl Yağcioğlu AE, Yoca G, Ayhan Y, Karaca RÖ, Çevik L, Müderrisoğlu A, et al. Relation of the Allelic Variants of Multidrug Resistance Gene to Agranulocytosis Associated With Clozapine. J Clin Psychopharmacol. 2016 Jun;36(3):257–61.

6.	Hun Senol S, Gurcan G, Ertugrul A, Karahan S, Anil Yagcioglu AE. Augmentation of Clozapine Due to Inadequate Treatment Response in Schizophrenia: Comparison of Patients with Augmented and Non-augmented Treatments. Turkish J Psychiatry. 2020;

7.	Leucht S, Kane JM, Etschel E, Kissling W, Hamann J, Engel RR. Linking the PANSS, BPRS, and CGI: Clinical implications. Neuropsychopharmacology. 2006;

8.	Leucht S, Rothe P, Davis JM, Engel RR. Equipercentile linking of the BPRS and the PANSS. Eur Neuropsychopharmacol. 2013;

9.	Busner J, Targum SD. The clinical global impressions scale: applying a research tool in clinical practice. Psychiatry (Edgmont). 2007 Jul;4(7):28–37.

10.	Leucht S, Davis JM. What is the “best intro”—explanatory versus pragmatic antipsychotic drug trials. The Lancet Psychiatry. 2020.

11.	Anderson CA. Data Quality Control. In: Analysis of Complex Disease Association Studies. 2011.

12.	Coleman JRI, Euesden J, Patel H, Folarin AA, Newhouse S, Breen G. Quality control, imputation and analysis of genome-wide genotyping data from the Illumina HumanCoreExome microarray. Brief Funct Genomics. 2016;

13.	McCarthy S, Das S, Kretzschmar W, Delaneau O, Wood AR, Teumer A, et al. A reference panel of 64,976 haplotypes for genotype imputation. Nat Genet. 2016;

14.	Das S, Forer L, Schönherr S, Sidore C, Locke AE, Kwong A, et al. Next-generation genotype imputation service and methods. Nat Genet. 2016;

15.	Watanabe K, Taskesen E, van Bochoven A, Posthuma D. Functional mapping and annotation of genetic associations with FUMA. Nat Commun. 2017 Nov;8(1):1826.

16.	Watanabe K, Umićević Mirkov M, de Leeuw CA, van den Heuvel MP, Posthuma D. Genetic mapping of cell type specificity for complex traits. Nat Commun. 2019 Jul;10(1):3222.

17.	Sey NYA, Fauni H, Ma W, Won H. Connecting gene regulatory relationships to neurobiological mechanisms of brain disorders. bioRxiv. 2019.

18.	Wang D, Liu S, Warrell J, Won H, Shi X, Navarro FCP, et al. Comprehensive functional genomic resource and integrative model for the human brain. Science. 2018 Dec;362(6420).

19.	Won H, de la Torre-Ubieta L, Stein JL, Parikshak NN, Huang J, Opland CK, et al. Chromosome conformation elucidates regulatory relationships in developing human brain. Nature. 2016 Oct;538(7626):523–7.

20.	Reimand J, Kull M, Peterson H, Hansen J, Vilo J. g:Profiler--a web-based toolset for functional profiling of gene lists from large-scale experiments. Nucleic Acids Res. 2007 Jul;35(Web Server issue):W193-200.

21.	Choi SW, Mak TS-H, O’Reilly PF. Tutorial: a guide to performing polygenic risk score analyses. Nat Protoc. 2020 Sep;15(9):2759–72.

22.	Auton A, Brooks LD, Durbin RM, Garrison EP, Kang HM, Korbel JO, et al. A global reference for human genetic variation. Nature. 2015 Oct;526(7571):68–74.

23.	Lee S-B, Wheeler MM, Thummel KE, Nickerson DA. Calling Star Alleles With Stargazer in 28 Pharmacogenes With Whole Genome Sequences. Clin Pharmacol Ther. 2019 Dec;106(6):1328–37.

24.	Gaedigk A, Ingelman-Sundberg M, Miller NA, Leeder JS, Whirl-Carrillo M, Klein TE. The Pharmacogene Variation (PharmVar) Consortium: Incorporation of the Human Cytochrome P450 (CYP) Allele Nomenclature Database. Clin Pharmacol Ther. 2018 Mar;103(3):399–401.

25.	Whirl-Carrillo M, McDonagh EM, Hebert JM, Gong L, Sangkuhl K, Thorn CF, et al. Pharmacogenomics knowledge for personalized medicine. Clin Pharmacol Ther. 2012 Oct;92(4):414–7.

26.	Mrazek DA, Biernacka JM, O’Kane DJ, Black JL, Cunningham JM, Drews MS, et al. CYP2C19 variation and citalopram response. Pharmacogenet Genomics. 2011 Jan;21(1):1–9.

27.	Saiz-Rodríguez M, Ochoa D, Belmonte C, Román M, Vieira de Lara D, Zubiaur P, et al. Polymorphisms in CYP1A2, CYP2C9 and ABCB1 affect agomelatine pharmacokinetics. J Psychopharmacol. 2019 Apr;33(4):522–31.

28.	Lesche D, Mostafa S, Everall I, Pantelis C, Bousman CA. Impact of CYP1A2, CYP2C19, and CYP2D6 genotype- and phenoconversion-predicted enzyme activity on clozapine exposure and symptom severity. Pharmacogenomics J. 2020 Apr;20(2):192–201.

29.	Price AL, Weale ME, Patterson N, Myers SR, Need AC, Shianna K V, et al. Long-range LD can confound genome scans in admixed populations. Vol. 83, American journal of human genetics. 2008. p. 132–9.

30.	Flockhart, D. 2007. Drug Interactions: Cytochrome P450 Drug Interaction Table Indiana University School of Medicine.

31.	Ardlie KG, DeLuca DS, Segrè A V., Sullivan TJ, Young TR, Gelfand ET, et al. The Genotype-Tissue Expression (GTEx) pilot analysis: Multitissue gene regulation in humans. Science (80- ). 2015;

32.	Mayerova M, Ustohal L, Jarkovsky J, Pivnicka J, Kasparek T, Ceskova E. Influence of dose, gender, and cigarette smoking on clozapine plasma concentrations. Neuropsychiatr Dis Treat. 2018;14:1535–43.

33.	Wagner E, McMahon L, Falkai P, Hasan A, Siskind D. Impact of smoking behavior on clozapine blood levels - a systematic review and meta-analysis. Acta Psychiatr Scand. 2020 Sep;

34.	Smith RL, O’Connell K, Athanasiu L, Djurovic S, Kringen MK, Andreassen OA, et al. Identification of a novel polymorphism associated with reduced clozapine concentration in schizophrenia patients-a genome-wide association study adjusting for smoking habits. Transl Psychiatry. 2020 Jun;10(1):198.

35.	Li J, Yoshikawa A, Brennan MD, Ramsey TL, Meltzer HY. Genetic predictors of antipsychotic response to lurasidone identified in a genome wide association study and by schizophrenia risk genes. Schizophr Res. 2018 Feb;192:194–204.
